# Sociodemographic and geographic variation in mortality attributable to air pollution in the United States

**DOI:** 10.1101/2024.04.17.24305943

**Authors:** Pascal Geldsetzer, Daniel Fridljand, Mathew V. Kiang, Eran Bendavid, Sam Heft-Neal, Marshall Burke, Alexander H. Thieme, Tarik Benmarhnia

## Abstract

There are large differences in premature mortality in the USA by racial/ethnic, education, rurality, and social vulnerability index groups. Using existing concentration-response functions, particulate matter (PM_2.5_) air pollution, population estimates at the tract level, and county-level mortality data, we estimated the degree to which these mortality discrepancies can be attributed to differences in exposure and susceptibility to PM_2.5_. We show that differences in mortality attributable to PM_2.5_ were consistently more pronounced between racial/ethnic groups than by education, rurality, or social vulnerability index, with the Black American population having by far the highest proportion of deaths attributable to PM_2.5_ in all years from 1990 to 2016. Over half of the difference in age-adjusted all-cause mortality between the Black American and non-Hispanic White population was attributable to PM_2.5_ in the years 2000 to 2011.

## Main Text

Despite improvements in the overall life expectancy in the USA over the past decades, significant inequalities among different racial/ethnic and socioeconomic groups remain a public health challenge^1^. For example, in 2019, the life expectancy at birth was 78.8 years for the non-Hispanic (NH) White population but only 74.8 years for Black Americans^2^.

Air pollution, particularly exposure to fine particulate matter (PM_2.5_), is thought to be a major risk factor for premature death, both worldwide and in the USA^3^. Although most areas in the USA have seen declines in air pollution over the past decades, the pollution-related health burden remains substantial^3^. Improvements in air pollution exposure have been unequally distributed across population subgroups^4–6^. For instance, Tessum et al.^5^ showed an overall decrease in PM_2.5_ exposure in the USA between 2005 and 2015, but that such benefits were less pronounced among Black Americans, communities with lower educational attainment, and rural communities. Several federal environmental policies have been implemented since the 1970 Clean Air Act to address the fact that air pollution exposure differs across USA communities and acknowledge the environmental justice implications of exposure to air pollution. In particular, Executive Order 12898 (focusing on identifying and addressing the disproportionately high adverse human health effects from environmental exposures and on developing strategies for implementing environmental justice), Executive Order 14008 (focusing on climate change-related impacts and encouraging mitigations strategies that may also have co-benefits on greenhouse gas emissions and public health) and the National Ambient Air Quality Standard program state that air quality standards must be set at a level that protects the most vulnerable populations. More information on these policies is provided in the Supplementary Text.

In addition to being systematically more exposed to higher levels of air pollution, structurally disadvantaged communities are also thought to be more susceptible to adverse health effects from air pollution, which has sometimes been referred to as an environmental justice “double jeopardy”^7^. For example, the concentration-response function (CRF) linking PM_2.5_ exposure to mortality appears to be more pronounced among Black Americans^8^. Such differential susceptibility is thought to be due to differential distributions in pre-existing comorbidities, lack of access to health care, racialized occupational sorting into jobs with more hazardous exposures, and other social and structural determinants of health^7^. Indeed, there is a rich literature documenting how structural racism^9^, related to historical policies such as redlining as well as contemporary inequalities in health care access, can explain such differential susceptibility to air pollutants^10^.

Using existing CRFs, PM_2.5_ air pollution and population estimates at the census tract level, and mortality data at the county level, the aim of this study was to quantify the contribution of PM_2.5_ to racial/ethnic, educational, geographic, and social vulnerability-related inequalities in mortality in the USA to inform appropriate policy interventions. Studies that estimate the number of deaths attributable to air pollution, and PM_2.5_ in particular, usually rely on existing CRFs from the literature to infer such burden^11^. These studies have used a single CRF for the whole pooled population, assuming no differences in susceptibility to PM_2.5_, which may drastically underestimate the unequal health burden of air pollution across vulnerable communities. Given the evidence on differences in susceptibility to PM_2.5_ by racial/ethnic groups in the USA ^9–14^, we used the CRF from Di et al.^15^ for our primary analyses. The CRF from Di et al. is the only existing CRF that provides race-ethnicity-specific estimates for each of the main racial/ethnic categories in the US census and is based on a large sample of the population from all US counties^15^. Our analysis relies on the assumption that this CRF accurately depicts a causal effect of PM_2.5_ exposure on mortality. To examine the degree to which the use of CRFs that ignore differences in the concentration-response association by racial/ethnic group underestimates disparities in PM_2.5_-attributable mortality between racial/ethnic groups, we then compare our results to those obtained from two recent and widely used uniform CRFs for the USA population in secondary analyses ^11,16^. In this study, we use the classification of racial/ethnic groups as a proxy for long-established and systemic consequences of political, historical, and economic structures, social constructs as well as environmental racism^10,12,17^.

## Results

### Exposure to PM_2.5_ at the national level

In 2016, 0.8% of all census tracts and 0.9% of the overall population were exposed to an annual mean PM_2.5_ concentration above 12µg/m^3^, which is the legally required threshold set by the current National Ambient Air Quality Standard^18^. In comparison, in 1990, 83.4% of census tracts and 85.9% of the population were exposed to PM_2.5_ levels above 12µg/m^3^. The decline in PM_2.5_ exposure (as well as all-cause mortality) overall, and separately by subpopulation over time is plotted in figures Fig. S 2 - Fig. S 7. The mean population-weighted PM_2.5_ exposure levels, when averaged over the period from 2000 to 2016, were highest among Black Americans at 9.38 μg/m³, followed by Asians or Pacific Islanders at 9.21 μg/m³, Hispanics or Latinos at 9.13 μg/m³, Non-Hispanic Whites at 8.25 μg/m³ and American Indians or Alaska Natives at 7.46 μg/m³ (Fig. S 4). Between 2000 and 2016, the disparity in PM_2.5_ exposure between Black Americans and Non-Hispanic Whites narrowed from 2.17 μg/m³ to 0.94 μg/m³.

### Estimated PM_2.5_-attributable mortality at the national level

In the overall population, the estimated PM_2.5_-attributable mortality declined from 79.2 (95% CI, 77.1 to 81.4) age-adjusted deaths per 100,000 in 1990 to 11.7 (95% CI, 11.4 to 12.0) in 2016. We observed this steep downward trend for all studied subpopulations, with absolute differences between these groups narrowing considerably over the study period (**Fig. 1**, Fig. S. 8). However, the lines representing each subpopulation in figure 1 did not intersect at any time between 1990 and 2016. Thus, the most affected groups in 2016 were also the most affected groups in 1990.

**Fig. 1.**
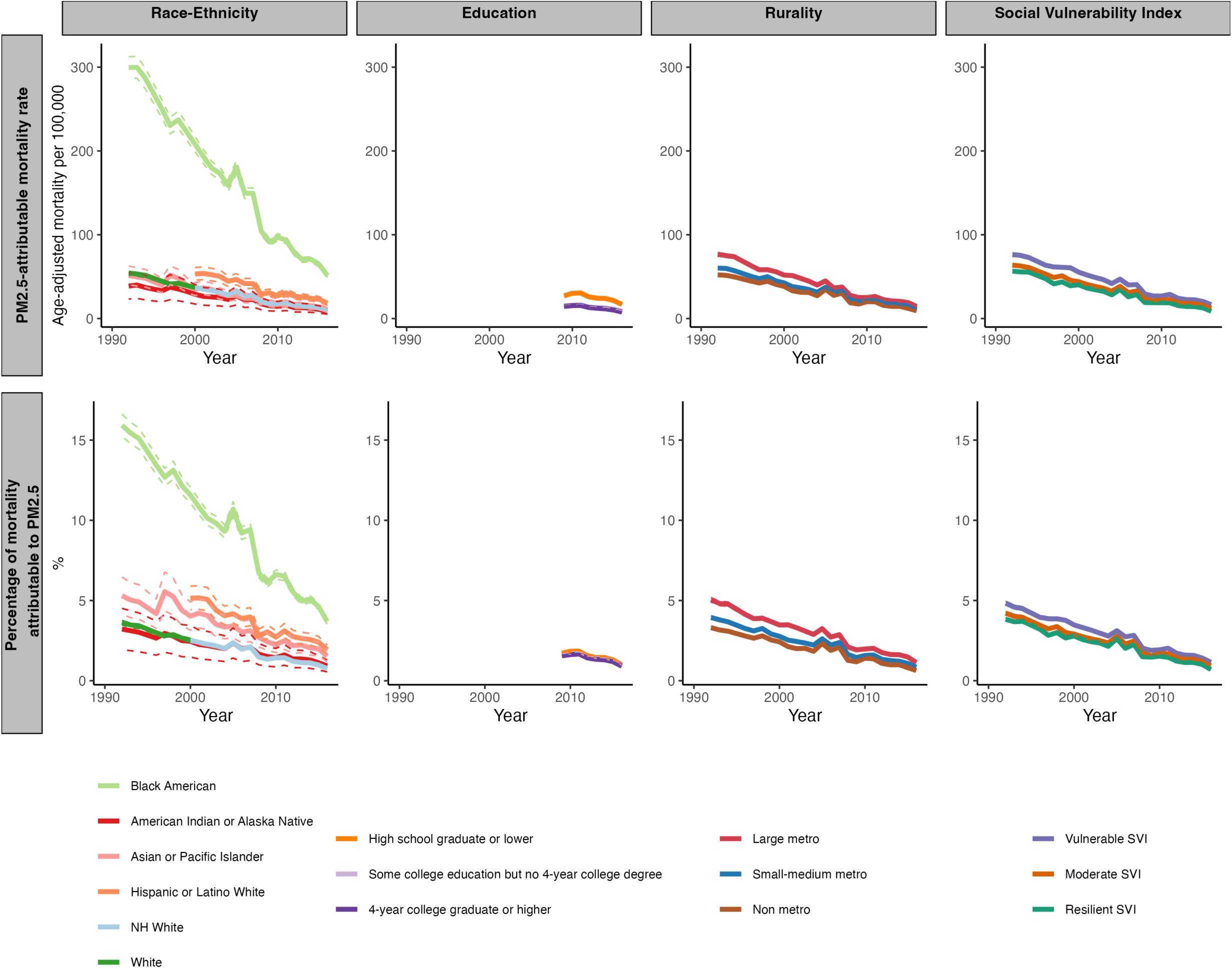
Age-adjusted PM_2.5_-attributable mortality rate by racial/ethnic group, education level, rurality level, and the social vulnerability index. The first row shows the age-adjusted mortality rate that we estimate is attributable to PM2.5. The second row shows the percentagae of all-cause mortality that we estimate is attributable to PM2.5. The dashed lines depict 95% confidence intervals. 95% confidence intervals for rurality are too narrow to be visible. Abbreviations: NH=Non-Hispanic, SVI=social vulnerability index.

From 1990 to 2016, Black Americans experienced an estimated 86% reduction in the age-adjusted mortality rate attributable to PM2.5, the largest decline observed among all racial/ethnic groups (Fig. 1). However, these larger improvements among Black Americans must be considered in relation to their higher starting point in the PM_2.5_-attributable mortality rate relative to other racial/ethnic groups. In fact, although the absolute size of the difference in the PM_2.5_-attributable age-adjusted mortality rate between Black Americans and other racial/ethnic groups decreased over the study period, the relative size of these differences remained similar over time (Fig. S 9). Examining the estimated percent of all-cause mortality that is attributable to PM_2.5_, rather than the PM_2.5_-attributable mortality rate, for each population group yielded similar trends (Fig. 1). Of note, the declines in PM_2.5_ exposure over the study period have led to a decrease in the percent of all-cause mortality that is attributable to PM_2.5_ among each of our population subgroups. Unlike the decline in the PM_2.5_-attributable mortality rate over time, this finding was not self-evident because it indicates that for each population subgroup mortality from PM_2.5_ decreased more rapidly than mortality from other causes.

More than half of the difference in all-cause mortality between Black Americans and the Non-Hispanic White population was attributable to PM_2.5_ in each year from 2000 to 2011 (**Fig. 2**). With a decrease from 53.4% (95% CI, 51.2% to 55.9%) in 2000 to 49.9% (95% CI, 47.8% to 52.2%) in 2015, this proportion, however, has declined over time. The percent of the difference in all-cause mortality to Black Americans that can be explained by differences in exposure and susceptibility to PM_2.5_ between racial/ethnic groups was lower for the Hispanic or Latino White, Asian or Pacific Islander, and American Indian or Alaska Native population than for the Non-Hispanic White population. Nonetheless, at a mean for the period 2000 to 2015 of 20.8% (95% CI, 20.7% to 20.9 %), 16.1% (95% CI, 16.1% to 16.2%) and 12.8% (95% CI, 12.7% to 12.8%) for the American Indian or Alaska Native, Hispanic or Latino White, Asian or Pacific Islander, and population, respectively, the percentages were still substantial.

**Fig. 2.**
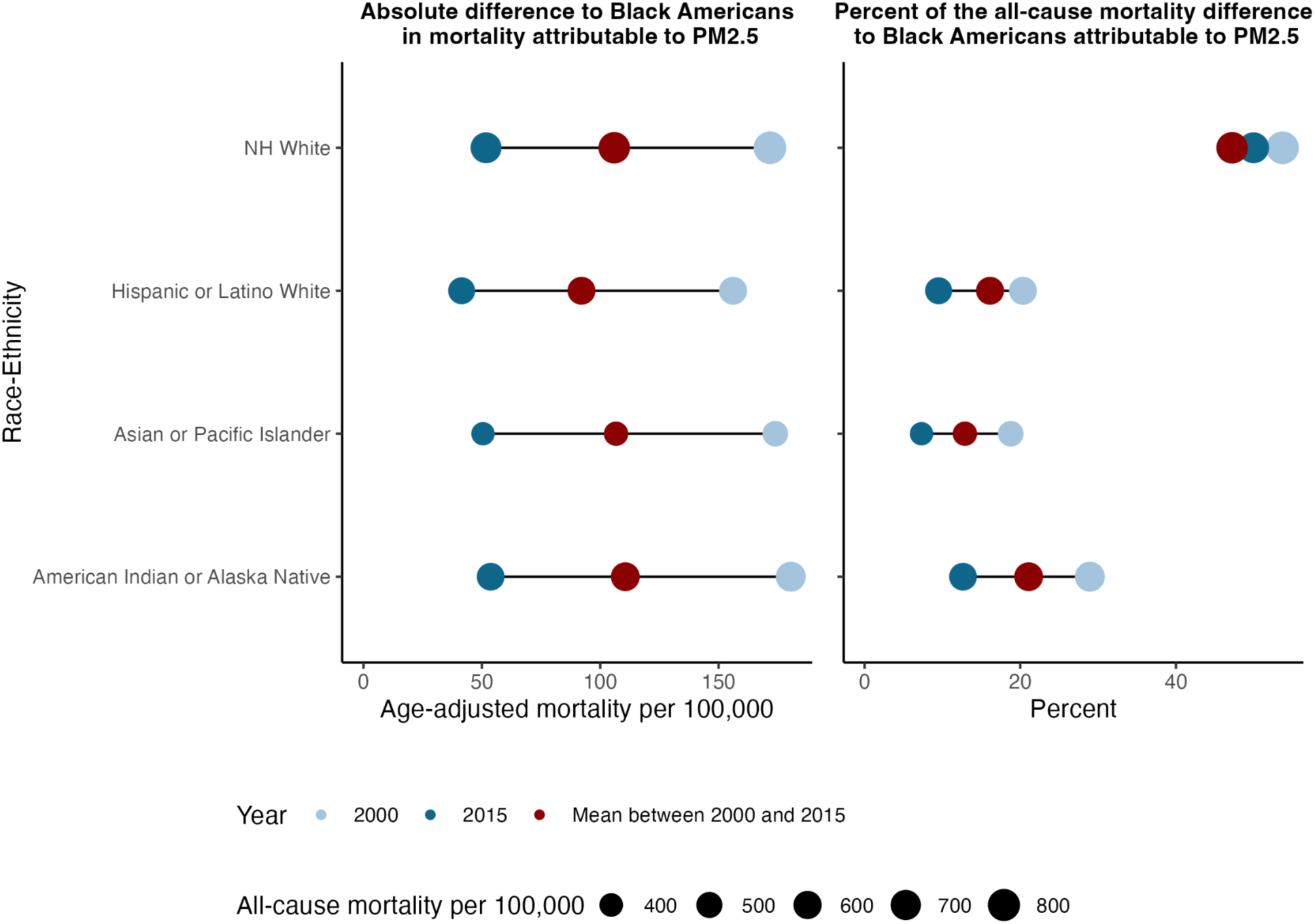
Extent to which the difference in the age-adjusted mortality rate between each racial/ethnic group and Black Americans can be attributed to PM_2.5_. Of the racial/ethnic groups considered in this study, Black Americans have both the highest all-cause mortality rate and highest PM2.5-attributable mortality rate. The first column is the absolute size (in mortality per 100,000) of the difference in mortality attributable to PM2.5 between each racial/ethnic group and Black Americans. The second column is the percent of the difference in all-cause mortality between each racial/ethnic group and Black Americans that can be attributed to PM2.5 exposure. Dark blue dots denote 2015 and light blue dots denote 2000. Red dots denote the unweighted mean from 2000 to 2015. The dot sizes are proportional to the age-adjusted all-cause mortality rate that is attributable to PM2.5. Abbreviations: NH=Non-Hispanic

We observed that disparities between racial/ethnic groups in PM2.5-attributable mortality exist at all levels of education, rurality, socioeconomic status, household characteristics, housing type and transportation, and social vulnerability index (**Figures 3**, S 10 – S 15). In fact, variation in PM2.5-attributable mortality by racial/ethnic group further increased when the analysis was restricted to the population with high educational attainment or those residing in non-metropolitan areas (Fig. S 16). The variation in PM2.5-attributable mortality by racial/ethnic group was similar for the different levels of socioeconomic status, social vulnerability index, and housing type and transportation (Fig. S 16). Black Americans and Hispanic or Latino Whites had the highest PM2.5-attributable mortality at all levels of these variables in the year 2016 (Figures 3, S 10 – S 14). Black Americans also had a higher PM2.5-attributable mortality than the respective average for every level of these variables in 2016 (Figures 3, Fig. S 10– Fig. S 14). Hispanic or Latino Whites living in socially vulnerable counties experienced double the PM2.5 attributable mortality than Hispanic or Latino Whites living in socially resilient counties in 2016 (Fig. S 10, Fig. S 17).

**Fig. 3.**
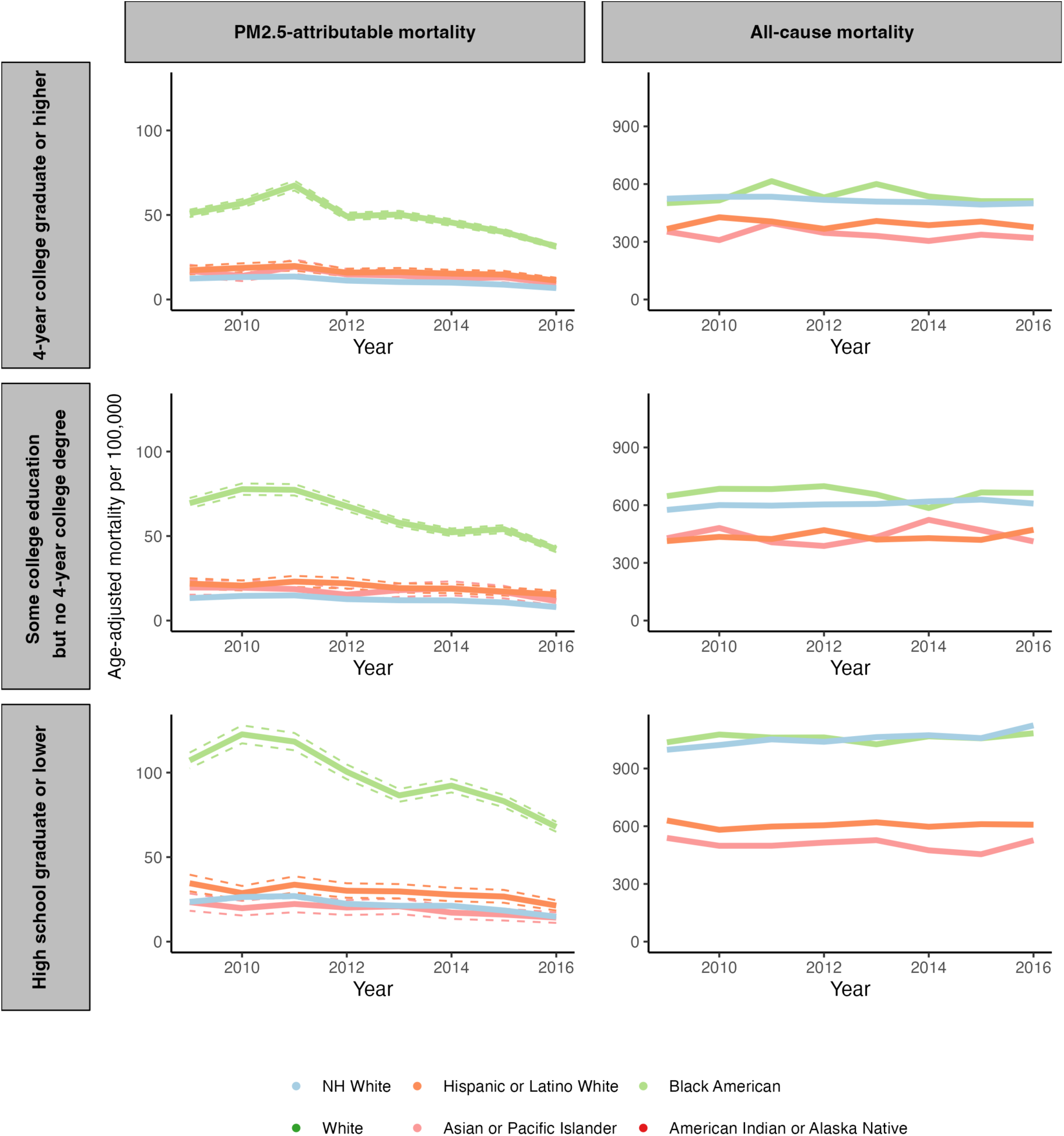
Age-adjusted PM_2.5_-attributable mortality rate and all-cause mortality rate for each racial/ethnic group stratified by educational attainment. The dashed lines depict 95% confidence intervals. The US Census Bureau has not published data on educational attainment by age for the racial/ethnic group “American Indian or Alaska Native”^34^. This racial/ethnic group has, thus, been omitted from this figure. Abbreviations: NH=Non-Hispanic.

Those with low education and Black Americans had a higher PM2.5-attributable mortality than the respective average for every level of rurality, socioeconomic status, housing type and transportation, and social vulnerability index in 2016. Disparities in PM2.5-attributable mortality by educational attainment were more pronounced for high socioeconomic status than for middle and low socioeconomic status (Fig. S 18, Fig. S 19). Those with a high school diploma or a lower level of education experienced a higher PM2.5-attributable mortality than those with a higher educational attainment at all levels of rurality, social vulnerability index, socioeconomic status, household characteristics, minority status, and housing type and transportation (Fig. S 19 - Fig. S 24). High school graduates or lower living in large metropolitan areas experienced 36.9% (95% CI, 34.5% to 39.3%) higher PM_2.5_ attributable mortality than those with the same level of education living in non metro areas in 2016 (Fig. S 25).

Disparities in PM_2.5_-attributable mortality by education or rurality were not as pronounced as for racial/ethnic groups (Fig. 1). This finding was also corroborated when adapting a coefficient of variation approach (15), which found that estimated age-adjusted PM_2.5_-attributable mortality varied more by racial/ethnic group than by education or rurality in all years of our study period (Fig. S 26). Nonetheless, differences in PM_2.5_-attributable mortality by education and by rurality were still apparent. In the year 2016, those with a high school diploma or lower experienced 16.9 (95% CI, 16.4 to 17.4) PM_2.5_-attributable age-adjusted deaths per 100,000 (Fig. 1). This rate is nearly double compared to those with some college education but no 4-year degree, who had a rate of 8.8 (95% CI, 8.5 to 9.0), and those with a 4-year college degree or higher, who had a rate of 7.7 (95% CI, 7.5 to 7.9) (Fig. 1). Similar patterns are observed in all-cause mortality rates (Fig. S 5). Between 1990 and 2016, there was a significant reduction in the absolute differences in PM_2.5_-attributable age-adjusted mortality rates per 100,000 between large metro and non-metro areas, declining from 28.7 to 5.3. Despite this decrease in absolute disparity, the relative difference remained stable and even showed a slight increase (Fig. 1). Specifically, large metro areas had 1.48 times the PM_2.5_-attributable mortality rate of non-metro areas in 1990, and this ratio increased to 1.57 times in 2016 (Fig. 1).

### Variation in estimated PM_2.5_-attributable mortality across states and counties

We observed substantial variation across states both in the age-adjusted PM_2.5_-attributable mortality rate (Fig. S 27) as well as the percent of age-adjusted all-cause mortality that can be attributed to PM_2.5_ (**Fig. 4**). Fig. S 28 depicts the state-level positive association between PM_2.5_ exposure and all-cause mortality. In all states with a PM_2.5_-attributable mortality rate above zero, Black Americans had a higher percent of all-cause mortality that can be attributed to PM_2.5_ (Fig. 4), as well as a higher PM_2.5_-attributable mortality rate (Fig. S 27), in 2016 than the Non-Hispanic White population. Similarly, those with a high school diploma or lower education had a higher PM_2.5_-attributable mortality in all states compared to groups with a higher level of education (Fig. S 28). In 31 of the 34 states that had both counties designated as “large metro” and “non metro”, “large metro” counties had a higher PM_2.5_-attributable mortality compared to “non metro” counties (Fig. S 28).

**Fig. 4.**
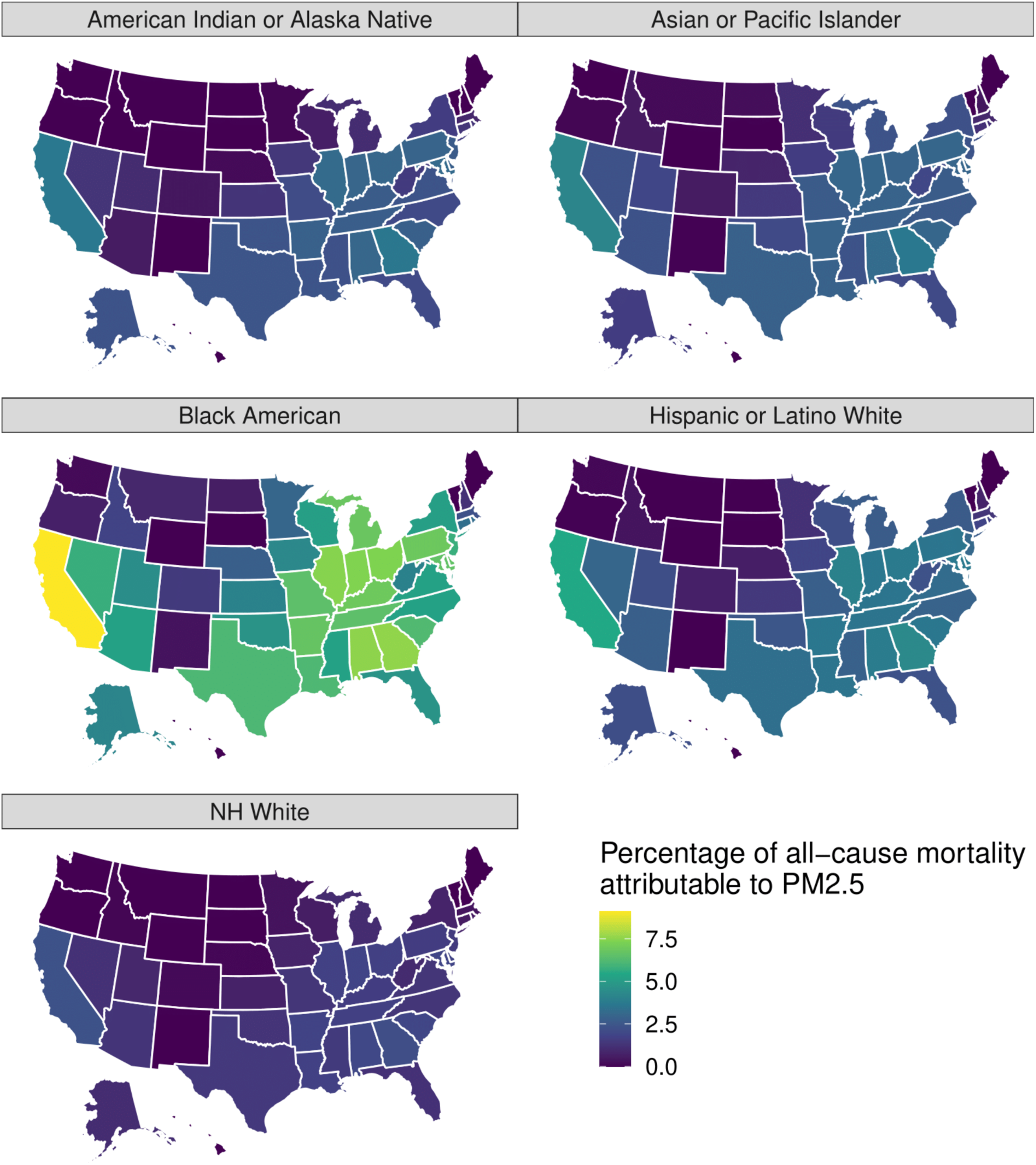
Percentage of the age-adjusted all-cause mortality rate that was attributable to PM_2.5_ in the year 2016, by state and racial/ethnic group. Abbreviations: NH=Non-Hispanic.

When examining variation across counties, we found that in virtually all (96.6%) counties for which data were available, Black Americans had a higher age-adjusted PM_2.5_-attributable mortality rate than the Non-Hispanic White population (**Fig. 5**, Fig. S 29) for the mean value across the period 2000 to 2016. The comparison between the Black American and Hispanic or Latino White population was more varied, with a number of counties in the South-Eastern USA having a lower PM_2.5_-attributable mortality rate for Black Americans than for Hispanic or Latino Whites. The lowest PM_2.5_-attributable mortality rates for all three racial/ethnic groups for which we had a sufficient sample size at the county level (Black American, Hispanic or Latino White, and Non-Hispanic White) tended to be in counties in the Mountain West. When taking into account spatial autocorrelation, we identified multiple county clusters with significantly higher or lower differences in the PM_2.5_-attributable mortality rate between racial/ethnic groups than in surrounding areas (Fig. S 30).

**Fig. 5.**
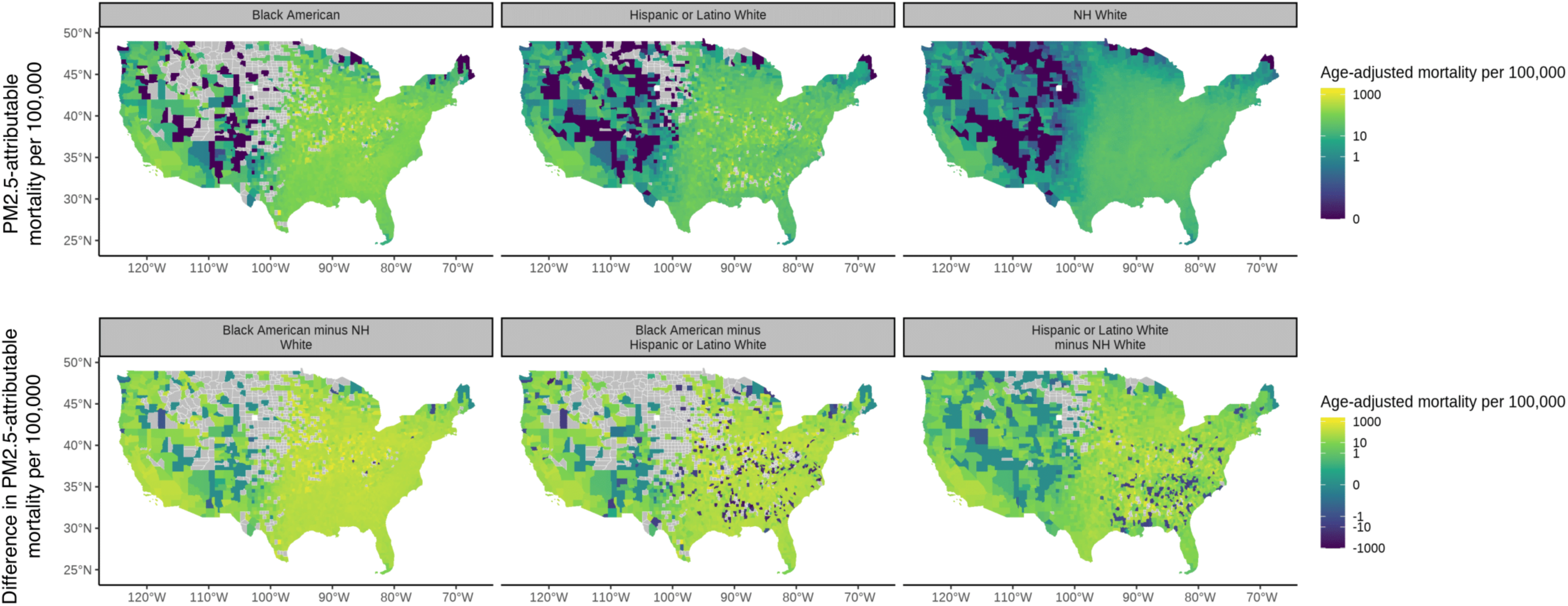
Differences in the age-adjusted PM_2.5_-attributable mortality rate between racial/ethnic groups at the county level for the period 2000 to 2016. These figures show the (unweighted) mean value across the period 2000 to 2016. The first row is the PM2.5-attributable mortality per 100,000 for each racial/ethnic group. The second row is the absolute difference in PM2.5-attributable mortality per 100,000 between racial/ethnic groups. Abbreviations: NH=Non-Hispanic.

All our analysis results, including differences in PM_2.5_-attributable mortality by racial/ethnic, education, level of rurality, and social vulnerability-related groups, as well as trends over time, remained similar in relative terms when restricting the study population to those aged 65 years and older (Figures S 31 – S 45).

## Discussion

Our study shows that improvements in air quality in the USA have decreased estimated PM_2.5_-attributable mortality for all subpopulations that we examined. However, our analysis also highlights the remaining inequalities in PM_2.5_-attributable mortality between different subpopulations. These inequalities were most pronounced between racial/ethnic groups. In fact, more than half of the difference in mortality between the Non-Hispanic White population and Black Americans was attributable to PM_2.5_ in each year from 2000 to 2011.

Our results indicate the strong association of the race and ethnicity group with adverse environmental health outcomes – an association that is even stronger than for education, rurality or social vulnerability-related factors. This finding aligns with a growing body of evidence ^9,10,19^ demonstrating that racial/ethnic categories are not simply proxies for socioeconomic differences but are also (imperfect) proxy measures for exposure to historical and contemporary discriminatory practices. Racism and the discrimination of racial/ethnic minorities emerged early in the founding of the USA and evolved into government-sponsored displacement, exclusion, and residential segregation^9,10,19^. Residential segregation is a substantial consequence of structural racism that still negatively impacts the health of racial/ethnic minorities today through various mechanisms involving both a disproportionate exposure to air pollutants and higher susceptibility to these pollutants^4,7,9,15,20–22^. Sources of air pollution emissions are often located in marginalized communities, as residents of these areas tend to have less economic opportunity, resources, and social capital, as well as limited political power, to influence the decision-making processes that determine where such sources of pollution are placed^9,10,20^. In addition to its role in explaining differences in PM_2.5_ exposure between racial/ethnic groups, structural racism is likely also a major driver of the unequal distribution of factors that contribute to the, on average, higher susceptibility to PM_2.5_ exposure among many racial/ethnic minorities^15^. These factors include social determinants of health, such as exclusion from job and educational opportunities, inadequate access to healthcare, and maladaptive coping behaviors^9^.

An additional important finding of our analysis is that inequalities in PM_2.5_-attributable mortality between racial/ethnic groups were obscured when ignoring differences in the estimated susceptibility to PM_2.5_ across racial/ethnic groups. Using race-ethnicity-specific mortality rates and PM_2.5_ exposure measurements, but assuming that the mortality effects of a given unit of PM_2.5_ are equal across all racial/ethnic groups, will greatly underestimate differences in PM_2.5_-attributable mortality. Similarly, using CRFs that do not take into account differences in susceptibility to PM_2.5_ between racial/ethnic groups will generally underestimate the benefit of reductions in PM_2.5_ on health outcomes among structurally disadvantaged groups. We, thus, advocate for the use of race-ethnicity-specific CRFs in future health impact studies to ensure air quality policies protect subpopulations most at risk.

We note some caveats of our study. A first set of limitations relates to the CRFs that we used in our analysis. First, although the race-ethnicity-specific CRF by Di et al.^15^ used in this analysis was developed in the age group 65+ years, our manuscript presents results for the age group 25+ years. To investigate whether our results would differ substantially if we conducted our study in this older age group only, we have implemented all analyses shown in this manuscript when restricting the study population to the age group 65+. The results (shown in the Supplementary Materials) demonstrate that all relative differences and patterns over time by racial/ethnic, education, and geography group remain similar. Second, we use a uniform CRF to derive the mortality response for all subpopulations not related to racial/ethnic group. There is evidence of differences in susceptibility to PM_2.5_ for the different subpopulations. For example, the composition of air pollution in urban areas has more adverse health effects than in rural areas^23^. We may, thus, underestimate the differences in PM_2.5_-attributable mortality between subpopulations not related to racial/ethnic group.

Third, because the true underlying PM_2.5_-attributable mortality rate is not practically measurable, we cannot estimate the validity of any given CRF. We therefore present results using multiple CRFs, selecting Di et al.’s model as our primary CRF due to its unique provision of race-ethnicity-specific estimates and its foundation in a nationwide sample^11,16,24^.

A second set of limitations relates to our PM_2.5_ exposure measurements. We relied on assigning PM_2.5_ exposure based on the current location of residence. This could be problematic for two reasons. First, if people live and work at different locations and pollution levels are systematically different between these locations, our attribution estimates may be biased. However, available evidence for urban areas finds the discrepancies in work and residential PM_2.5_ levels are small in absolute values (<0.1µg/m^3^)^25^, suggesting that assigning exposures by residence location may be a reasonable approximation of total exposure. A related concern arises if mortality is driven mainly by long-term cumulative exposure, populations change residence locations during the study period, and there exists a systematic correlation between exposure at previous residence location and exposure at current residence location. However, existing literature finds that both immediate and long-term exposure matter for health outcomes, and again does not find consistent support for patterns of geographic mobility being associated with pollution levels in the USA^26^. Another potential limitation associated with exposure assignment was due to pollution and population data being available at a geographically more granular level (0.01° by 0.01° and the census tract level, respectively) than our mortality data (available at the county level). As the geographic level at which we conduct our analysis increases, the risk of ecological bias increases.

However, it is worth noting that the ’smoothing’ effect of using county-level data is more likely to result in underestimation rather than overestimation of disparities between subpopulations. This is because the averaging process could dilute more extreme values that contribute to these disparities, making them appear less pronounced than they actually are. We, however, did not observe a substantial association between the proportion of all-cause mortality attributable to PM_2.5_ with within-county variation in PM_2.5_ (S32, Panels C), implying that aggregation of PM_2.5_ exposure from the census tract to the county level is unlikely to be an important source of bias in our analysis. Another limitation is that our final confidence intervals for PM2.5-attributable mortality do not incorporate uncertainty arising from the PM2.5-exposure estimates, because the PM2.5-exposure estimates on a 0.01° by 0.01° grid from Meng et al. only provide mean estimates (without uncertainty estimates).

Another limitation is that Di et al.’s CRF is derived from zip-code level data, while our study is based on coarser county-level data. Hence there is a difference in exposure measurement errors between those geographic levels. This could affect the validity of the point estimates and confidence interval of the CRF by Di et al. estimates when applied to our study. For the first point, a previous study^27^ suggests that the bias for the point estimate introduced by the exposure measurement error is generally small.

A third set of limitations of our study relates to our measurement of socioeconomic variables. First, we had to rely on racial/ethnic group as a proxy measure for exposure to racism. Richer data on exposure to racism would be a key asset for future research in this area^28^. Second, there is evidence that educational attainment recorded on death certificates tends to overestimate the decedent’s level of education^29^. Lastly, because information on the social vulnerability-related factors was not available on the death certificates, we assigned the social vulnerability-related factors to all individual deaths within a county. It is important to note that contextual factors such as the social vulnerability index we used may not capture the individual-level socioeconomic status. Yet, contextual socioeconomic vulnerability may be a more relevant indicator to inform/guide targeted policies across the US counties to reduce air pollution related health disparities^30^. That being said, it would be interesting to explore multi-level socioeconomic disparities in relation to deaths attributable to air pollution in future work as such data becomes available.

## Methods

We harmonised mortality counts, population counts and PM_2.5_ concentration estimates to estimate PM_2.5_-attributable mortality across various sociodemographic population subgroups in the United States between 1990 and 2016. The population subgroups used in our analyses were racial/ethnic groups, educational attainment groups, rurality levels, socioeconomic status, household characteristics, racial and ethnic minority status, housing type and transportation, and social vulnerability index. We derived population counts at the census tract level from the U.S. Census Bureau, using linear interpolation to account for missing data in intercensal years. We acquired restricted-use county-level mortality counts from the U.S. National Vital Statistics System. Both data sets were pre-disaggregated by racial/ethnic groups and educational attainment at their respective sources. Additionally, we mapped levels of rurality, socioeconomic status, household characteristics, racial and ethnic minority status, housing type and transportation levels, and the social vulnerability index to the population counts and mortality counts based on county level look-up tables from the National Center for Health Statistics and the Centers for Disease Control and Prevention.

We mapped PM_2.5_ concentration levels from an established model^31^ and ground-based measurement data^32^ to census tracts. We assigned population-weighted mean PM_2.5_ exposure estimates to each county. To estimate the mortality burden that is attributable to PM_2.5_ at the county (and, subsequently, state and national) level, we combined our annual population-weighted mean PM_2.5_ exposure estimate and mortality counts at the county level with a CRF. The main CRF used in this analysis was by Di et al.^15^. For state or national-level analyses, we aggregated county-level all-cause and PM2.5-attributable mortality counts to the respective state or national level. We converted these raw mortality counts into age-adjusted mortality rates.

A detailed description of the data sources and methods can be found in the supplement.

## Data Availability

Data and code are publicly accessible on GitHub (https://github.com/FridljDa/pm25_inequality) and Zenodo (doi:10.5281/zenodo.10038691). Population estimates from the ACS and NCHS are publicly available and shared on the repositories above. Death certificate data was obtained from the National Center for Health Statistics, which mandates that all cells with fewer than 10 deaths and at the subnational level must be suppressed. Data derived from death certificates are, thus, only shared at the national level.

doi:10.5281/zenodo.10038691

https://github.com/FridljDa/pm25_inequality

## Acknowledgments

We thank Gabriel Carrasco-Escobar (University of California San Diego) for his help with the spatial analyses. We thank anonymous reviewers for helpful comments.

## Author contributions

Conceptualization: PG

Methodology: PG, DF, MVK, EB, SHN, MB, AT, TB

Investigation: DF

Software/ Formal analysis: DF

Visualization: PG, DF, MVK, AT, TB

Funding acquisition: PG

Project administration: PG, TB

Supervision: PG, TB

Writing – original draft: PG, DF, TB

Writing – review & editing: PG, DF, MVK, EB, SHN, MB, AT, TB

## Data and materials availability

Data and code are publicly accessible on GitHub (https://github.com/FridljDa/pm25_inequality) and Zenodo^33^. Population estimates from the ACS and NCHS are publicly available and shared on the repositories above. Death certificate data was obtained from the National Center for Health Statistics, which mandates that all cells with fewer than 10 deaths and at the subnational level must be suppressed. Data derived from death certificates are, thus, only shared at the national level.

## Materials and Methods

### Supplementary Text S1

#### List of Figures

Fig. S 1. Diagram summarizing the analysis steps to estimate age-adjusted PM2.5-attributable mortality rates.

Fig. S 2. Time trend in the population-weighted annual mean PM2.5 exposure for the age group 25+ years.

Fig. S 3. Time trend in the age-adjusted all-cause mortality rate for the age group 25+ years.

Fig. S 4. Population-weighted annual mean PM2.5 exposure in μg/m3 by sub population for the age group 25+ years.

Fig. S 5. Age-adjusted mortality per 100,000 from all causes by subpopulation for the age group 25+ years.

Fig. S 6. Age-adjusted mortality per 100,000 from all causes and population-weighted annual mean PM2.5 exposure for the population aged 25+ years in US states and the District of Columbia.

Fig. S 7. County level map of population-weighted mean PM2.5 exposure levels by racial/ethnic group in 2016.

Fig. S 8 Age-adjusted mortality per 100,000 attributable to PM2.5 exposure by subpopulation for the age group 25+ years.

Fig. S 9. Percent at which the age-adjusted PM2.5-attributable mortality rate was lower for each racial/ethnic group relative to Black Americans for the age group 25+ years.

Fig. S 10. Age-adjusted PM2.5-attributable mortality rate and all-cause mortality rate for each racial/ethnic group stratified by the social vulnerability index.

Fig. S 11. Age-adjusted PM2.5-attributable mortality rate and all-cause mortality rate for each racial/ethnic group stratified by socioeconomic status.

Fig. S 12. Age-adjusted PM2.5-attributable mortality rate and all-cause mortality rate for each racial/ethnic group stratified by household characteristics.

Fig. S 13. Age-adjusted PM2.5-attributable mortality rate and all-cause mortality rate for each racial/ethnic group stratified by minority status.

Fig. S 14. Age-adjusted PM2.5-attributable mortality rate and all-cause mortality rate for each racial/ethnic group stratified by Housing Type and Transportation.

Fig. S 15. Age-adjusted PM2.5-attributable mortality rate for the age group 25+ years in the USA.

Fig. S 16. Coefficient of Variation (CoV) of age-adjusted all-cause mortality attributable to PM2.5 by racial/ethnic group for different sociodemographic groups for the age group 25+ years.

Fig. S 17. Relative difference in PM2.5-attributable mortality rate for each racial/ethnic group between different sociodemographic groups.

Fig. S 18. Coefficient of Variation (CoV) of age-adjusted all-cause mortality attributable to PM2.5 by educational attainment for different sociodemographic groups for the age group 25+ years.

Fig. S 19. Age-adjusted PM2.5-attributable mortality rate and all-cause mortality rate for each education level stratified by socioeconomic status.

Fig. S 20. Age-adjusted PM2.5-attributable mortality rate and all-cause mortality rate for each education level stratified by rurality.

Fig. S 21. Age-adjusted PM2.5-attributable mortality rate and all-cause mortality rate for each education level stratified by social vulnerability index.

Fig. S 22. Age-adjusted PM2.5-attributable mortality rate and all-cause mortality rate for each education level stratified by household characteristic.

Fig. S 23. Age-adjusted PM2.5-attributable mortality rate and all-cause mortality rate for each education level stratified by minority status.

Fig. S 24. Age-adjusted PM2.5-attributable mortality rate and all-cause mortality rate for each education level stratified by housing type and transportation.

Fig. S 25. Relative difference in PM2.5-attributable mortality rate for each educational attainment group between different sociodemographic groups.

Fig. S 26. Coefficient of Variation (CoV) of age-adjusted all-cause mortality attributable to PM2.5 when stratifying by different sociodemographic groups for the age group 25+ years.

Fig. S 27. Age-adjusted PM2.5-attributable mortality rate for the age group 25+ years in the year 2016, by state and racial/ethnic group.

Fig. S 28. Age-adjusted mortality per 100,000 from all causes and attributable to PM2.5 (among those aged 25+ years) in US states and the District of Columbia in 2016 for the racial/ethnic groups “Black American” and “NH White”, low and high educational attainment, and rurality levels “Non-metro” and “large metro”.

Fig. S 29. County-level scatter plot of the age-adjusted PM2.5-attributable mortality per 100,000 for selected racial/ethnic groups and education levels.

Fig. S 30. County-level spatial clusters with high (red colors) and low (blue colors) differences in the age-adjusted PM2.5-attributable mortality rate between racial/ethnic groups.

Fig. S 31. Age-adjusted PM2.5-attributable mortality rate by racial/ethnic group, education level, and rurality level for the age group 65+.

Fig. S 32. Percentage of the age-adjusted all-cause mortality rate that was attributable to Fig. S 33. Differences in the age-adjusted PM2.5-attributable mortality rate between the racial/ethnic groups at the county level for the period 2000 to 2016 for the age group 65+ years.

Fig. S 34. Population-weighted annual mean PM2.5 exposure in μg/m3 by subpopulation for the age group 65+ years.

Fig. S 35. Age-adjusted mortality per 100,000 from all causes by racial/ethnic group, education attainment level, and rurality level for the age-group 65+ years.

Fig. S 36. Age-adjusted mortality per 100,000 from all causes and population-weighted average PM2.5 exposure for the population aged 65+ years in US states and District of Columbia.

Fig. S 37. Percent at which the PM2.5-attributable mortality rate per 100,000 is lower for each racial/ethnic group relative to Black Americans for the age group 65+ years.

Fig. S 38. Age-adjusted PM2.5-attributable mortality rate for the age group 65+ years in the USA.

Fig. S 39. Coefficient of Variation (CoV) of age-adjusted all-cause mortality attributable to PM2.5 when stratifying by different sociodemographic groups for the age group 65+ years.

Fig. S 40. Age-adjusted PM2.5-attributable mortality rate for the age group 65+ years in the year 2016, by state and racial/ethnic group.

Fig. S 41. Age-adjusted mortality per 100,000 from all causes and attributable to PM2.5 (among those aged 65+ years) in US states in 2016 for the racial/ethnic groups “Black American” and “NH White”, low and high educational attainment, and rurality levels “Non-metro” and “large metro”

Fig. S 42. County-level scatter plot of the age-adjusted mortality per 100,000 attributable to PM2.5 for selected racial/ethnic groups and education level comparisons for age group 65+ years.

Fig. S 43. County-level spatial clusters with high (red colors) and low (blue colors) differences in the age-adjusted PM2.5-attributable mortality rate between racial/ethnic groups for the age group 65+ years.

Fig. S 44. Analyses to examine the importance of using race-ethnicity specific CRFs for the age group 65+ years.

Fig. S 45. Differences in the proportion of all-cause mortality that was attributable to PM2.5 for the age group 65+ years between Black Americans and other racial/ethnic groups.

Fig. S 46. Exploring the link between PM2.5-attributable mortality at the county level and the variation of the PM2.5 estimates aggregated from the census tract to county level.

Fig. S 47. Shape of the CRFs by Di et al. (“Di”), the Global Burden of Disease project (“GBD”), and Burnett et al. (“GEMM”)

Fig. S 48. Percent of the difference in the age-adjusted mortality rate between each race-ethnicity and “Black American” that is attributable to PM2.5, using either race-ethnicity-specific CRFs (solid lines) or a uniform CRF (dashed lines).

Fig. S 49. Extent to which the difference in the age-adjusted mortality rate between each racial/ethnic group and Black Americans can be attributed to PM2.5.

Fig. S 50. Analyses to examine the importance of using race-ethnicity specific CRFs for the age group 25+ years.

Fig. S 51. Differences in the proportion of all-cause mortality that was attributable to PM2.5 for the age group 25+ years between Black Americans and other racial/ethnic groups.

Fig. S 52. Histogram of the number of census tract per county for the period 2000 to 2016.

Fig. S 53. Association of the variation in census-tract level PM2.5 measurements with county-level PM2.5-attributable mortality for the period 2000 to 2016 stratified by quantiles of the number of census tracts per county.

Fig. S 54. Percentage of all-cause mortality attributable to PM2.5 for each racial/ethnic group stratified by rurality.

Fig. S 55. Percentage of all-cause mortality attributable to PM2.5 for each racial/ethnic group stratified by the social vulnerability index.

Fig. S 56. Percentage of all-cause mortality attributable to PM2.5 for each racial/ethnic group stratified by socioeconomic status.

Fig. S 57. Percentage of all-cause mortality attributable to PM2.5 for each racial/ethnic group stratified by household characteristics.

Fig. S 58. Percentage of all-cause mortality attributable to PM2.5 for each racial/ethnic group stratified by minority status.

Fig. S 59. Percentage of all-cause mortality attributable to PM2.5 for each racial/ethnic group stratified by housing type & transportation.

Fig. S 60. Percentage of all-cause mortality attributable to PM2.5 for each educational attainment level stratified by rurality.

Fig. S 61. Percentage of all-cause mortality attributable to PM2.5 for each educational attainment level stratified by the social vulnerability index.

Fig. S 62. Percentage of all-cause mortality attributable to PM2.5 for each educational attainment level stratified by Socioeconomic Status.

Fig. S 63. Percentage of all-cause mortality attributable to PM2.5 for each educational attainment level stratified by Household Characteristic.

Fig. S 64. Percentage of all-cause mortality attributable to PM2.5 for each educational attainment level stratified by Minority Status.

Fig. S 65. Percentage of all-cause mortality attributable to PM2.5 for each educational attainment level stratified by Housing Type & Transportation.

## List of Tables

Table S 1. Data sources for the population counts used in the calculation of age-adjusted mortality rates.

Table S 2. Data sources for the population counts used in the estimation of population-weighted mean PM2.5 exposure.

Table S 3. ICD-10 codes used in each concentration-response function.

## Materials and Methods

This analysis brought together data from several sources. Fig. S 1 summarizes the different input data along with the geographic level at which this data was available, how we aggregated these data to the county level in order to generate a geographically harmonized dataset, and how the data were combined to arrive at age-adjusted PM_2.5_-attributable mortality rates for each population subgroup. In this Materials and Methods section, we provide a detailed description of the following steps of our analysis, with a separate subheading for each step: i) our definition of each racial/ethnic, education, rurality, and social vulnerability-related group; ii) each input variable used to calculate age-standardized mortality rates; iii) each input variable used to calculate population-weighted PM_2.5_ exposure; iv) the steps to geographically harmonize our data; v) the calculations to estimate age-adjusted PM_2.5_-attributable mortality; vi) the calculations to estimate the degree to which differences in all-cause mortality between racial/ethnic groups can be attributed to PM_2.5_; vii) a coefficient of variation approach used to compare the degree of disparity in PM_2.5_-attributable mortality across racial/ethnic groups versus education and rurality groups; viii) how we generated each map in our main manuscript and Supplementary Materials; and ix) our analyses to investigate the importance of using race-ethnicity-specific concentration-response functions (CRFs) when studying differences in PM_2.5_-attributable mortality between racial/ethnic groups. The studied time period was 1990-2016. We used R software, version 4.1.1 for all analyses (https://github.com/FridljDa/pm25_inequality).

### Defining the population subgroups

The population subgroups used in our analyses were racial/ethnic groups, education groups, and rurality levels, socioeconomic status, household characteristics, racial and ethnic minority status, housing type and transportation, and the social vulnerability index. We describe below how we defined each of these groups in our data.

#### Racial/ethnic group

The racial/ethnic groups considered in this study were: “White” (85.9% of study population in 1990), “Black American” (10.7% in 1990, 12.7% in 2016), “Asian or Pacific Islander” (2.8% in 1990, 6.3% in 2016), and “American Indian or Alaska Native” (0.7% in 1990, 1.2% in 2016). From 2000 onwards, the data from the Decennial Census allowed us to additionally disaggregate “White” into the racial/ethnic groups “Non-Hispanic (NH) White” (66.3% in 2016) and “Hispanic White” (13.4% in 2016).

#### Educational attainment level

We used educational attainment definitions from the US Census Bureau. Educational attainment was recorded into the following three categories: “low”, defined as high school graduate or lower (44.7% of study population in 2009, 40.6% in 2016); “medium”, defined as some college education, but no 4-year college degree (27.7% in 2009, 29.1% in 2016); and “high”, defined as 4-year college graduate or higher (27.5% in 2009, 30.3% in 2016)^35^. We chose these broad education categories because they minimize potential misclassification bias resulting from minor discrepancies in education reporting across the data sources for population and mortality counts^36^. The US Census Bureau has not published data on educational attainment for the racial/ethnic group “American Indian or Alaska Native” by age^35^. The racial/ethnic group “American Indian or Alaska Native” is, thus, not shown in Figure 3 in the main manuscript.

#### Rurality level

We used the 2013 version of the Urban Rural Classification Scheme from the National Center for Health Statistics to assign a rurality level to each county in our dataset^37^. To achieve improved clarity of our findings, we aggregated six rurality levels in the scheme into three categories. Specifically, we aggregated “large fringe metro” and “large central metro” into “large metro” (56.6% of study population in 2000, 54.8% in 2016), “small metro” and “medium metro” into “small-medium metro” (29.2% in 2000, 30% in 2016), and “non-core” and “micropolitan” into “non metro” (14.2% in 2000, 15.2% in 2016). We did not assign a rurality level for deaths for which the county of residence and occurrence of death were not available (0% of deaths in 1990 and 0.0008% in 2016).

#### Socioeconomic Status

We assigned socioeconomic status (SES) to each county in our dataset using estimates from the CDC’s Agency for Toxic Substances and Disease Registry^38^. These estimates integrate metrics such as the population percentage living below 150% of the poverty line, unemployment rates, housing costs, educational levels, and health insurance coverage. The agency ranks counties on a percentile scale for SES. We categorized these rankings into ’Low SES’ (0-33^rd^ percentile), ’Middle SES’ (34-66th percentile), and ’High SES’ (67-100th percentile). In our study, ’High SES’ signifies better resource and opportunity access than ’Low SES’.

#### Household Characteristics and Disability

We classified counties using the CDC’s Agency for Toxic Substances and Disease Registry’s estimates of Household Characteristics^38^. These estimates consider various demographic factors: population percentages of those over 65 or under 18, civilians with disabilities, single-parent households, and individuals with limited English proficiency. Each county received a percentile rank for Household Characteristics, which we grouped into three categories: ’Low HC’ (0-33rd percentile), ’Middle HC’ (34-66th percentile), and ’High HC’ (67-100th percentile). In our study, a ’High HC’ designation signals greater household stability and demographic benefits compared to ’Low HC’ counties.

#### Racial and Ethnic Minority Status

We categorized counties using the CDC’s Agency for Toxic Substances and Disease Registry estimates for Racial and Ethnic Minority Status^38^. These estimates reflect the composition of various racial and ethnic groups, excluding Hispanic or Latino: Black or African American, American Indian and Alaska Native, Asian, Native Hawaiian and Other Pacific Islander, Two or More Races, and Other Races. Counties received a percentile rank reflecting their Minority Status. We then organized these ranks into three levels: ’Low MS’ for counties in the 0-33rd percentile, ’Middle MS’ for the 34-66th percentile, and ’High MS’ for the 67-100th percentile. In our study, a ’High MS’ level suggests a lower prevalence of racial and ethnic minorities, while a ’Low MS’ level indicates a higher prevalence.

#### Housing Type and Transportation

We categorized each county in our dataset using the CDC’s Agency for Toxic Substances and Disease Registry estimates for housing and transportation factors^38^. These factors include the presence of multi-unit structures, mobile homes, crowding (more than one person per room), lack of vehicle access, and group quarters such as dormitories and nursing homes. The agency provides a percentile ranking for each county based on these criteria. We classified these rankings into three levels: ’Low HS’ for counties in the 0-33rd percentile, indicating more challenging housing and transportation conditions; ’Middle HS’ for those in the 34-66th percentile; and ’High HS’ for counties in the 67-100th percentile, which suggests better housing and transportation conditions.

#### Social Vulnerability Index

We applied the CDC’s Agency for Toxic Substances and Disease Registry’s Social Vulnerability Index (SVI) to classify counties in our dataset^38^. The SVI evaluates a community’s resilience by examining socioeconomic status, household composition, minority status, and housing types. Each county was ranked by percentile for SVI. We grouped these rankings into: ’Vulnerable SVI’ (0-33rd percentile), indicating higher susceptibility; ’Moderate SVI’ (34-66^th^ percentile); and ’Resilient SVI’ (67-100th percentile), suggesting lower vulnerability to events like natural disasters or epidemics.

### Data sources to calculate age-standardized mortality rates

This study analyzed de-identified, restricted-use mortality counts from the US National Vital Statistics System covering all deaths occurring within the United States^39^. For both of these datasets, every entry is a record of a deceased person taken from death certificates that were submitted to the vital statistics departments of every US state and the District of Columbia. For the years 1990-1998, the cause of death was coded according to the International Classification of Diseases (ICD) Ninth Revision. For the years 1999-2016, the cause of death was coded according to the International Classification of Diseases (ICD) Tenth Revision. We assigned each death to the county of residence of the deceased individual. When the county of residence was missing but the address where the death occurred was present (which was the case in 0.16% of cases in 1990 and 0% in 2016), we used the county of death instead. We limited our analysis to deaths occurring in the age group 25 years and older, because this is the most common age group analyzed in studies on the health effects of long-term PM_2.5_ exposure^9,16^, and because some of the population counts (see “Data sources to calculate population-weighted PM_2.5_ exposure”) were only available for this age group^35^.

We ignored deaths for which the age at the time of death was not available. This was the case for 0.005% of deaths in 2016. Furthermore, we ignored deaths for which the decedent’s racial/ethnic group or education was unknown or did not fall in the pre-specified categories. This amounted to 0.027% of deaths in 1990 and 0.004% of deaths in 2016 for the racial/ethnic group, and 4.87% of deaths in 2009 and 1.72% of deaths in 2016 for education. After these exclusions, a total of 63 261 546 deaths between 1990 and 2016 (2 057 450 deaths in 1990 and 2 678 827 deaths in 2016) remained in the analysis. The finest geographic resolution available for the mortality counts was county.

### Data sources to calculate population-weighted PM_2.5_ exposure

#### PM_2.5_ exposure

We compiled annual mean estimates of PM_2.5_ concentration on a 0.01° by 0.01° (0.9 km by 1.1 km) grid from an established model for the 48 states of the contiguous USA and the District of Columbia^31^. The model combines chemical transport modeling, satellite remote sensing, and ground-based measurements. Meng et al. have reported that the estimated PM_2.5_ level from this model was generally consistent with direct ground-based PM_2.5_ measurements, with a cross-validation R^2^ between 0.6 and 0.85^31^. For Alaska and Hawaii, which were not included in the model by Meng et al.^31^, we collected ground-based measurement data from pre-generated annual summary files provided by the United States Air Quality System Data Mart^32^. In 2016, the monitors measuring PM_2.5_ in the state of Alaska were located in Anchorage, Fairbanks, the City and Borough of Juneau, and the Matanuska-Susitna Borough, and in the state of Hawaii in Honolulu, Kauai, and Maui. Averaged over the years 1990-2016, the population weighted-median distance between a census tract centroid and a monitoring site in Alaska and Hawaii was 14 km and 8 km, respectively.

#### Population counts for estimation of population-weighted mean PM_2.5_ exposure

A detailed list of the data sources used for population counts in the estimation of population-weighted mean PM_2.5_ exposure can be found in table S2. To estimate population-weighted mean PM_2.5_ exposure for each population subgroup, we used population counts at the lowest geographic level available, which was the census tract. Most census tracts in the USA have a population size between 1200 and 8000 (86.7% of census tracts in 1990; 93.5% in 2016). Population counts by racial/ethnic group and age group at the census tract level are from the US Decennial Census for the years 1990, 2000, and 2010^40–42^, and from the 5-year estimates in the American Community Survey for the years 2009, and 2011-2016^35^. The American Community Survey data were provided by the US Census Bureau after imputing missing information.

Population estimates for the periods 1991-1999 and 2001-2008 were obtained by linearly interpolating the data from the 1990 and 2000 Decennial Census for the period 1991 to 1999, and 2000 and 2010 Decennial Census for the period 2001 to 2008. Because the boundaries of census-defined geographies are adjusted for every Decennial Census in response to changes in population, we first used the Longitudinal Tract Data Base^43^ to crosswalk all Decennial Census to 2010 vintage census tract boundaries. This method is well validated^44^. The data from the Decennial Census and the interpolations were aggregated by 5-year age groups (with the highest category being age 85+ years).

Between 2009 and 2016, the American Community Survey offered population estimates at the census-tract level, categorized by age group and highest educational attainment^35^. We restricted counts by educational level to ages 25 and above, as persons under age 25 may not have completed their education. For the period 2009-2016, the American Community Survey also provides population estimates for all permutations of racial/ethnic group and education level at the census-tract level for the age group 25 years and above^35^.

### Geographic harmonization

We aggregated our PM_2.5_ estimates, originally at a 0.01° by 0.01° (0.9 km by 1.1 km) grid resolution, to the county level to match the resolution of our mortality counts. Fig. S 1 summarizes this process.

We assigned annual PM_2.5_ concentration estimates to each census tract by mapping the nearest PM_2.5_ estimate from a 0.01° by 0.01° (0.9 km by 1.1 km) grid to the centroid of the respective tract. To aggregate PM_2.5_ exposure data from census tracts to counties, we utilized population estimates for each racial/ethnic, educational, rural, and social-vulnerability subgroup at the census tract level (refer to Table S2). This allowed us to calculate the population-weighted mean PM_2.5_ exposure annually for each subgroup at the county level. Figure S3 shows the population-weighted mean PM_2.5_ exposure, calculated using this process, at the national level for each population subgroup.

### Estimation of the age-adjusted PM_2.5_-attributable mortality rate

To estimate county-level mortality burden attributable to PM_2.5_, we combined our annual population-weighted mean PM_2.5_ exposure estimates at the county level with mortality counts at the county level, using a concentration-response function (CRF) within a health impact assessment framework. A CRF 𝑓 is a function from a concentration level 𝑧 taking values in [0,1] and, in this analysis, depends on the population subgroup 𝑐. It is used to calculate the response, the PM_2.5_-attributable mortality count at the county level, in the following form: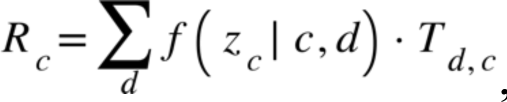, where is the population-weighted mean PM_2.5_ exposure at the county level, 𝑑 is a disease outcome (see table S3), is the mortality count for the subpopulation 𝑐 and disease 𝑑 at the county level, and 𝑅_*c*_ is the PM_2.5_-attributable mortality count for subpopulation 𝑐 at the county level. For analyses at the state or national level, the county-level all-cause and PM_2.5_-attributable mortality counts were then summed to the state or national level. We converted these raw mortality counts into an age-adjusted mortality rate using the direct method^1,45^. The reference population was the 2000 US standard population as used in the National Vital Statistics Reports, Vol. 57, No. 14^1^. The age categories in the US standard population were 25-34, 35-44, 45-54, 55-64, 65-74, 75-84, and 85+ years. In data sources that provided population counts for more granular age categories, we thus aggregated the counts to these coarser age categories. Table S1 lists, by geographic level, population subgroup, and year, each data source and age categories that were used for the calculation of age-adjusted mortality rates.

As explained in the main text, the CRF used in this analysis was by Di et al.^15^. Di et al. provide both a general CRF that does not distinguish by racial/ethnic group as well as CRFs specific to each racial/ethnic group considered in this study. We applied racial/ethnic group-specific CRFs only in analyses where we disaggregated results by these groups. The CRF 𝑓 from Di et al.^15^ is a continuous and stepwise linear function. Specifically, 𝑓 is zero for all values of annual mean PM_2.5_ between 0 and 5 μg/m^3^ and has a positive slope 𝑚_*c*_ (which varies by racial/ethnic group) for all annual mean PM_2.5_ levels greater than 5 μg/m^3^. Figure Fig. **S 47** illustrates the shape of the CRF. Di et al.^15^ provide the mean estimate along with a 95% CI for each 𝑚_*c*_.

The CRF by Di et al. was developed among US adults aged 65 years and older^15^, whereas our analysis applies this CRF to those aged 25 years and older. To examine whether restricting our study population to those aged 65 years and older would change any of our conclusions, we additionally conducted all analyses shown in the main manuscript when restricting the population to this older age group. The results from these analyses are shown in the Supplementary Materials (Figures Fig Fig. **S 31** - Fig. **S 45**).

### Estimation of the proportion of the all-cause mortality that is attributable to PM_2.5_

To quantify the role of PM_2.5_ attributable mortality relative to the all-cause mortality, we calculated the ratio between both. Specifically, we calculated the proportion of all-cause mortality that is attributable to PM_2.5_ using the formula:

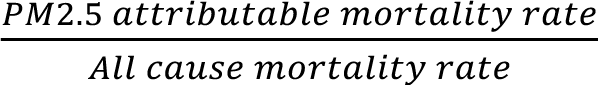

The percentage of all-cause mortality attributable to PM_2.5_ is obtained by multiplying this proportion by 100.

### Estimation of the proportion of the all-cause mortality difference to a given racial/ethnic group that is attributable to PM_2.5_

To quantify the role of PM_2.5_ in mortality disparities between population subgroups, we calculated the ratio between the difference in PM_2.5_-attributable mortality and the difference in all-cause mortality with respect to a fixed reference group. Specifically, we used the following formula:

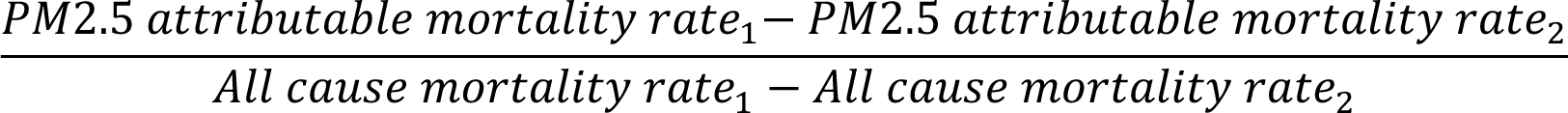

For the racial/ethnic groups, we chose “Black American” as reference group because this racial/ethnic group had both the highest PM_2.5_-attributable as well as all-cause mortality rate.

### Comparing the degree of disparity in PM_2.5_-attributable mortality

To compare the degree to which the age-adjusted PM_2.5_-attributable mortality rate varies for different population subgroup, we adopted the coefficient of variation (CoV) approach from Jbailey et al.^46^. To do so, we first computed the age-adjusted PM_2.5_-attributable mortality rate (𝑎) for each population subgroup. We then calculated:

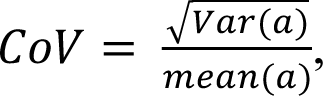

where 𝑉𝑎𝑟(𝑎) is the empirical variance of 𝑎 and 𝑚𝑒𝑎𝑛(𝑎) is the empirical mean of 𝑎. For example, consider the racial/ethnic groups “Black American”, “NH White”, “White Hispanic or Latino”, “Asian or Pacific Islander”, and “American Indian and Alaska Native”, where the age-adjusted mortality per 100,000 attributable to PM_2.5_ are, respectively, 𝑎 = (600, 400, 500, 400, 400). Then 𝑉𝑎𝑟(𝑎) = 8000, 𝑚𝑒𝑎𝑛(𝑎) = 460 and thus 𝐶𝑜𝑉 ≈ 0.19. Coefficient of variation, therefore, is a measure of the variability of the data normalized by its magnitude^15^.

### Spatial analyses

We first mapped, for each racial/ethnic group, the percentage of the all-cause age-adjusted mortality rate that is attributable to PM_2.5_ by US state and District of Columbia in the year 2016. We then proceeded to map the county-level age-adjusted PM_2.5_-attributable mortality per 100,000 for the three largest racial/ethnic groups (NH White, Hispanic or Latino White, and Black Americans) because county-level sample sizes were insufficient for other racial/ethnic groups. We also mapped county-specific differences in the age-adjusted PM_2.5_-attributable mortality rate between i) Black American and the NH White population, and ii) Black American and the Hispanic or Latino White population. All county-level maps were created for each year of our study period (i.e., from 2000 to 2016) as well as for the (unweighted) mean across the years 2000 to 2016. Lastly, we generated all county-level maps when taking spatial autocorrelation into account. To do so, we calculated the local indicator of spatial association (LISA) using local Getis-Ord Gi statistics with contiguity-based spatial weights at the county level^47^.

### Analyses to assess the effect of using a race-ethnicity-specific CRF

We carried out three sets of analyses to investigate the degree to which the use of CRFs that ignore differences in the concentration-response association by racial/ethnic group underestimate disparities in PM_2.5_-attributable mortality between racial/ethnic groups.

First, we compared our findings when using the race-ethnicity-specific CRF by Di et al.^15^ to those obtained when using the uniform CRFs by Cohen et al.^16^ and Burnett et al.^11^. We selected these two CRFs because they are two recent and widely used CRFs for the US population. The shape of these CRFs is provided in figure Fig. **S 47**. Each of the CRFs used in this analysis are based on hazard ratio models that incorporate risk information from PM_2.5_ and provide 95% confidence intervals. While Di et al.^15^ provide hazard ratios for all-cause mortality, the CRFs by Cohen et al.^16^ and Burnett et al.^11^ are cause-specific. The ICD-10 codes used for these cause-specific functions are listed in table S3. Using our notation from the formula presented under “Estimation of the age-adjusted PM_2.5_-attributable mortality rate” above, we can represent the CRF by Cohen et al.^16^ in the following way. Cohen et al. provide 900 draws 𝑓_5_(𝑧|𝑐, 𝑑), …, 𝑓_;<<_(𝑧|𝑐, 𝑑) for the CRF 𝑓(𝑧|𝑐, 𝑑) for each age-group 𝑐 and disease outcome 𝑑 and exposure level 𝑧. For each draw 𝑖, we calculated 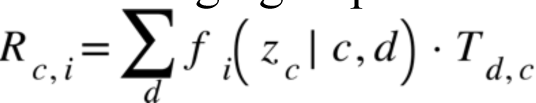. was then estimated by averaging out 𝑅_*c,i*_, i.e., 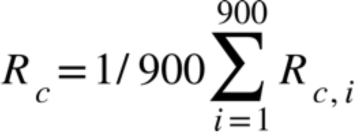. We estimated 95% confidence intervals for 𝑅_*c*_ by taking the 2.5^th^ and 97.5^th^ percentile from this distribution of 900 values of 𝑅_*c*_.

Confidence intervals for the sum of 𝑅_*c*_ across age groups were then obtained by assuming that each age group-specific 𝑅_*c*_follows an independent Gaussian distribution (thus allowing us to estimate the standard deviation of each 𝑅_*c*_from its confidence intervals). Burnett et al. ^11^(*12*) provide the mean estimate and standard deviation for 𝑓(𝑧|𝑐, 𝑑). 95% confidence interval bounds for 𝑓(𝑧|𝑐, 𝑑) were estimated by assuming a normal distribution around the mean for 𝑓(𝑧|𝑐, 𝑑) and, thus, using 1.96 times the standard deviation above and below the mean to arrive at the upper and lower 95% confidence interval bounds, respectively. Burnett et al.^11^ provide one main CRF 𝑓 for all non-communicable diseases. Thus, the formula for 𝑅_*c*_ reduces to 𝑅_*c*_ = 𝑓(𝑧_*c*_|𝑐, 𝑑) ⋅^𝑇^>.

Second, we compared our findings when using the race-ethnicity-specific CRF from Di et al.^15^ to those obtained when applying the CRF for “NH White” by Di et al. to all racial/ethnic groups. This comparator, thus, ignores that racial/ethnic groups in the USA have different susceptibilities to PM_2.5_ by assuming that all racial/ethnic groups have the same susceptibility to PM_2.5_ as the NH White population. We used the same coefficient of variation approach to compare the effect of this choice on the magnitude of differences in age-adjusted PM_2.5_-attributable mortality between racial/ethnic groups as described in these Supplementary Materials under “Comparing the degree of disparity in PM_2.5_-attributable mortality”.

Third, we used the observed mortality counts from 2016 and race-ethnicity-specific CRFs (again, from Di et al.^15^), but assumed the same PM_2.5_ exposure for all racial/ethnic groups by applying the population-weighted mean PM_2.5_ exposure in the overall USA population to all racial/ethnic groups. This approach accounts for varying susceptibility to PM_2.5_ among racial/ethnic groups while presuming uniform PM_2.5_ exposure across these groups.

### Comparison of CRF with race-ethnicity-specific estimates to CRFs that do not provide race-ethnicity-specific estimates

When using race-ethnicity-specific CRFs, we estimated that a large proportion of the differences in all-cause mortality between racial/ethnic groups can be attributed to PM_2.5_ exposure (Fig. 2, Fig. S 48). While we estimated that more than half of the difference in age-adjusted mortality between the NH White and Black American population was attributable to PM_2.5_ in each year from 2000 to 2011, the same proportion ranged only from 2.7% to 16.7% in each year of this period when using CRFs^16,18^ that do not provide race-ethnicity-specific estimates (Fig. S 48). To further investigate the importance of using race-ethnicity-specific CRFs when studying differences in PM_2.5_-attributable deaths between racial/ethnic groups, we conducted two additional analyses. First, we compared our findings to those obtained when applying the CRF by Di et al.^15^ for the NH White population to all racial/ethnic groups, thus assuming that all racial/ethnic groups have the same susceptibility to a given level of PM_2.5_ exposure. Implementing this stylized scenario leads to substantially lower variation by racial/ethnic group (Fig. S 48, Fig. S 49, Fig. S 50, Panel A), with only between 10.5% and 6.5% of the difference in mortality to Black Americans being attributable to PM_2.5_ in each year from 2000 to 2015. Second, we used the observed mortality counts from 2016 and race-ethnicity-specific CRFs, but assumed the same PM_2.5_ exposure for all racial/ethnic groups by applying the population-weighted average PM_2.5_ exposure in the overall USA population to all racial/ethnic groups. Underscoring the importance of taking into account differential susceptibility to PM_2.5_ exposure between racial/ethnic groups, this analysis (shown in Fig. S 50, Panel B) still leads to substantial disparities in PM_2.5_-attributable mortality between racial/ethnic groups, with 38.7 age-adjusted deaths per 100,000 for Black Americans, 14.1 for the Hispanic White population, 12.9 for American Indians or Alaska Natives, 10.4 for the NH White population, and 8.2 for Asian or Pacific Islanders at the overall population-weighted average PM_2.5_ exposure of 7.2µg/m^3^.

Similarly, we still observe substantial differences by racial/ethnic groups in PM_2.5_-attributable mortality when assuming that all counties achieve the PM_2.5_ targets set by the USA Environmental Protection Agency (12 μg/m^3^) or the World Health Organization (10 μg/m^3^) (Fig. S 50, Panel C).

### Uncertainty estimation

We derived the point estimate and confidence interval for Di et al.’s CRF from the original study^15^. In our analysis, we propagated these confidence intervals using the Delta Method and parametric bootstrapping. The Delta Method, under the assumption of Gaussian distributions, approximates the variance of a function of random variables primarily through first-order Taylor expansions. Parametric bootstrapping involves generating new sample datasets by resampling from a fitted model, and then re-estimating the parameter of interest on these new datasets to construct its empirical distribution and confidence intervals. The Delta Method was the preferred approach in cases where parametric bootstrapping was computationally prohibitive.

## Supplementary Text S1

### Selected text passages relevant to environmental justice from Executive Order 12898, Executive Order 14008, and the National Ambient Air Quality Standard program

#### Executive Order 12898 of February 11, 1994: (Federal Actions to Address Environmental Justice in Minority Populations and Low-Income Populations)

“Section 1–1. Implementation.

1–101. Agency Responsibilities. To the greatest extent practicable and permitted by law, and consistent with the principles set forth in the report on the National Performance Review, each Federal agency shall make achieving environmental justice part of its mission by identifying and addressing, as appropriate, disproportionately high and adverse human health or environmental effects of its programs, policies, and activities on minority populations and low-income populations in the United States and its territories and possessions, the District of Columbia, the Commonwealth of Puerto Rico, and the Commonwealth of the Mariana Islands.”

“Sec. 3–3. Research, Data Collection, and Analysis.

3–301. Human Health and Environmental Research and Analysis. (a) Environmental human health research, whenever practicable and appropriate, shall include diverse segments of the population in epidemiological and clinical studies, including segments at high risk from environmental hazards, such as minority populations, low-income populations and workers who may be exposed to substantial environmental hazards.”

“3–302. Human Health and Environmental Data Collection and Analysis. To the extent permitted by existing law, including the Privacy Act, as amended (5 U.S.C. section 552a): (a) each Federal agency, whenever practicable and appropriate, shall collect, maintain, and analyze information assessing and comparing environmental and human health risks borne by populations identified by race, national origin, or income. To the extent practical and appropriate, Federal agencies shall use this information to determine whether their programs, policies, and activities have disproportionately high and adverse human health or environmental effects on minority populations and low-income populations;”

Source:

Federal Register / Vol. 59, No. 32 / Wednesday, February 16, 1994 / Presidential Documents Clinton, William J. URL: https://www.archives.gov/files/federal-register/executive-orders/pdf/12898.pdf

#### Executive Order 14008 of January 27, 2021 (Tackling the Climate Crisis at Home and Abroad)

“Securing Environmental Justice and spurring Economic Opportunity:

Sec. 219. Policy. To secure an equitable economic future, the United States must ensure that environmental and economic justice are key considerations in how we govern. That means investing and building a clean energy economy that creates well-paying union jobs, turning disadvantaged communities—historically marginalized and overburdened—into healthy, thriving communities, and undertaking robust actions to mitigate climate change while preparing for the impacts of climate change across rural, urban, and Tribal areas. Agencies shall make achieving environmental justice part of their missions by developing programs, policies, and activities to address the disproportionately high and adverse human health, environmental, climate-related and other cumulative impacts on disadvantaged communities, as well as the accompanying economic challenges of such impacts. It is therefore the policy of my Administration to secure environmental justice and spur economic opportunity for disadvantaged communities that have been historically marginalized and overburdened by pollution and underinvestment in housing, transportation, water and wastewater infrastructure, and health care.” (Biden, J., page 7629).

Source:

Biden, J. “Executive Order 14008 of January 27, 2021: Tackling the Climate Crisis at Home and Abroad.” Federal Register 86.19 (2021): 7619-7633.

URL: https://www.regulations.gov/document/EPA-HQ-OPPT-2021-0202-0012

#### National Ambient Air Quality Standards for Particulate Matter; Final Rule of July 18, 1997

“SUMMARY: This document describes EPA’s decision to revise the national ambient air quality standards (NAAQS) for particulate matter (PM) based on its review of the available scientific evidence linking exposures to ambient PM to adverse health and welfare effects at levels allowed by the current PM standards. The current primary PM standards are revised in several respects: Two new PM_2.5_ standards are added, set at 15 μg/m^3^, based on the 3year average of annual arithmetic mean PM_2.5_ concentrations from single or multiple community-oriented monitors, and 65 μg/m^3^, based on the 3-year average of the 98th percentile of 24-hour PM_2.5_ concentrations at each population-oriented monitor within an area; and the current 24-hour PM10 standard is revised to be based on the 99th percentile of 24-hour PM10 concentrations at each monitor within an area. The new suite of primary standards will provide increased protection against a wide range of PMrelated health effects, including premature mortality and increased hospital admissions and emergency room visits, primarily in the elderly and individuals with cardiopulmonary disease; increased respiratory symptoms and disease, in children and individuals with cardiopulmonary disease such as asthma; decreased lung function, particularly in children and individuals with asthma; and alterations in lung tissue and structure and in respiratory tract defense mechanisms. The current secondary standards are revised by making them identical in all respects to the new suite of primary standards. The new secondary standards, in conjunction with a regional haze program, will provide appropriate protection against PM-related public welfare effects including soiling, material damage, and visibility impairment. In conjunction with the new PM_2.5_ standards, a new reference method has been specified for monitoring PM as PM_2.5_.” (Environmental Protection Agency, page 38652) “E. Environmental Justice Executive Order 12848 (58 FR 7629, February 11, 1994) requires that each Federal agency make achieving environmental justice part of its mission by identifying and addressing, as appropriate, disproportionately high and adverse human health or environmental effects of its programs, policies, and activities on minorities and low-income populations. These requirements have been addressed to the extent practicable in the RIA cited in this unit.” (Environmental Protection Agency, page 38708)

Source:

US Environmental Protection Agency. (1997). National ambient air quality standards for particulate matter: Final rule. Fed. Regist., 62(138), 38-651.

URL: https://archive.epa.gov/ttn/pm/web/pdf/pmnaaqs.pdf

**Fig. S 1.**
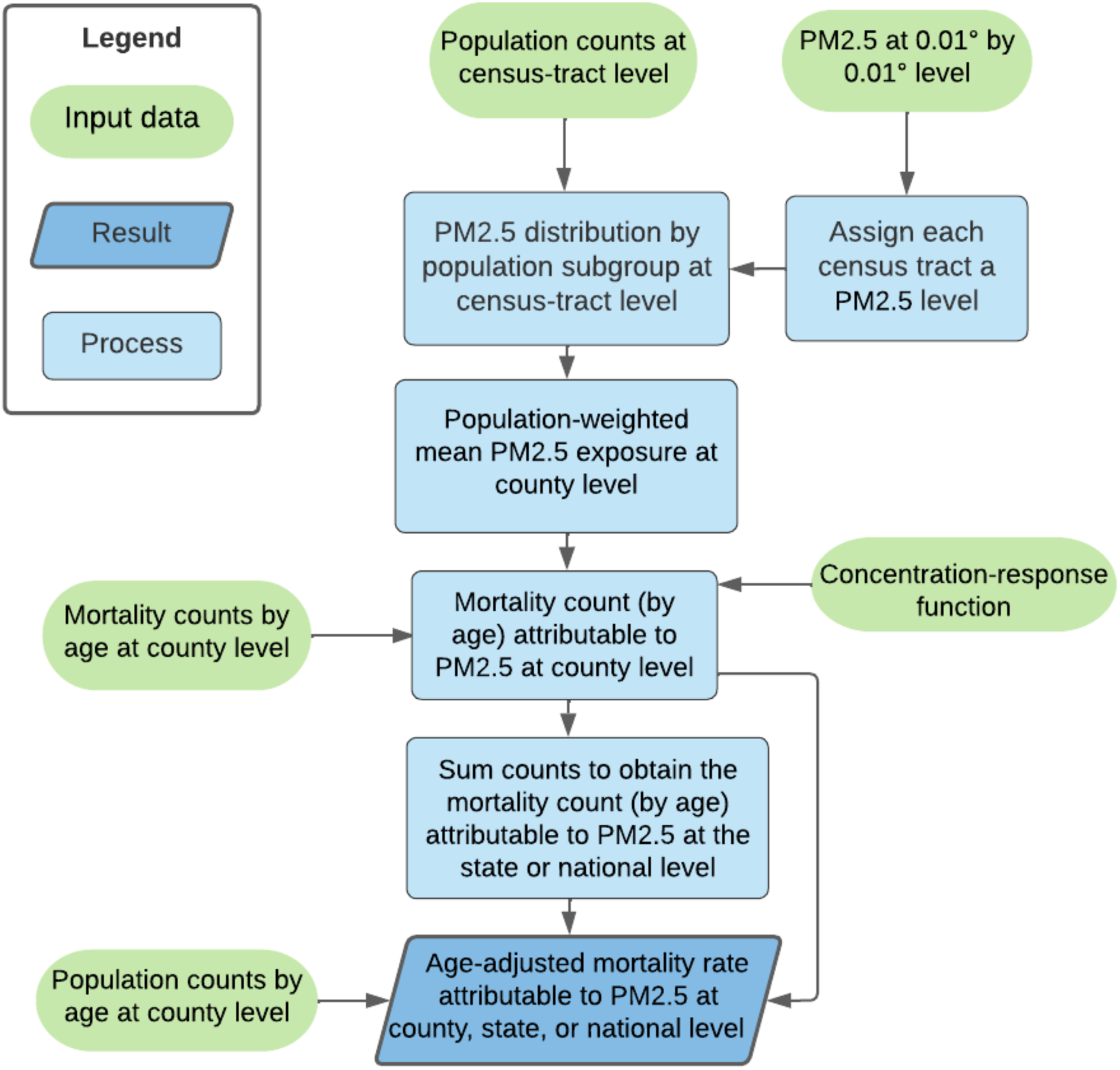
Diagram summarizing the analysis steps to estimate age-adjusted PM_2.5_-attributable mortality rates. All “Process” steps in the diagram were conducted separately for each year and subgroup in our study period. For example, we assigned the mean PM2.5 exposure level for an entire year to a given census tract.

**Fig. S 2.**
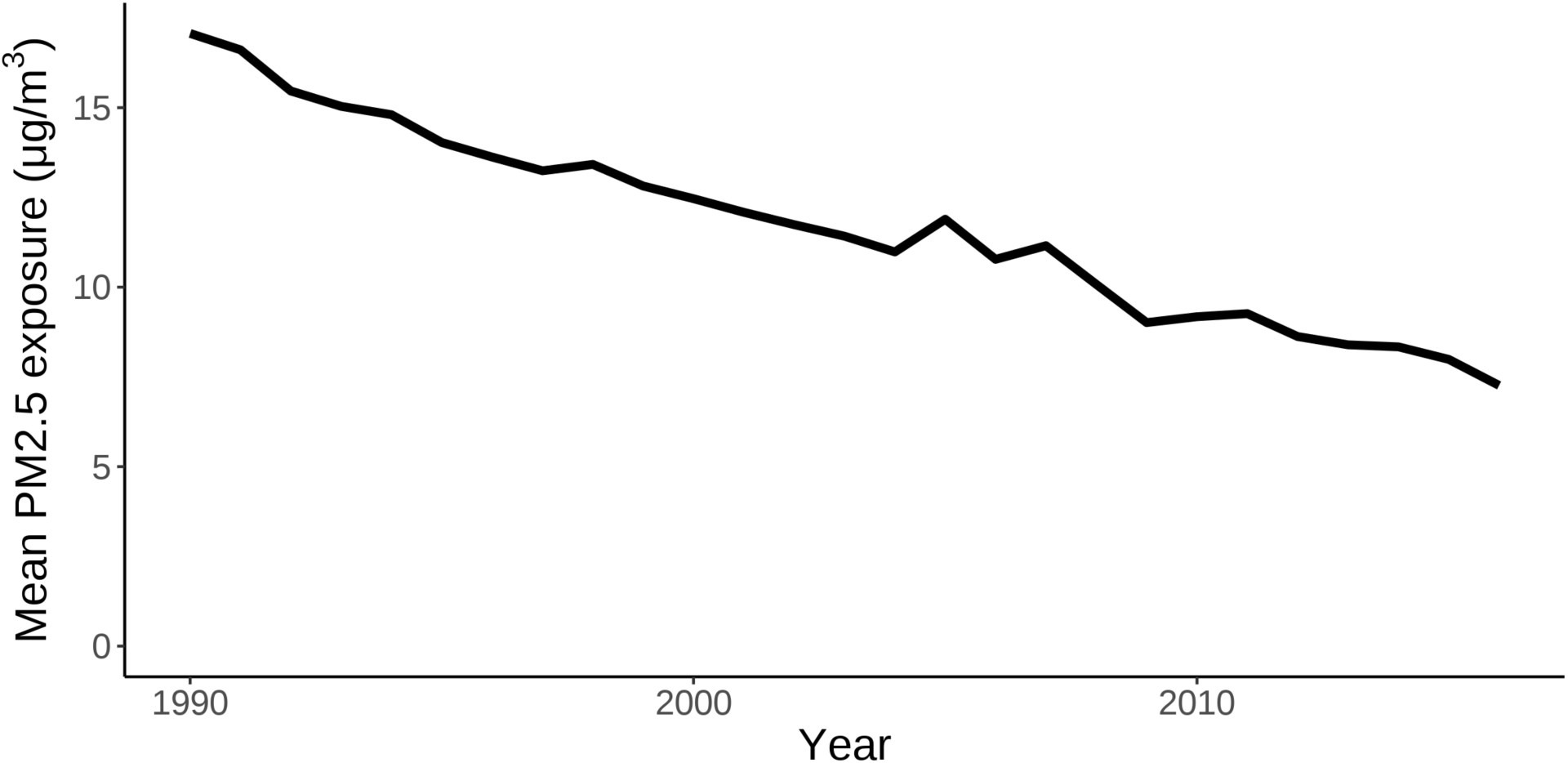
Time trend in the population-weighted annual mean PM_2.5_ exposure for the age group 25+ years.

**Fig. S 3.**
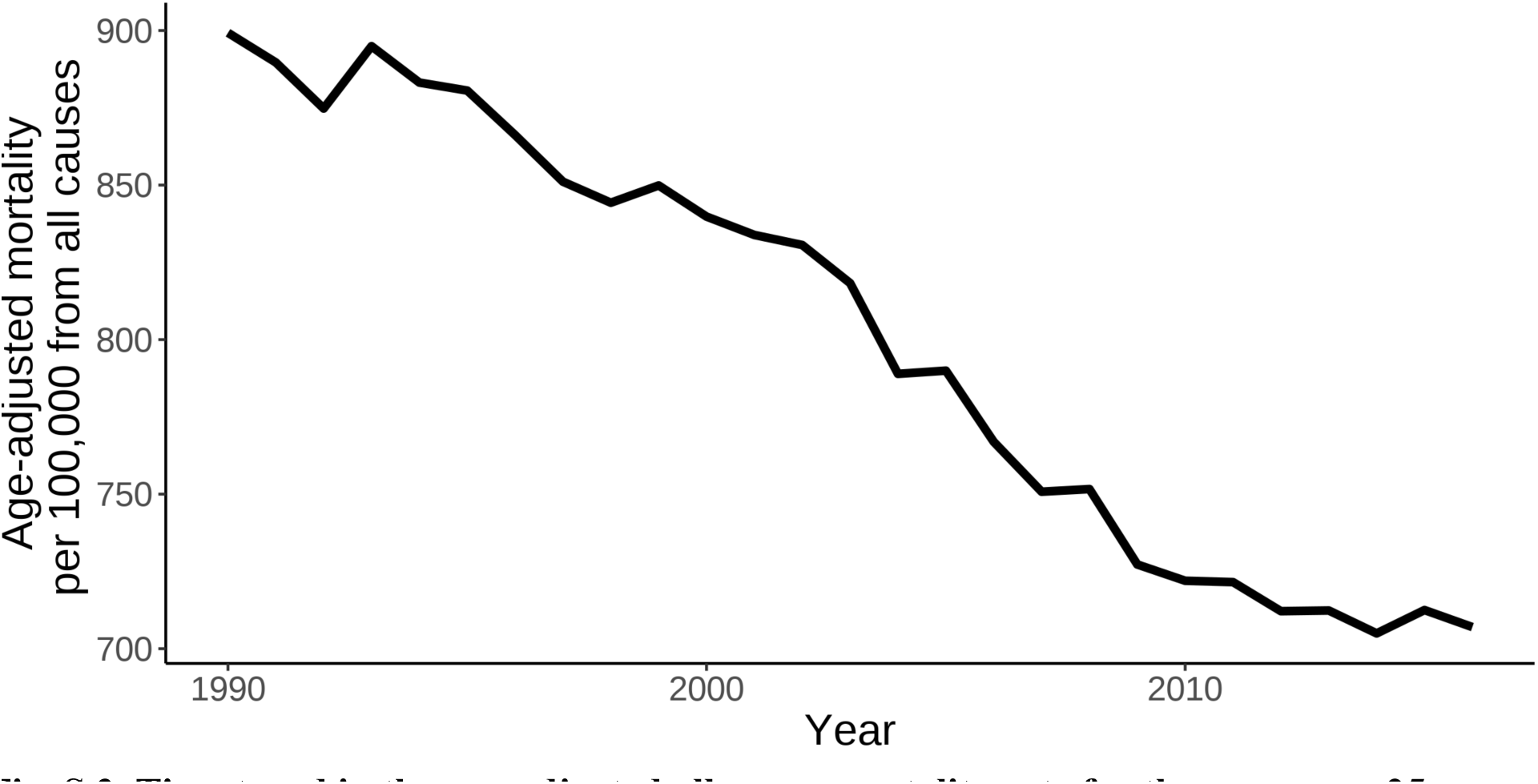
Time trend in the age-adjusted all-cause mortality rate for the age group 25+ years.

**Fig. S 4.**
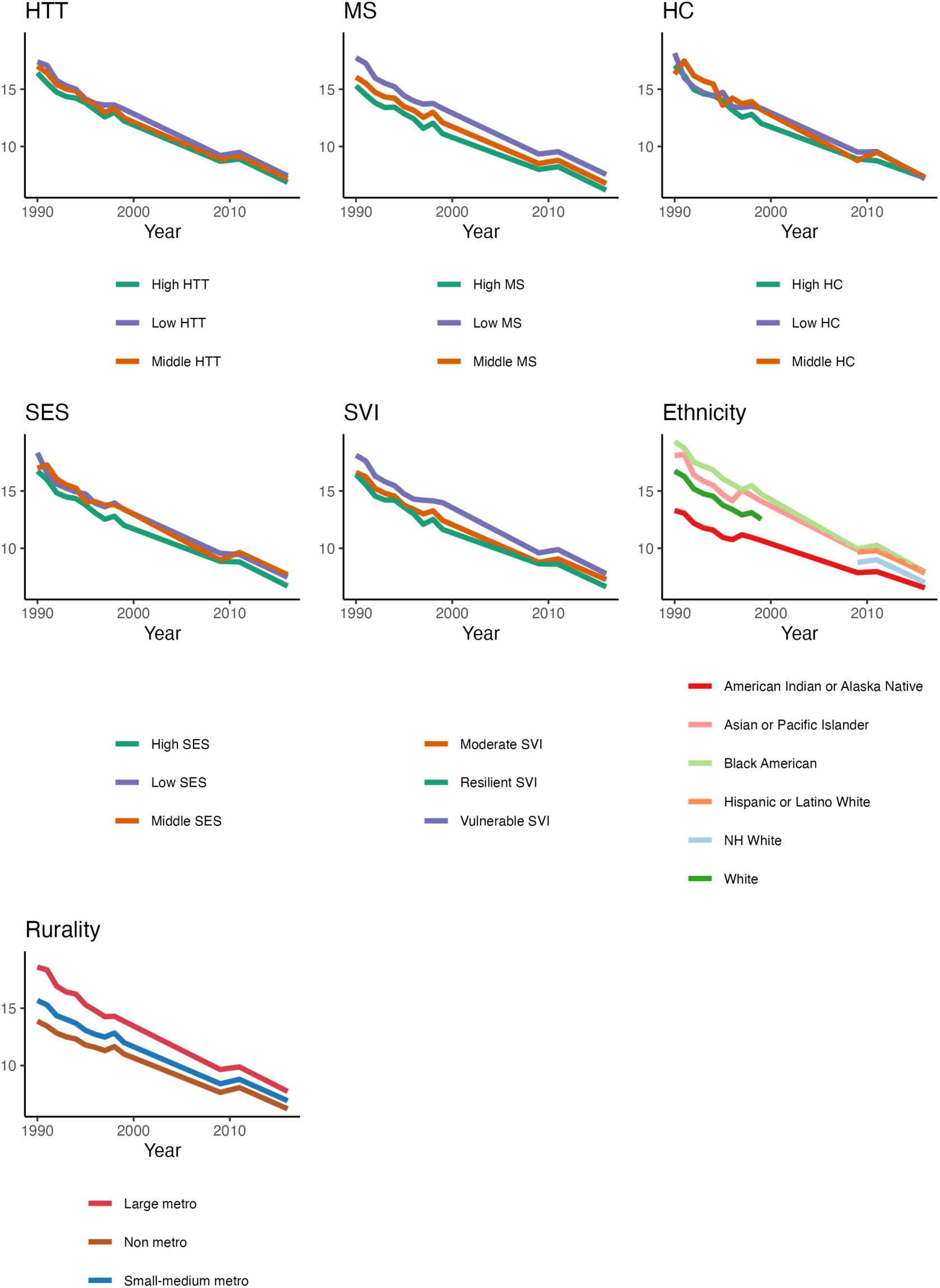
Population-weighted annual mean PM_2.5_ exposure in μg/m^3^ by sub population for the age group 25+ years. Abbreviations: SES = Socioeconomic Status, HC = Household Characteristics, MS = Minority Status, HTT = Housing Type & Transportation, SVI = Social Vulnerability Index, NH=Non-Hispanic.

**Fig. S 5.**
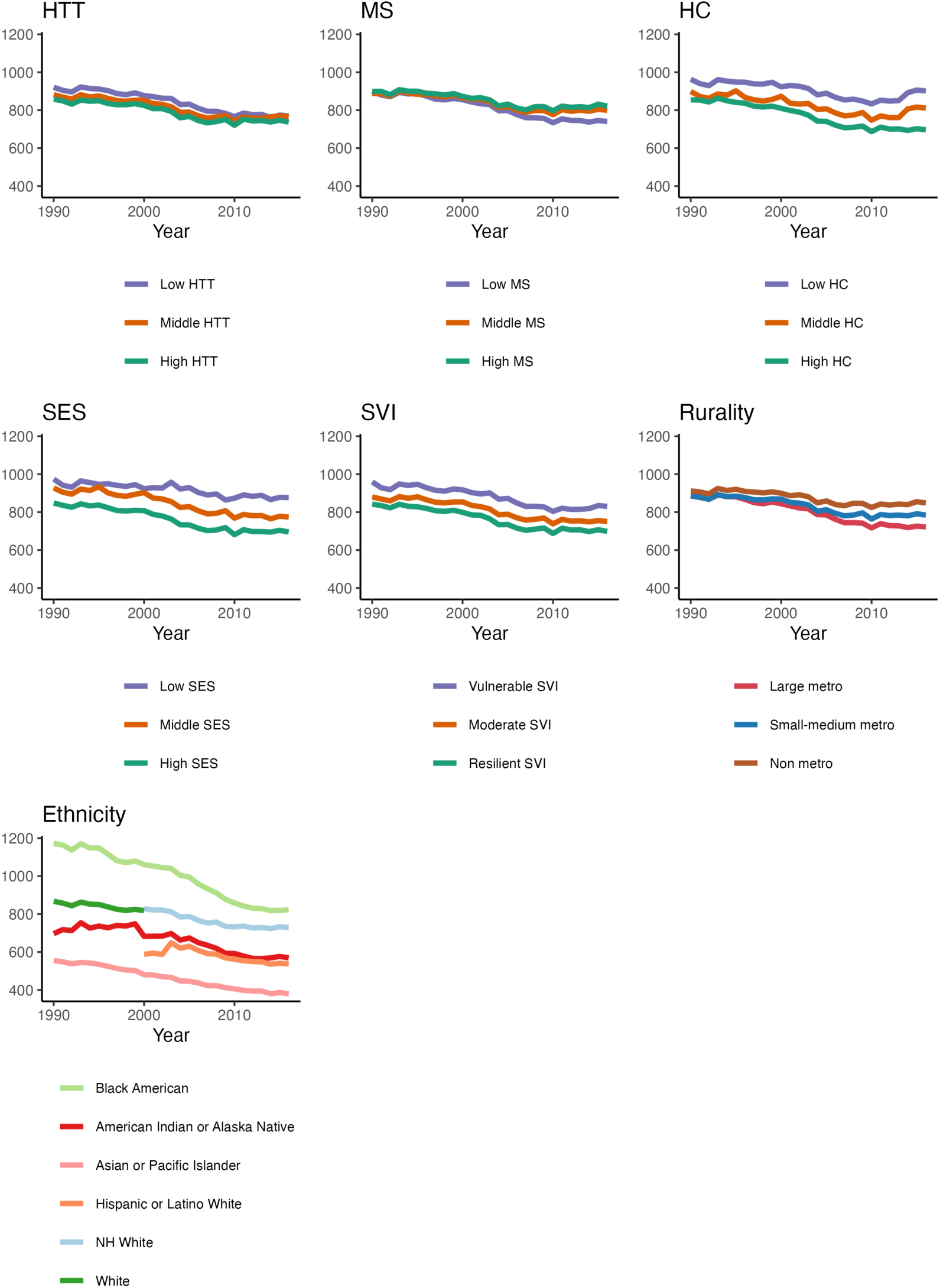
Age-adjusted mortality per 100,000 from all causes by subpopulation for the age group 25+ years. Abbreviations: SES = Socioeconomic Status, HC = Household Characteristics, MS = Minority Status, HTT = Housing Type & Transportation, SVI = Social Vulnerability Index, NH=Non-Hispanic.

**Fig. S 6.**
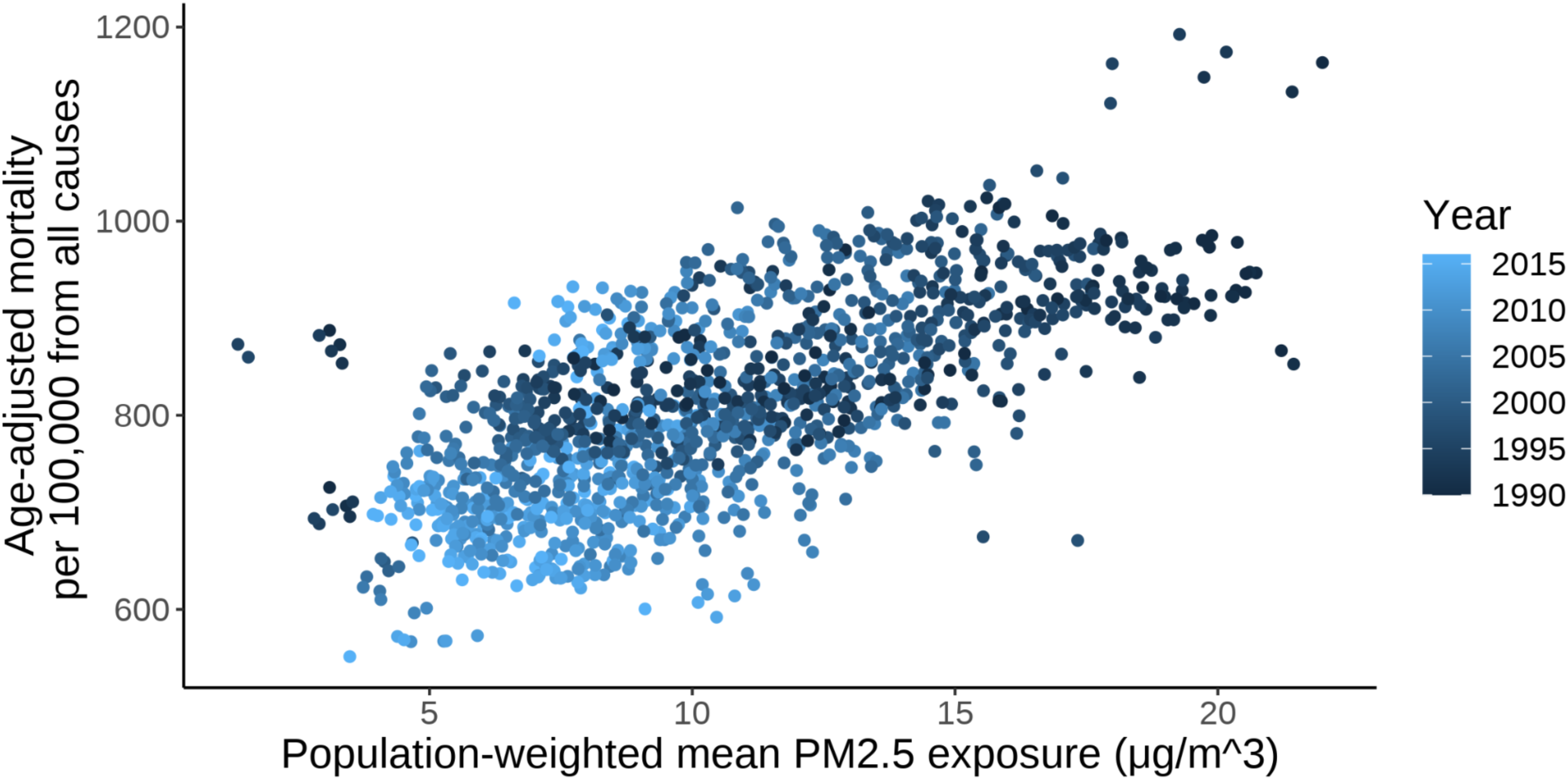
Age-adjusted mortality per 100,000 from all causes and population-weighted annual mean PM2.5 exposure for the population aged 25+ years in US states and the District of Columbia. Each dot corresponds to a year-state combination.

**Fig. S 7.**
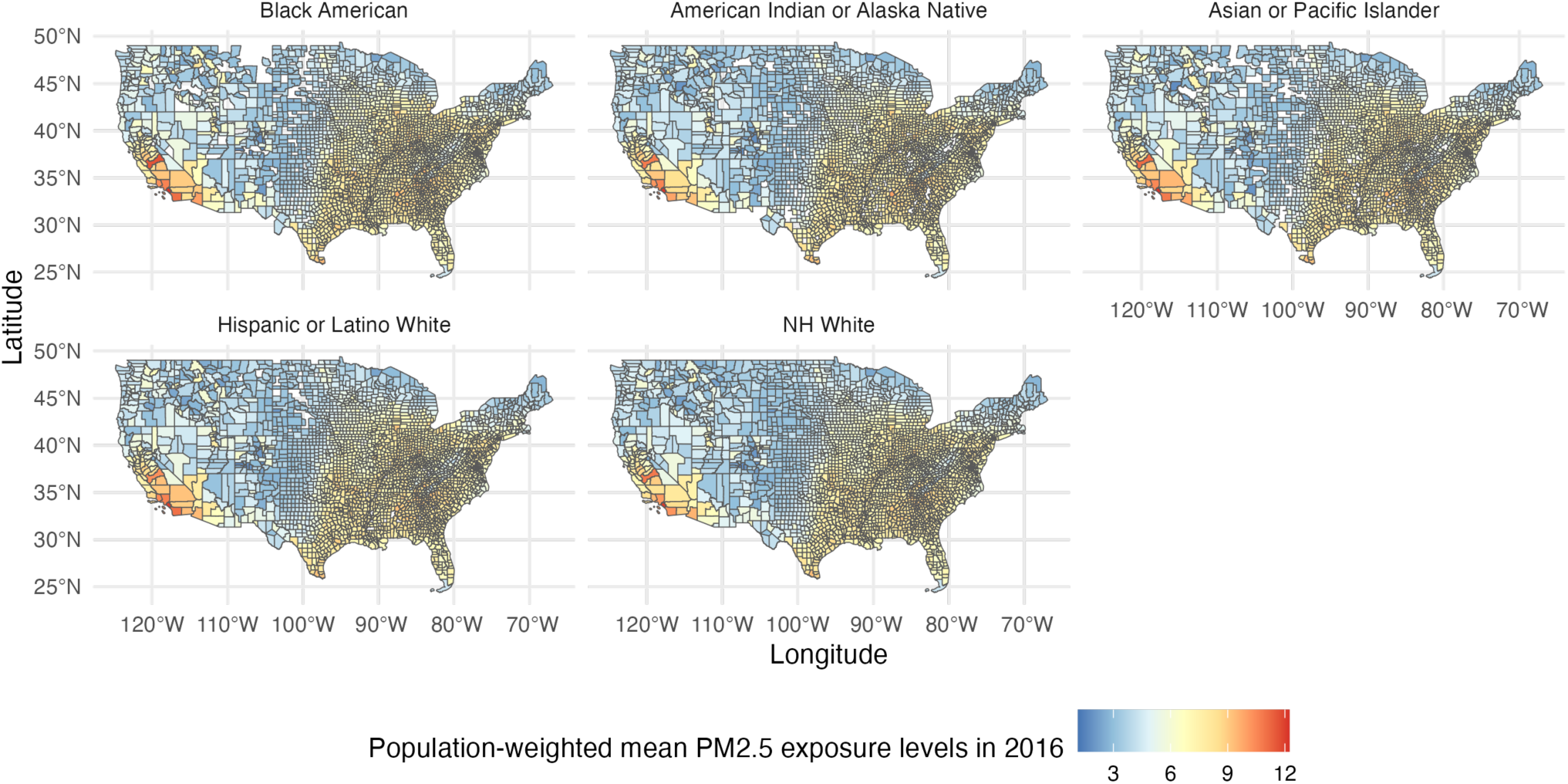
County level map of population-weighted mean PM2.5 exposure levels by racial/ethnic group in 2016. Abbreviations: NH=Non-Hispanic.

**Fig. S 8.**
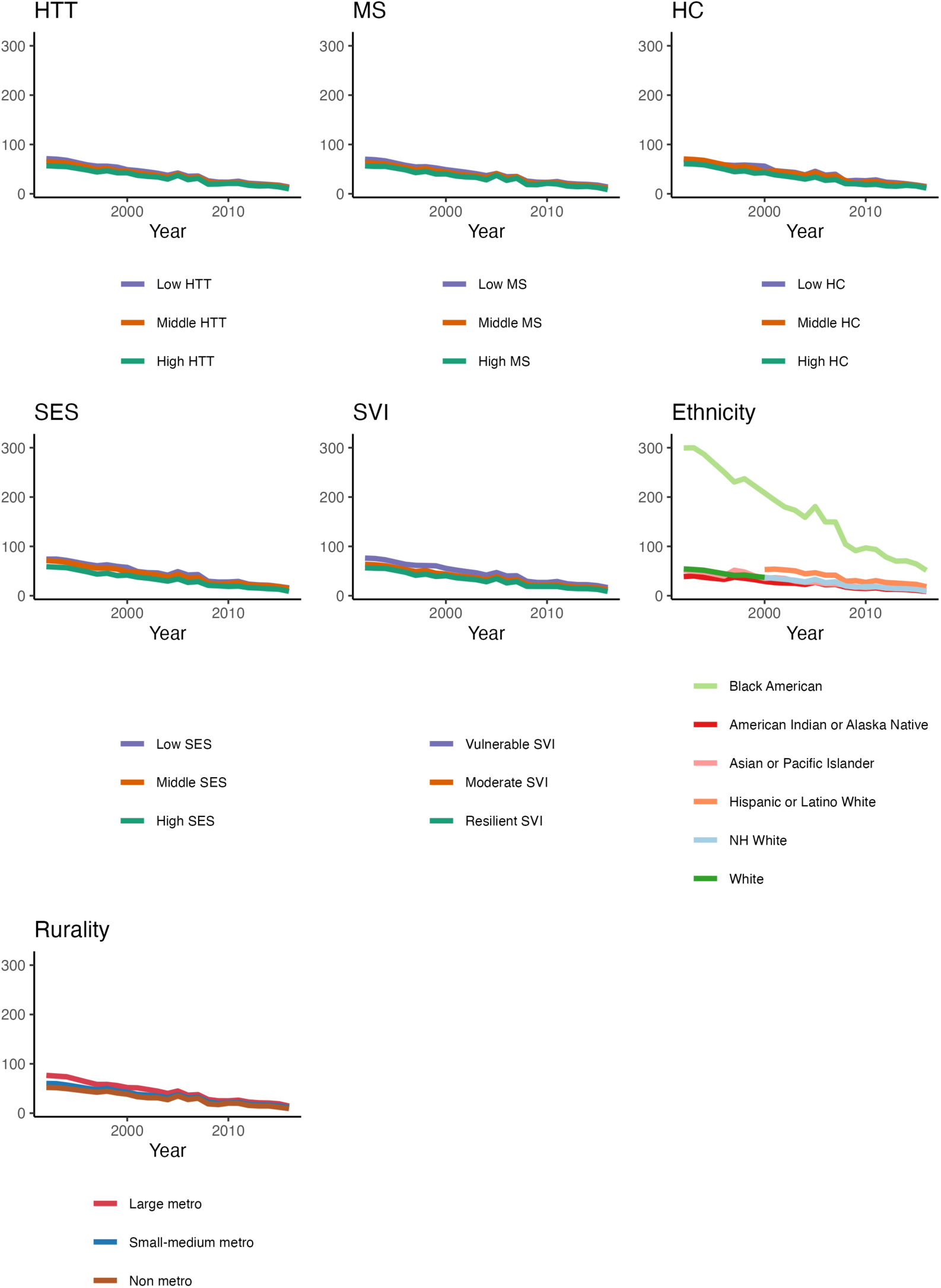
Age-adjusted mortality per 100,000 attributable to PM_2.5_ exposure by subpopulation for the age group 25+ years. Abbreviations: SES = Socioeconomic Status, HC = Household Characteristics, MS = Minority Status, HTT = Housing Type & Transportation, SVI = Social Vulnerability Index, NH=Non-Hispanic.

**Fig. S 9.**
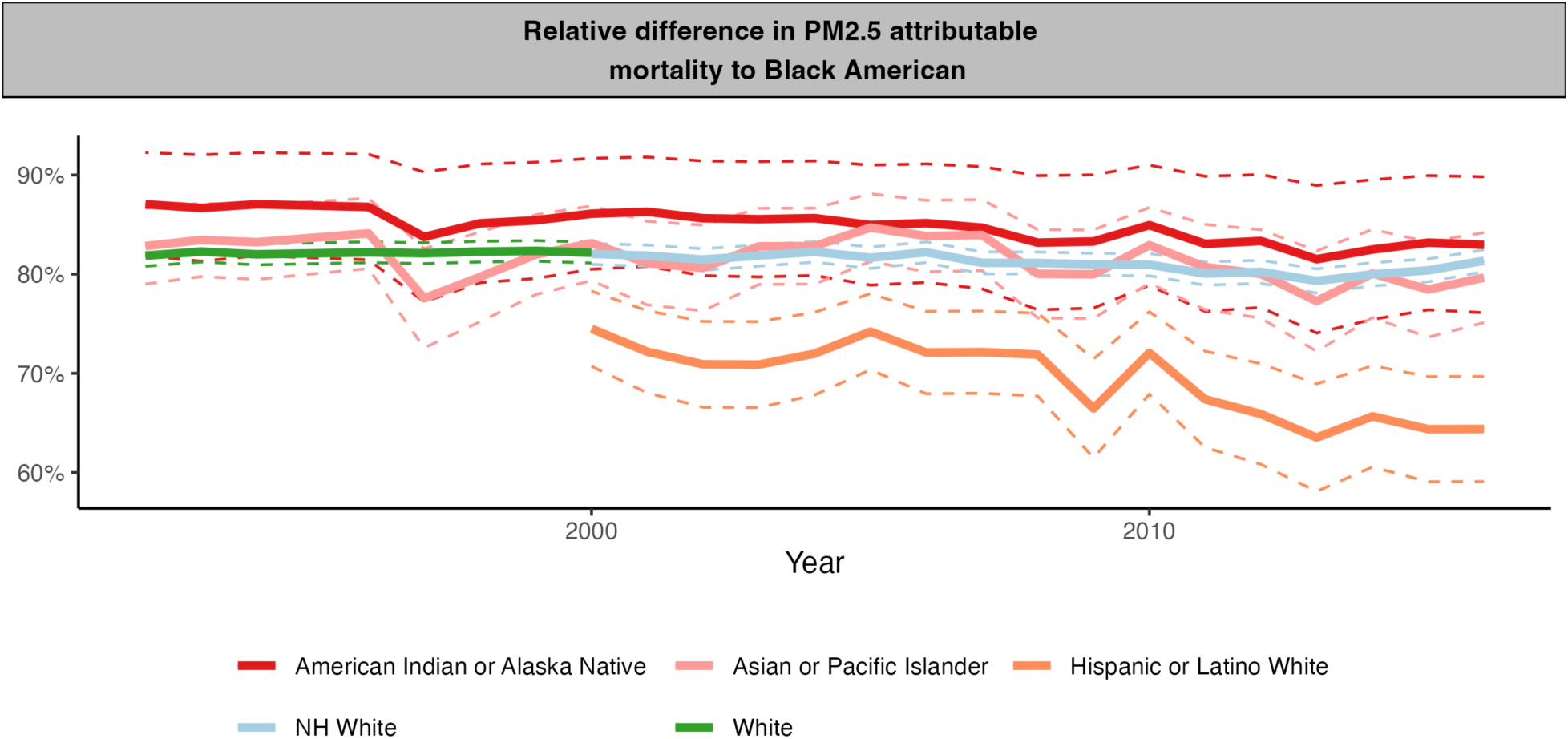
Percent at which the age-adjusted PM_2.5_-attributable mortality rate was lower for each racial/ethnic group relative to Black Americans for the age group 25+ years. This figure is for the age group 25+ years and spans the period 1990 to 2016. On average during the period 1990 to 2016, the PM2.5-attributable mortality rate for the “Asian or Pacific Islander” and “American Indian or Alaska Native” population was 81.8% and 85.3% lower, respectively, than for the “Black American” population. The PM2.5-attributable mortality rate for the “White (All Hispanic Origins)” population was 82.0% lower than for the Black American population during the period 1990 to 2000. Lastly, on average during the period 2000 to 2016, the PM2.5-attributable mortality rate for the “NH White” and “Hispanic or Latino White” population was 81.4% and 70.1% lower, respectively, than for the “Black American” population.

**Fig. S 10.**
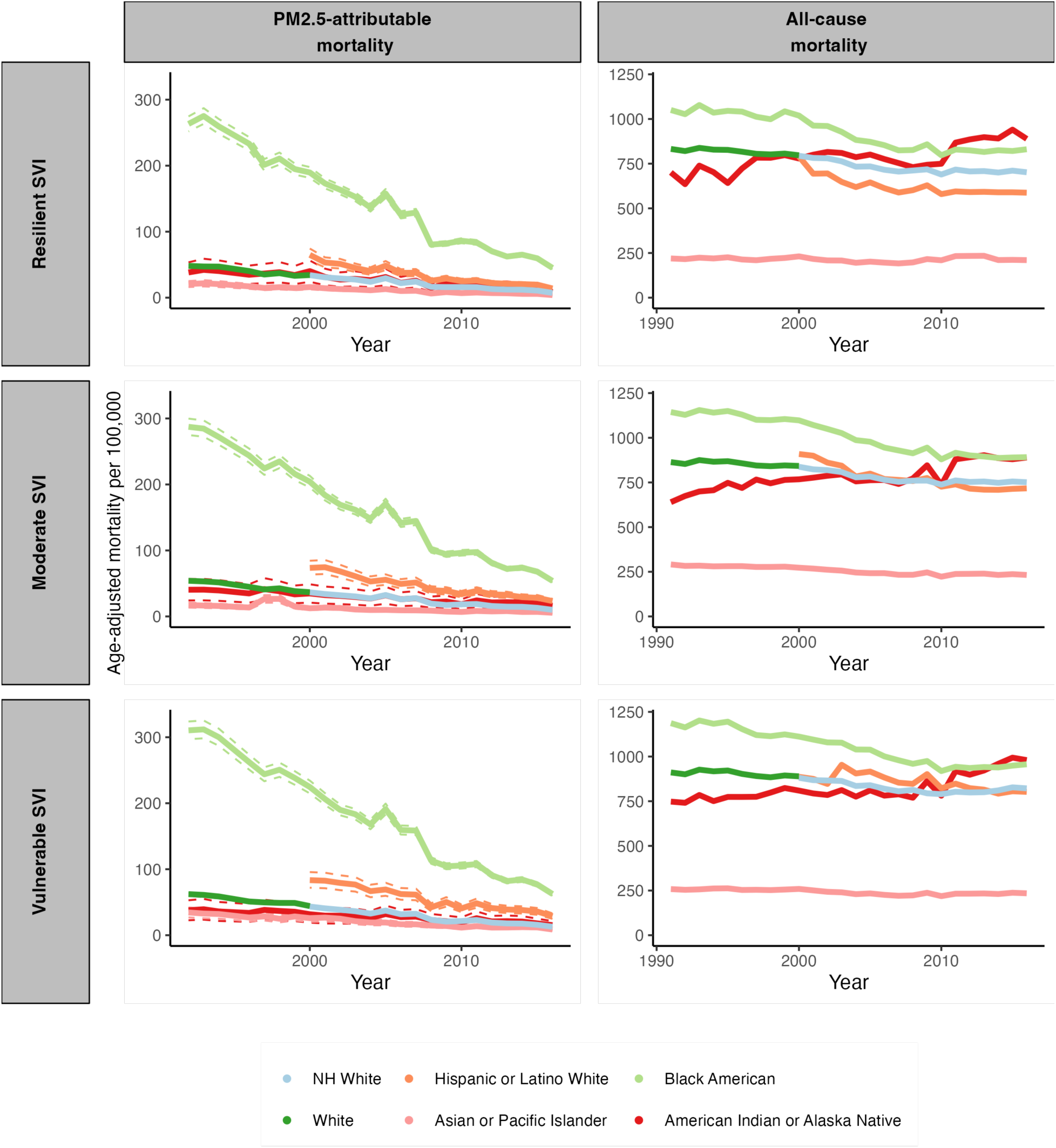
Age-adjusted PM_2.5_-attributable mortality rate and all-cause mortality rate for each racial/ethnic group stratified by the social vulnerability index. The dashed lines depict 95% confidence intervals. Abbreviations: NH=Non-Hispanic.

**Fig. S 11.**
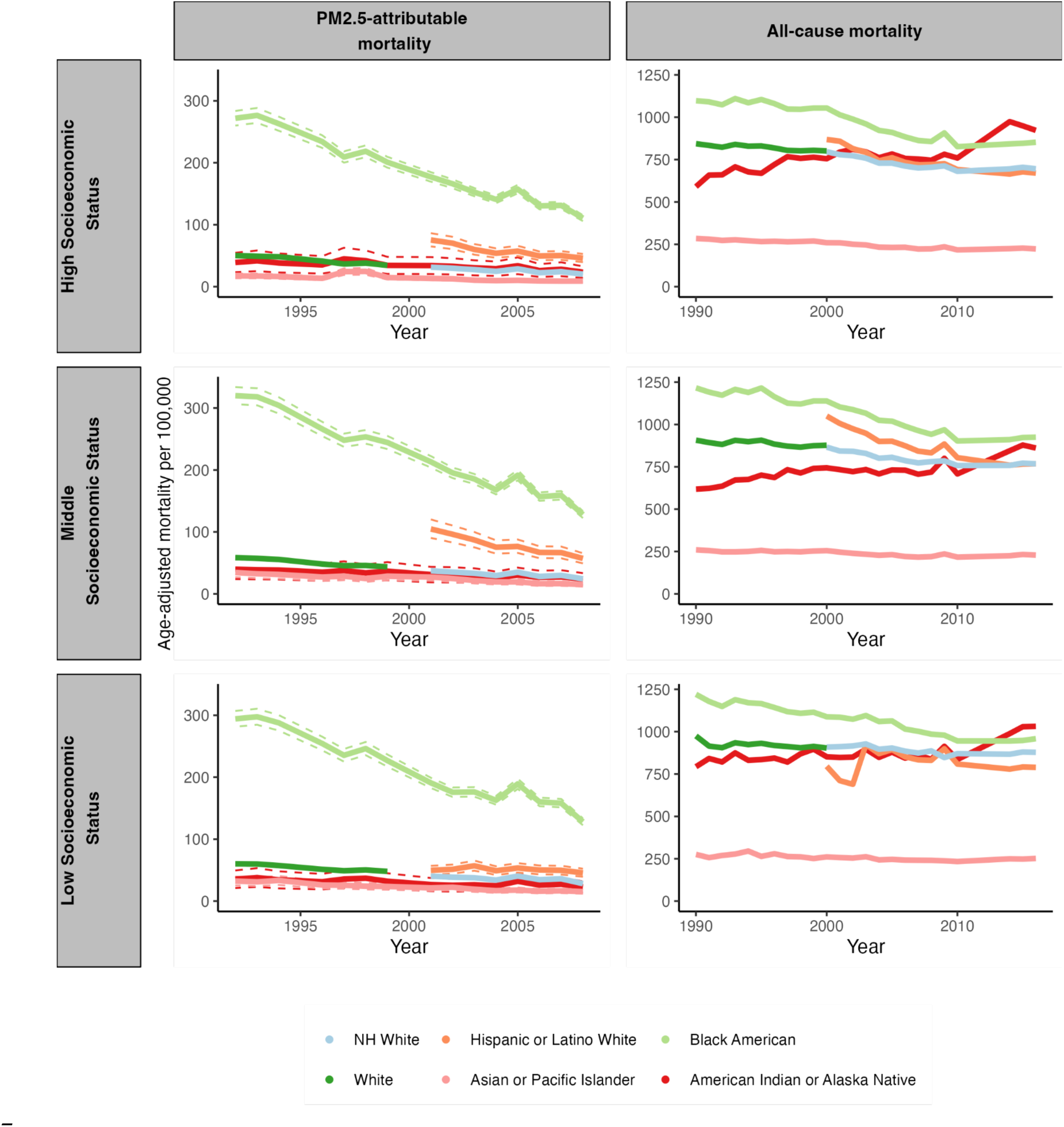
Age-adjusted PM_2.5_-attributable mortality rate and all-cause mortality rate for each racial/ethnic group stratified by socioeconomic status. The dashed lines depict 95% confidence intervals. Abbreviations: NH=Non-Hispanic.

**Fig. S 12.**
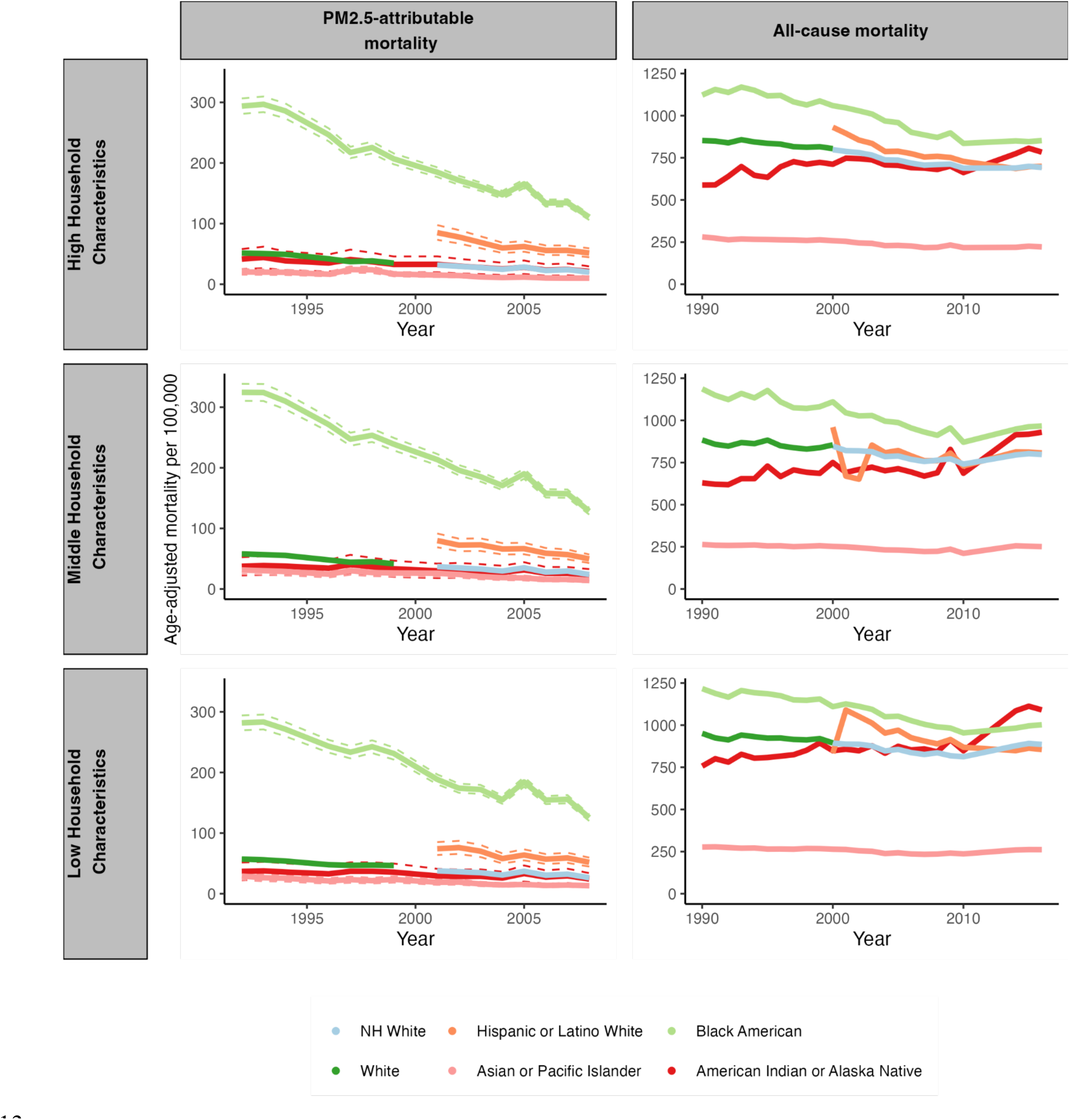
Age-adjusted PM_2.5_-attributable mortality rate and all-cause mortality rate for each racial/ethnic group stratified by household characteristics. The dashed lines depict 95% confidence intervals. Abbreviations: NH=Non-Hispanic.

**Fig. S 13.**
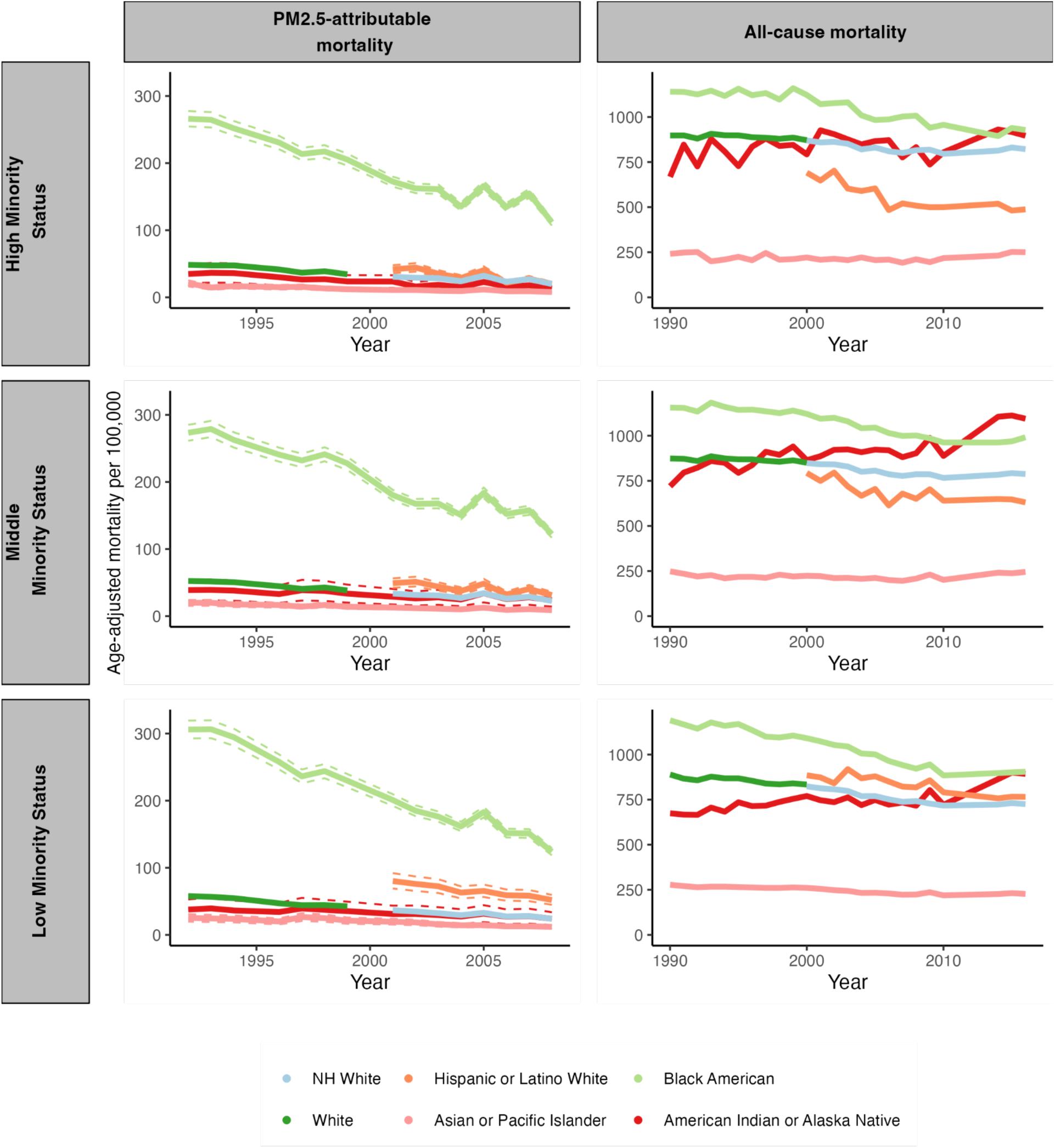
Age-adjusted PM_2.5_-attributable mortality rate and all-cause mortality rate for each racial/ethnic group stratified by minority status. The dashed lines depict 95% confidence intervals. Abbreviations: NH=Non-Hispanic.

**Fig. S 14.**
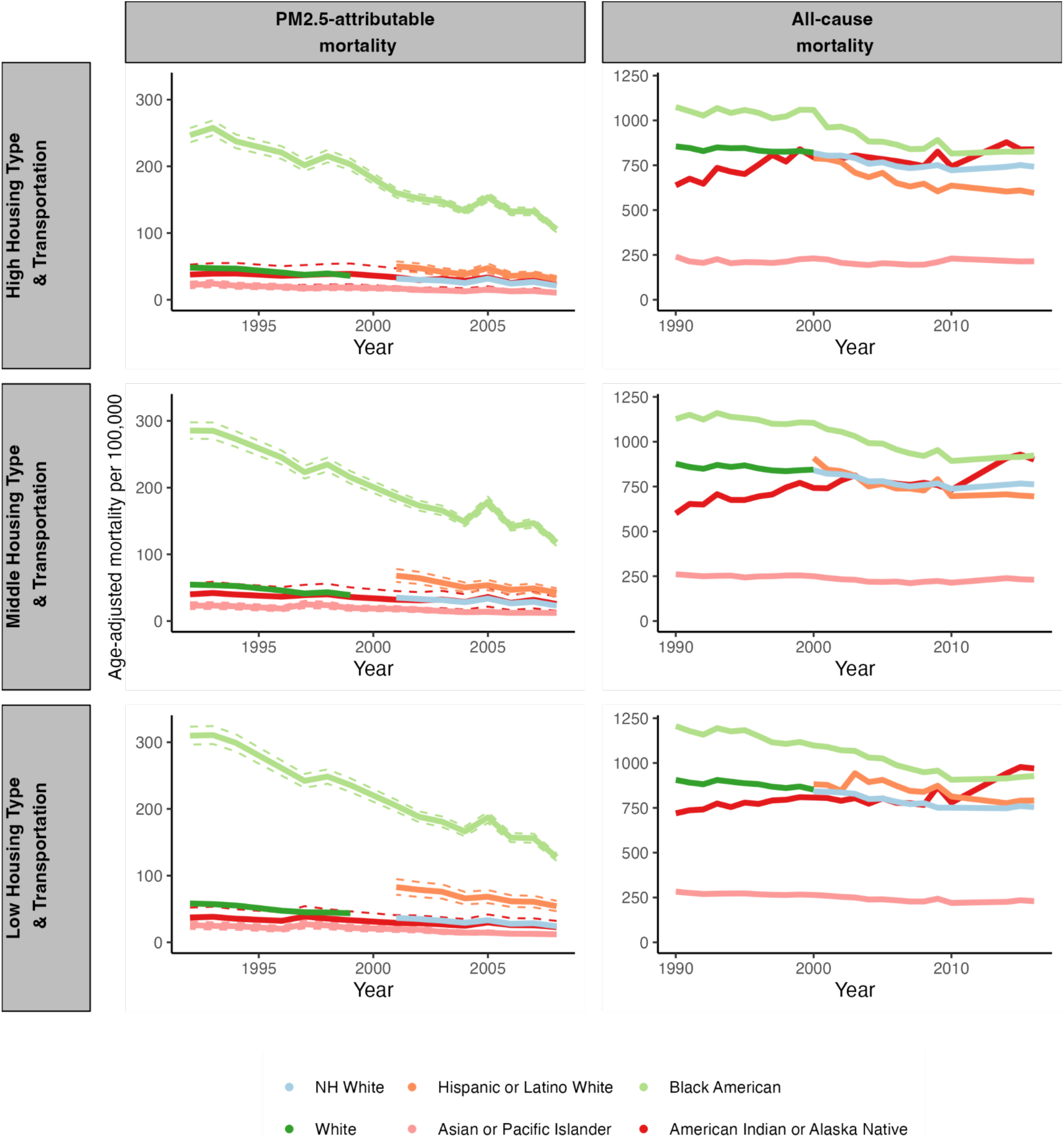
Age-adjusted PM_2.5_-attributable mortality rate and all-cause mortality rate for each racial/ethnic group stratified by housing type and transportation. The dashed lines depict 95% confidence intervals. Abbreviations: NH=Non-Hispanic.

**Fig. S 15.**
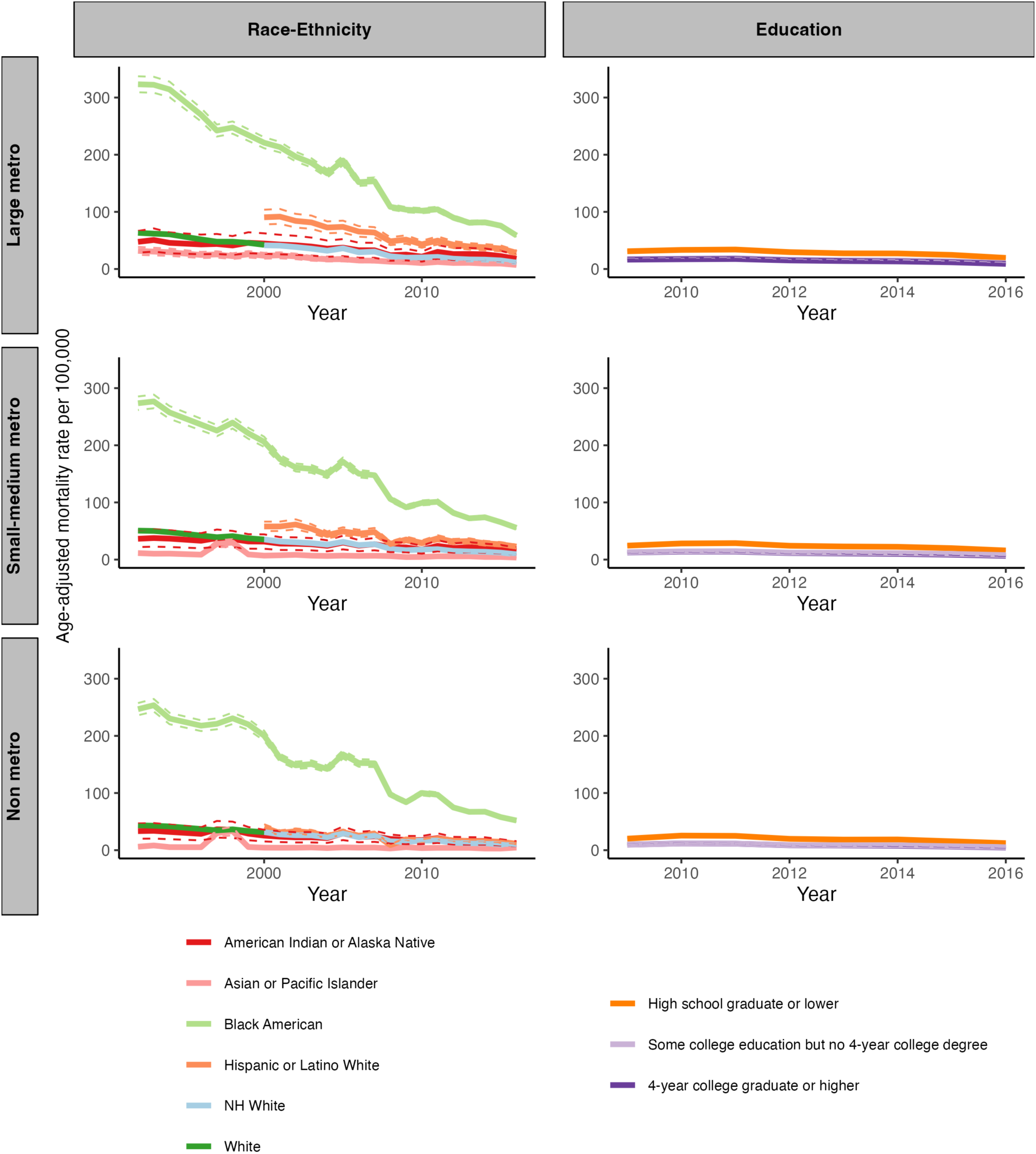
Age-adjusted PM_2.5_-attributable mortality rate for the age group 25+ years in the USA. The left column is by racial/ethnic group and rurality level. The right column is by education and rurality level. Abbreviations: NH=Non-Hispanic.

**Fig. S 16.**
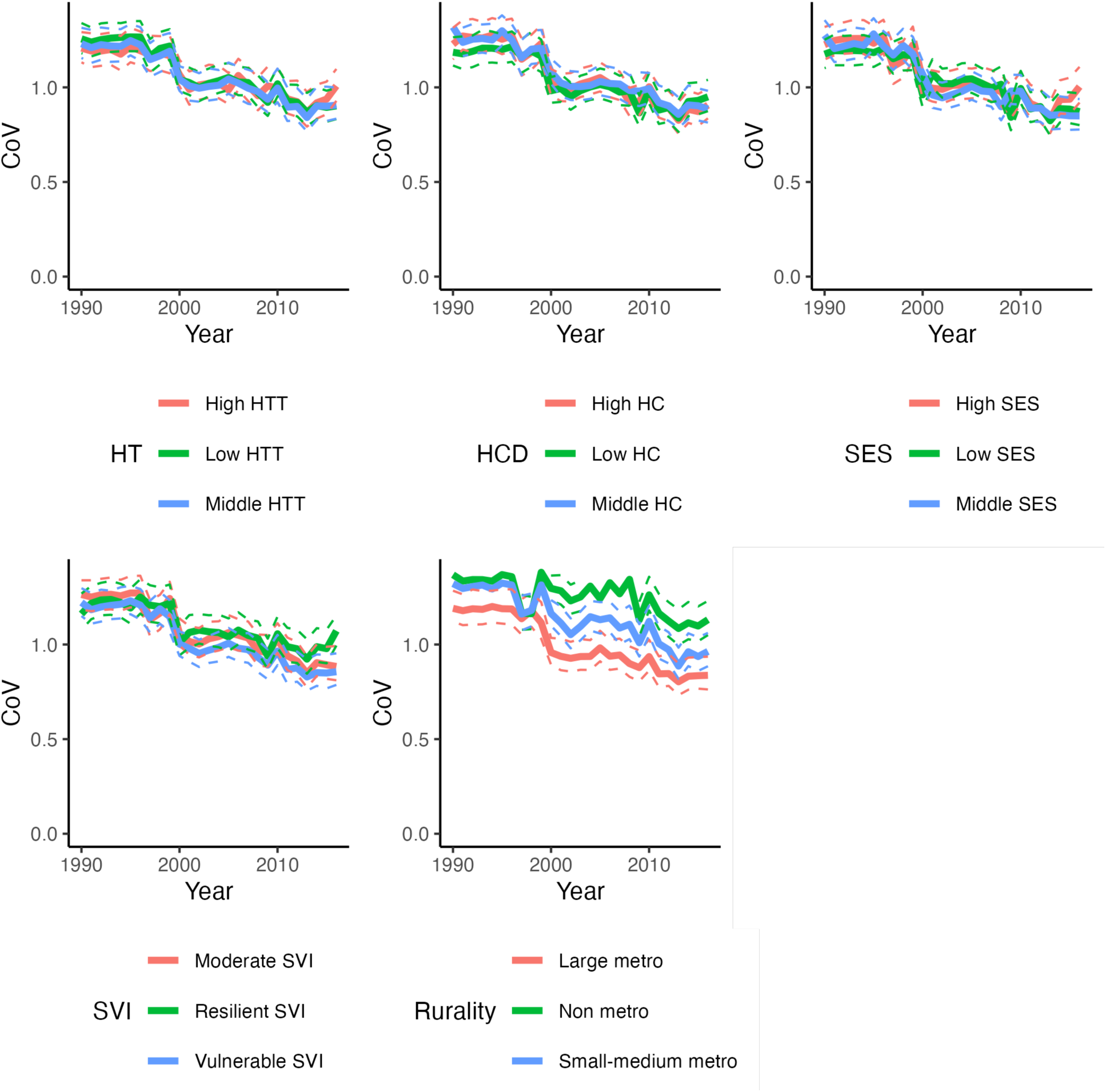
Coefficient of Variation (CoV) of age-adjusted all-cause mortality attributable to PM_2.5_ by racial/ethnic group for different sociodemographic groups for the age group 25+ years. The line for “High HC” corresponds to the CoV between the racial/ethnic group with the high household characteristics group. The CoV is the standard deviation of the age-adjusted PM2.5-attributable mortality rate divided by the mean age-adjusted PM2.5-attributable mortality rate. We excluded the analysis by racial/ethnic group and minority status because those variables are highly correlated. Abbreviations: SES = Socioeconomic Status, HC = Household Characteristics, HTT = Housing Type & Transportation.

**Fig. S 17.**
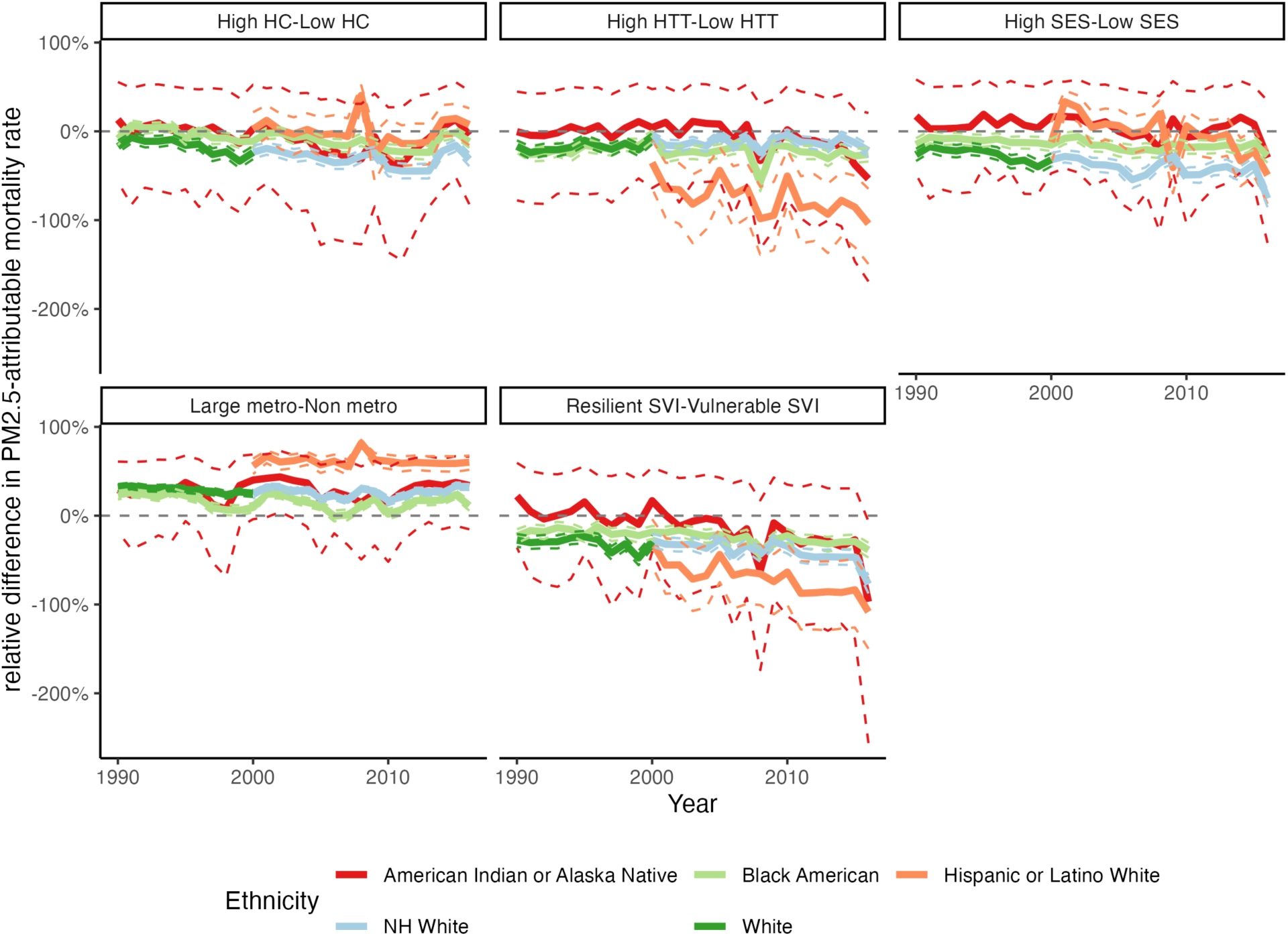
Relative difference in PM_2.5_-attributable mortality rate for each racial/ethnic group between different sociodemographic groups. The relative difference in PM2.5-attributable mortality is computed using the formula (PM2.5 attributable mortality 1 − PM2.5 attributable mortality 2)/PM2.5 attributable mortality 1. As an illustration, if the mortality rate for High HTT is 20 and for Low HTT is 36, the calculated relative difference would be (20 − 36)/20 = - 80%. Abbreviations: NH=Non-Hispanic, SES = Socioeconomic Status, HC = Household Characteristics, MS = Minority Status, HTT = Housing Type & Transportation, SVI = Social Vulnerability Index.

**Fig. S 18.**
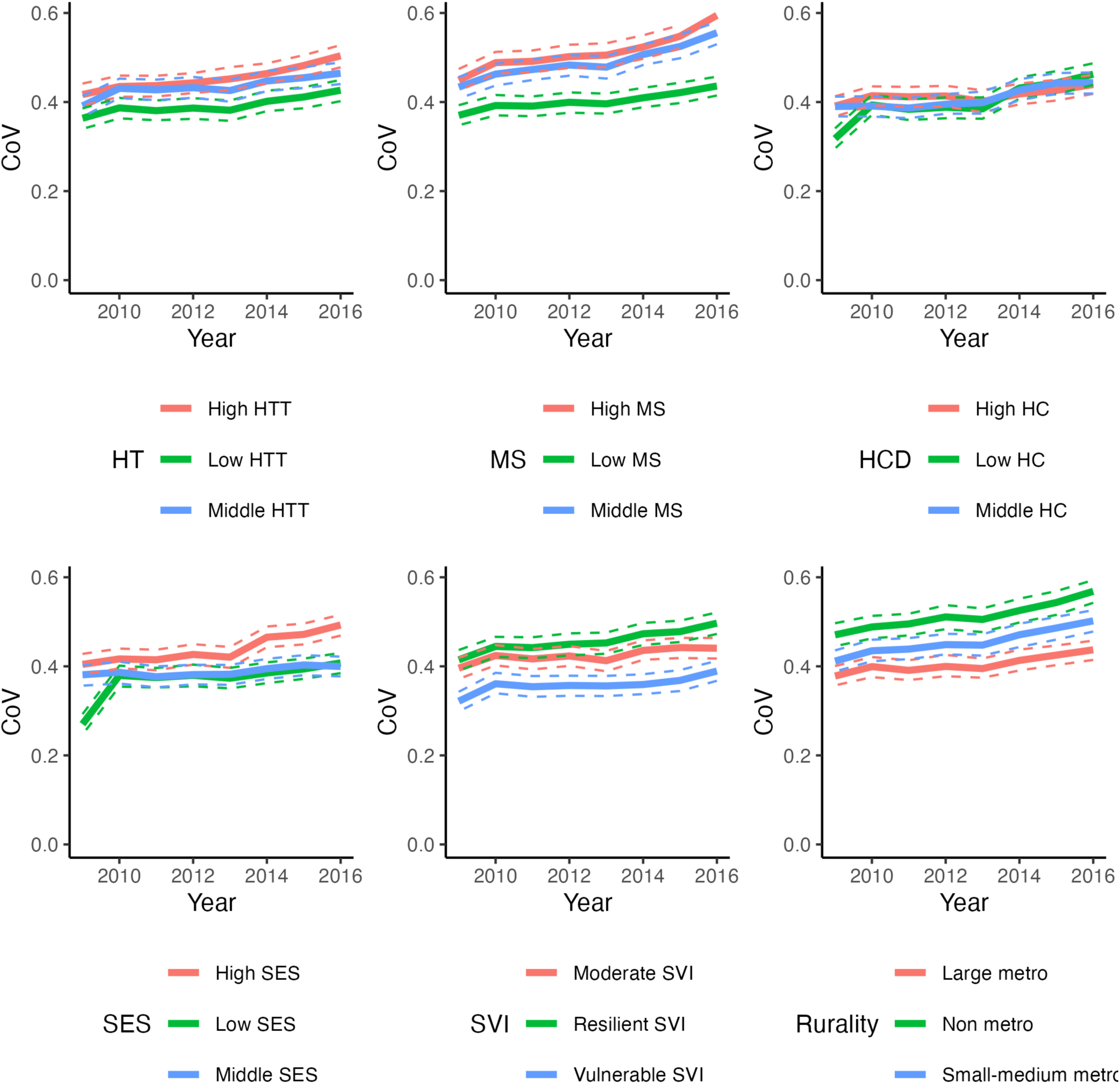
Coefficient of Variation (CoV) of age-adjusted all-cause mortality attributable to PM_2.5_ by educational attainment for different sociodemographic groups for the age group 25+ years. The line for “High HC” corresponds to the CoV between the educational attainment levels with the High Household Characteristics group. The CoV is the standard deviation of the age-adjusted PM2.5-attributable mortality rate divided by the mean age-adjusted PM2.5-attributable mortality rate. Abbreviations: SES = Socioeconomic Status, HC = Household Characteristics, MS = Minority Status, HTT = Housing Type & Transportation, SVI = Social Vulnerability Index.

**Fig. S 19.**
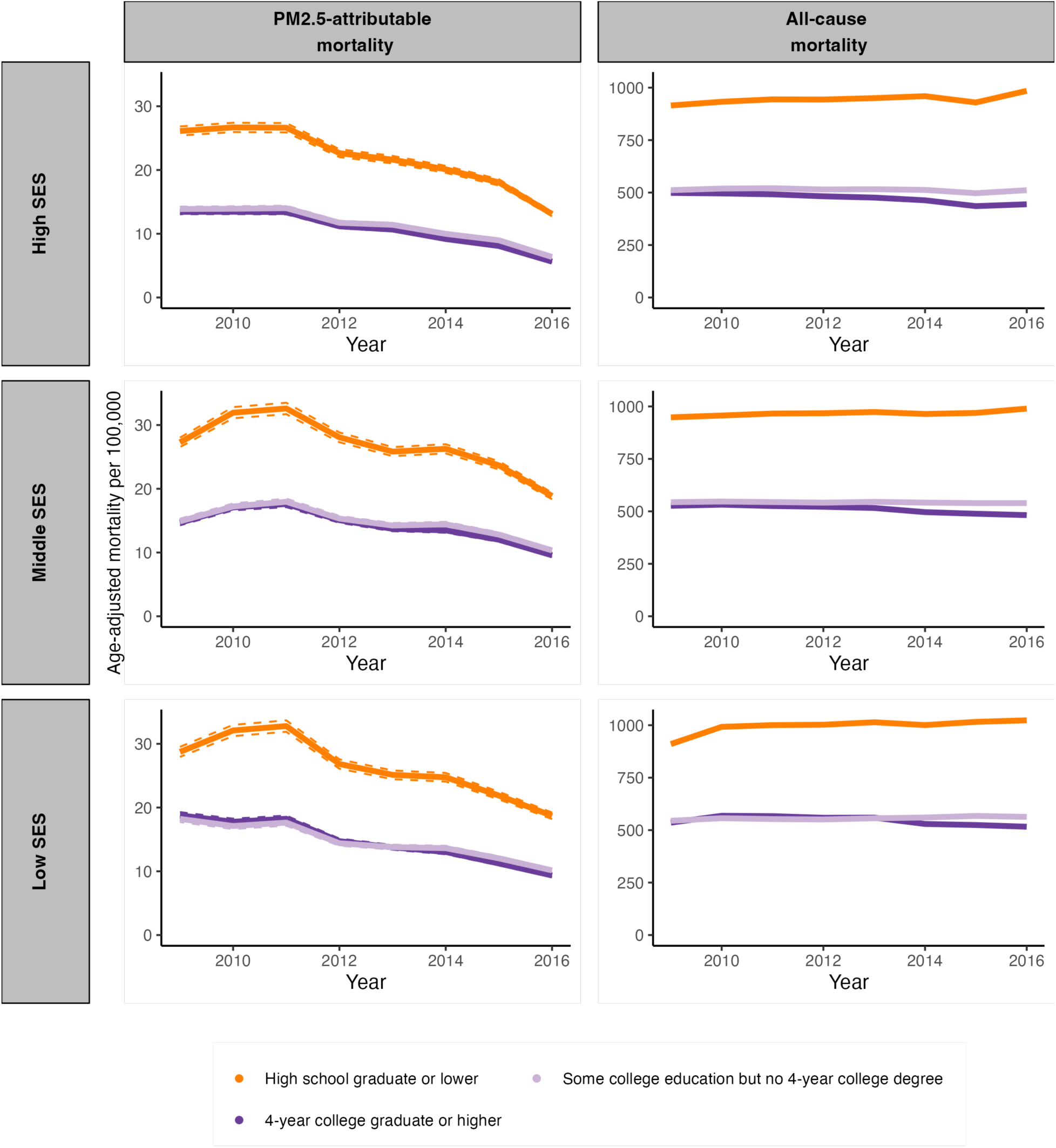
Age-adjusted PM_2.5_-attributable mortality rate and all-cause mortality rate for each education level stratified by socioeconomic status. The dashed lines depict 95% confidence intervals. Abbreviations: NH=Non-Hispanic, SES = socioeconomic status.

**Fig. S 20.**
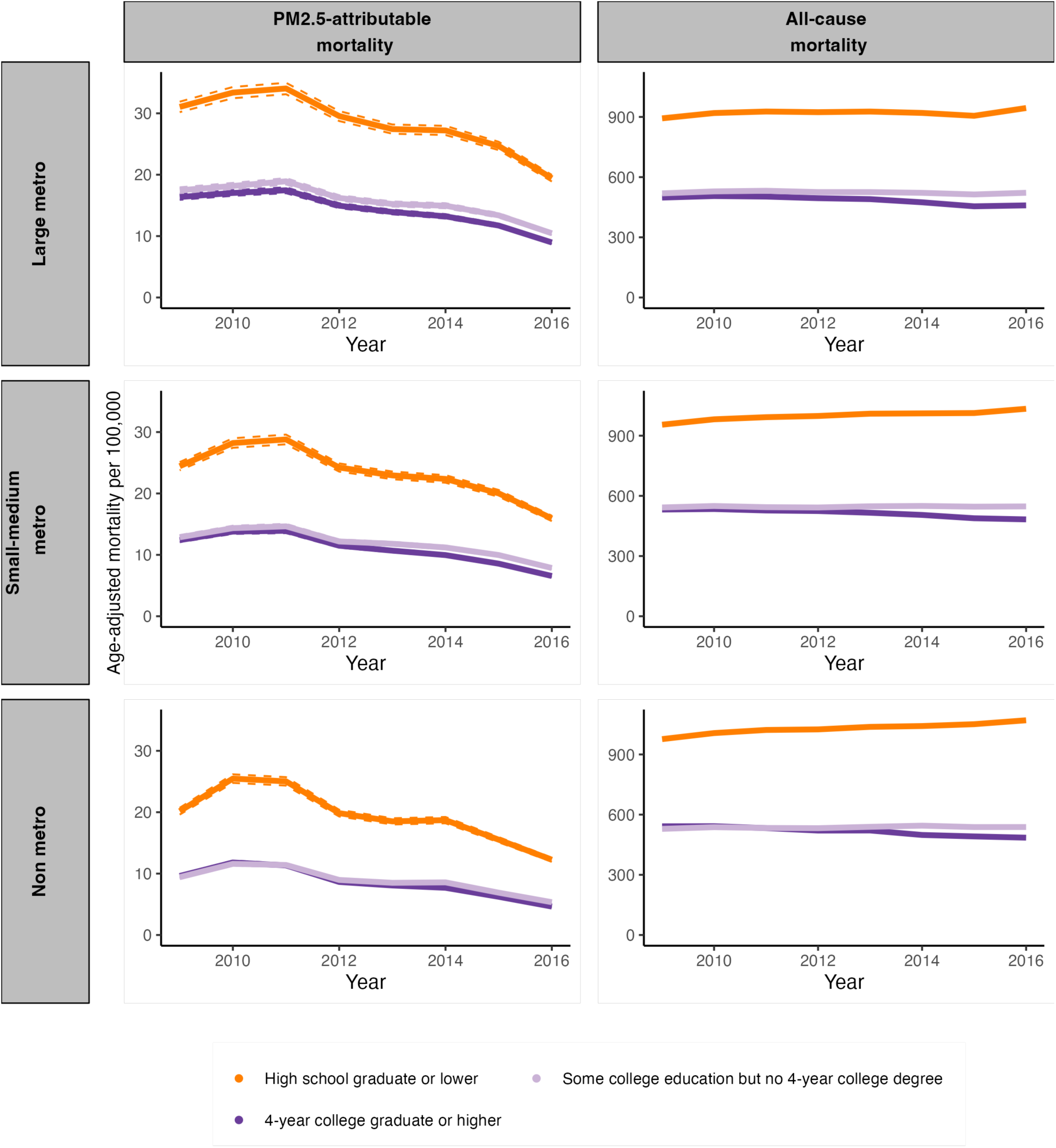
Age-adjusted PM_2.5_-attributable mortality rate and all-cause mortality rate for each education level stratified by rurality. The dashed lines depict 95% confidence intervals. Abbreviations: NH=Non-Hispanic.

**Fig. S 21.**
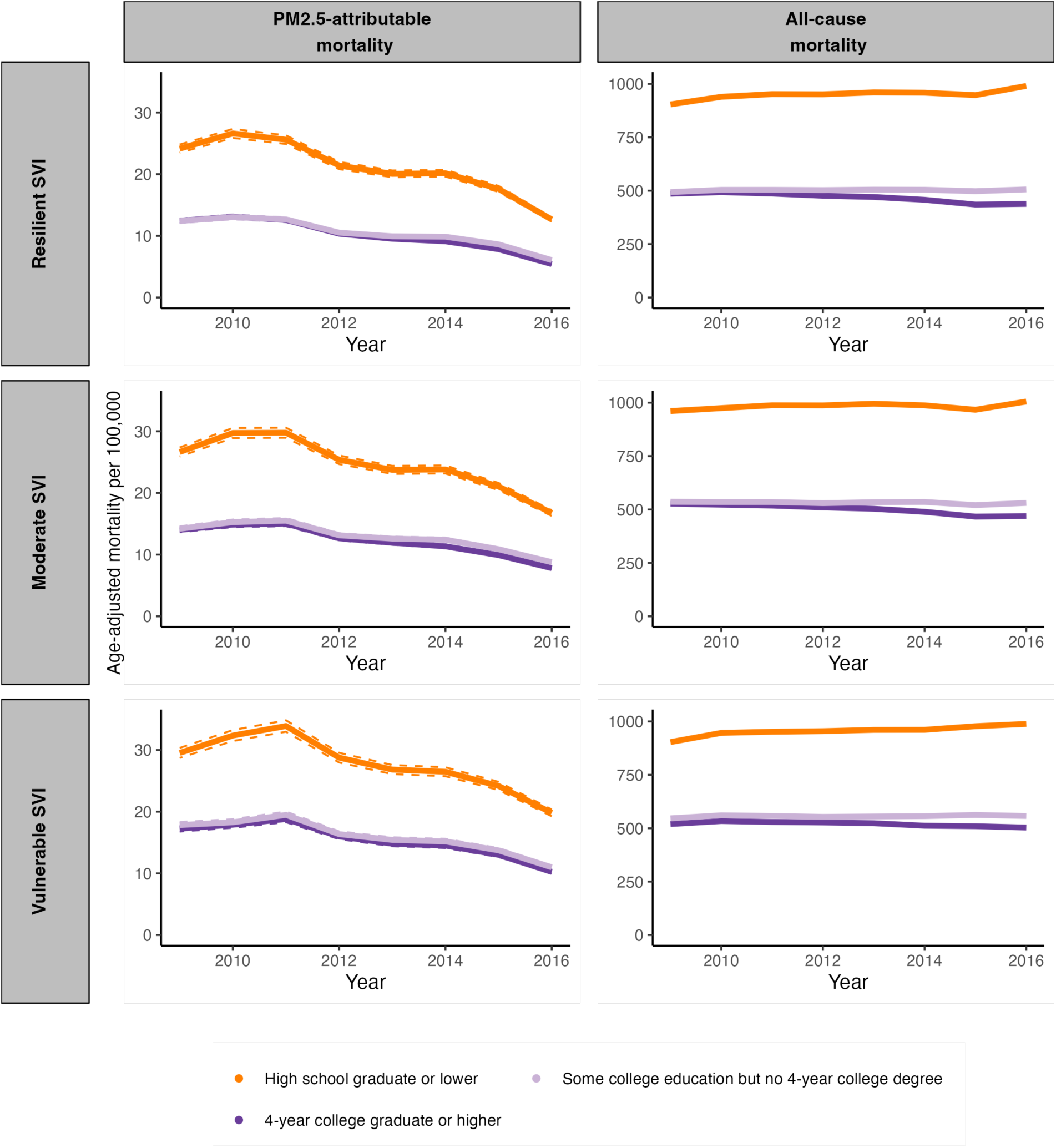
Age-adjusted PM_2.5_-attributable mortality rate and all-cause mortality rate for each education level stratified by social vulnerability index. The dashed lines depict 95% confidence intervals. Abbreviations: NH=Non-Hispanic.

**Fig. S 22.**
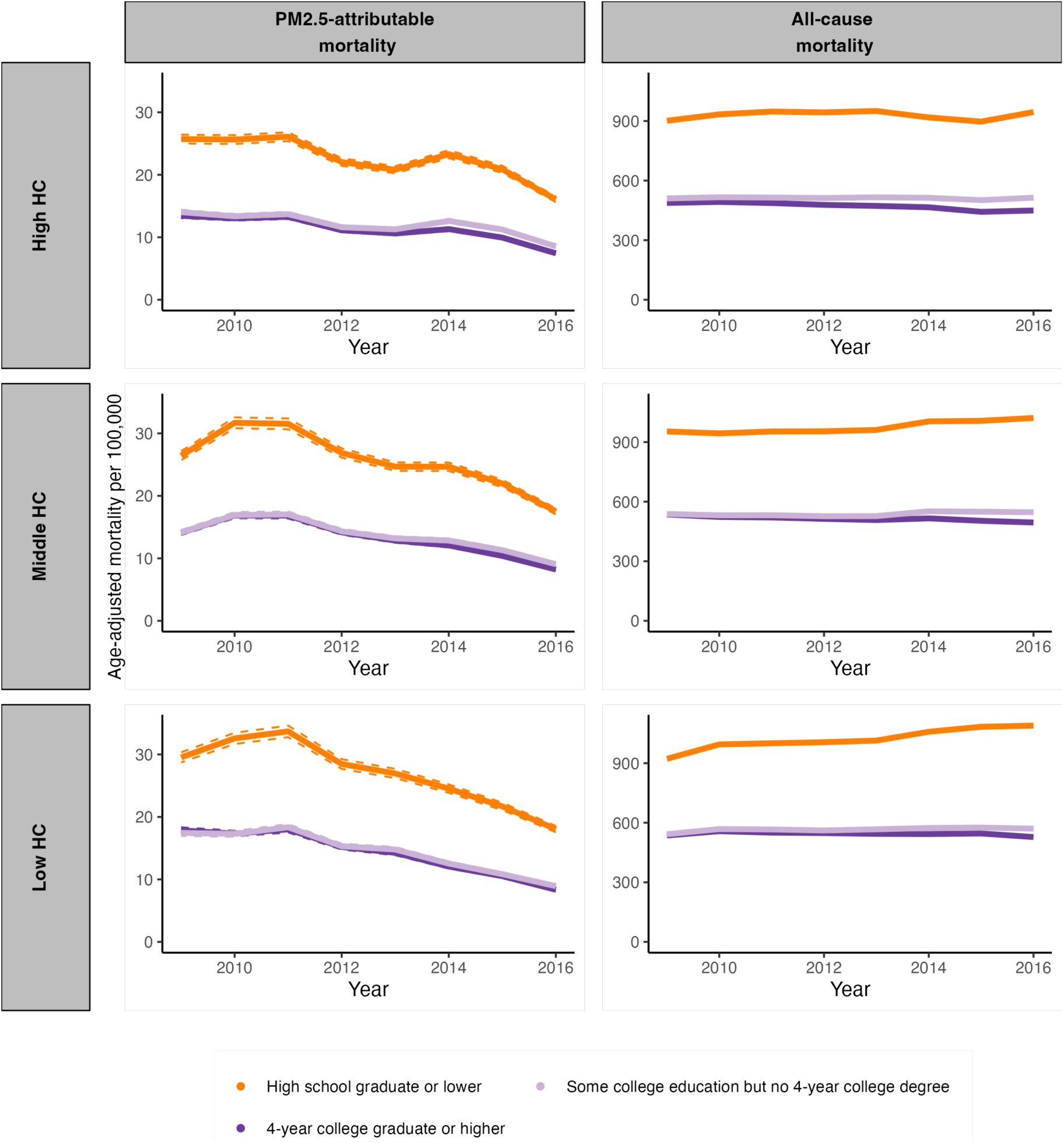
Age-adjusted PM_2.5_-attributable mortality rate and all-cause mortality rate for each education level stratified by household characteristic. The dashed lines depict 95% confidence intervals. Abbreviations: NH=Non-Hispanic, HC = household characteristic.

**Fig. S 23.**
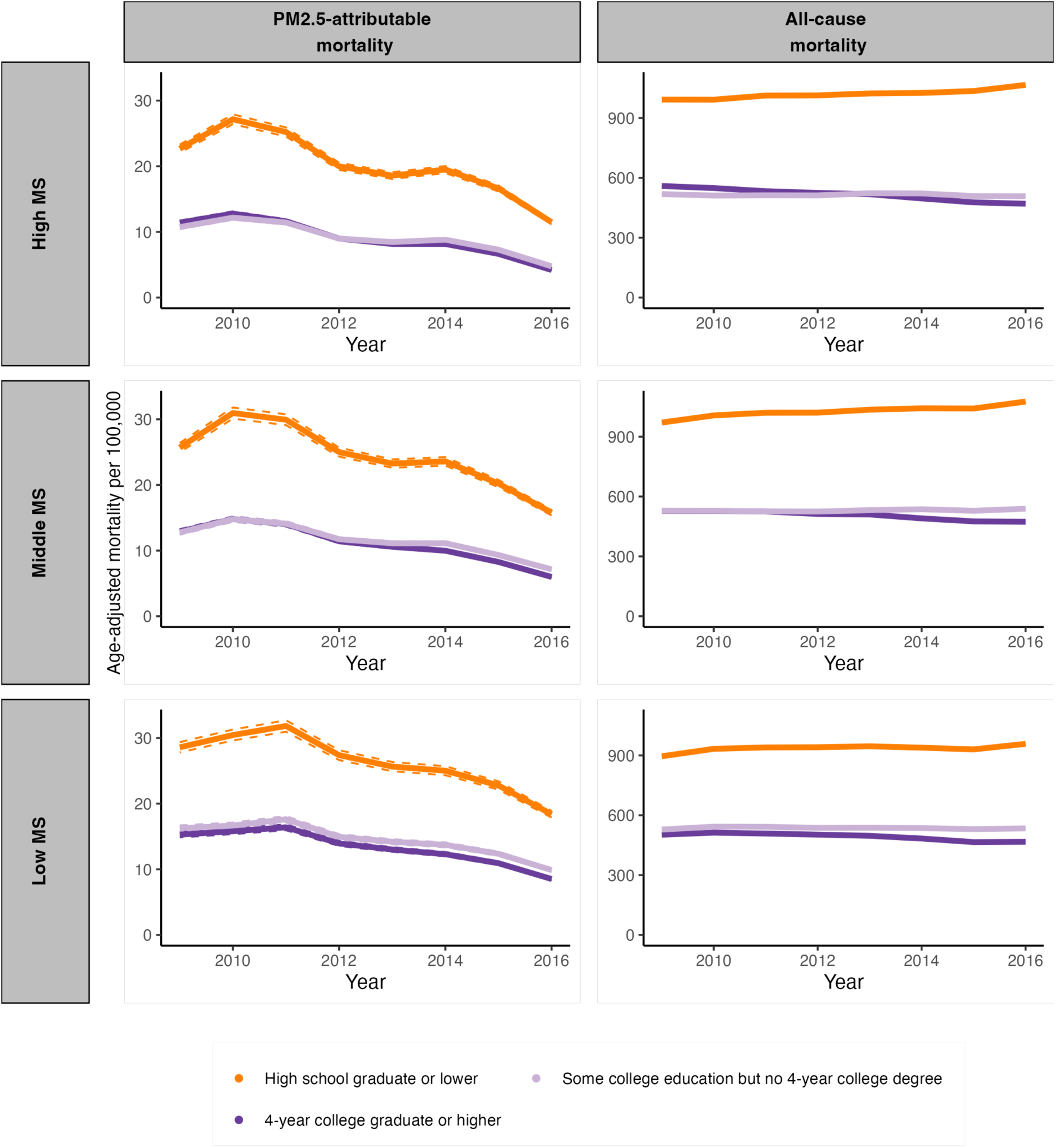
Age-adjusted PM_2.5_-attributable mortality rate and all-cause mortality rate for each education level stratified by minority status. The dashed lines depict 95% confidence intervals. Abbreviations: NH=Non-Hispanic, MS = minority status.

**Fig. S 24.**
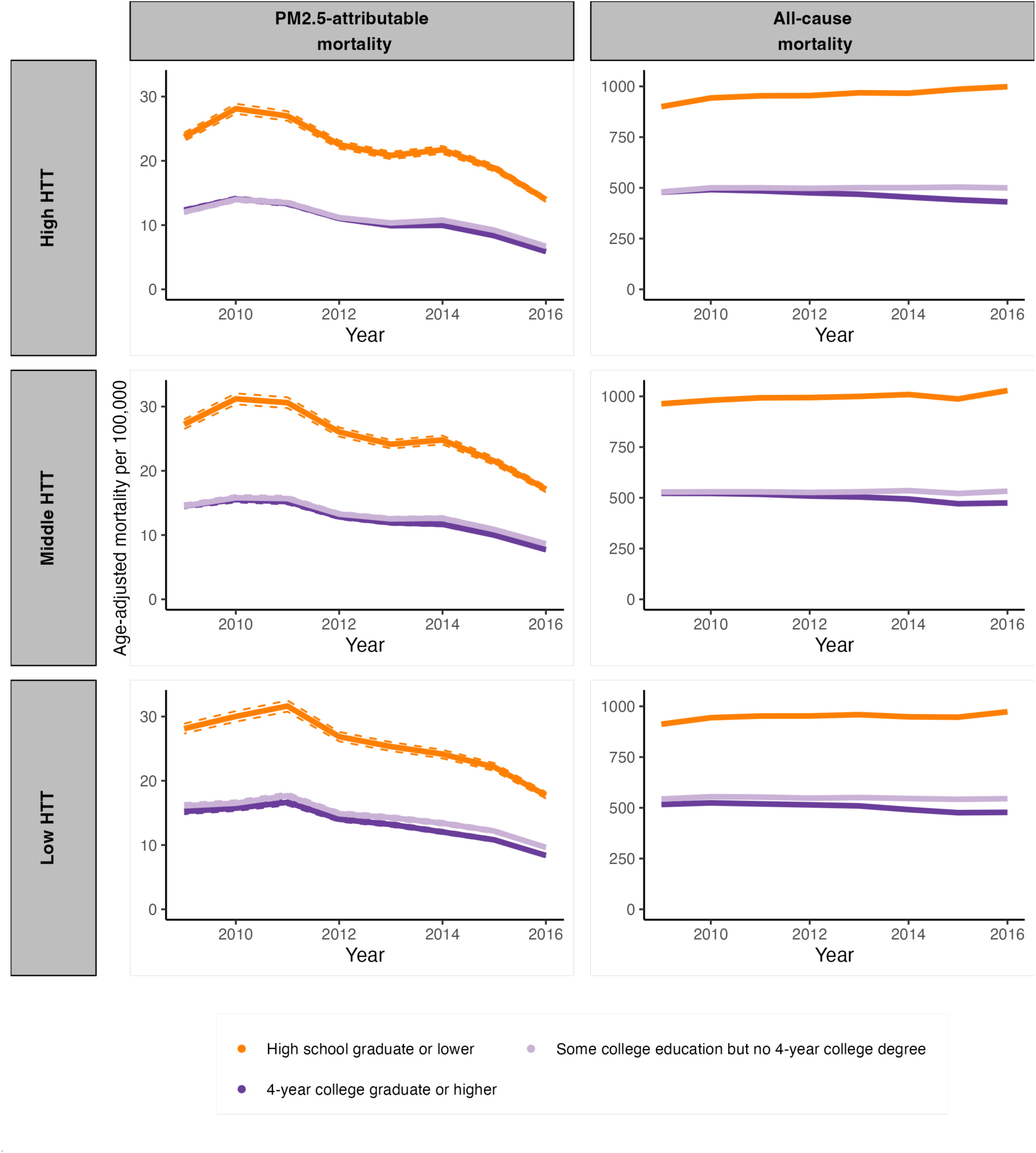
Age-adjusted PM_2.5_-attributable mortality rate and all-cause mortality rate for each education level stratified by housing type and transportation. The dashed lines depict 95% confidence intervals. Abbreviations: NH=Non-Hispanic, HTT = housing type and transportation.

**Fig. S 25.**
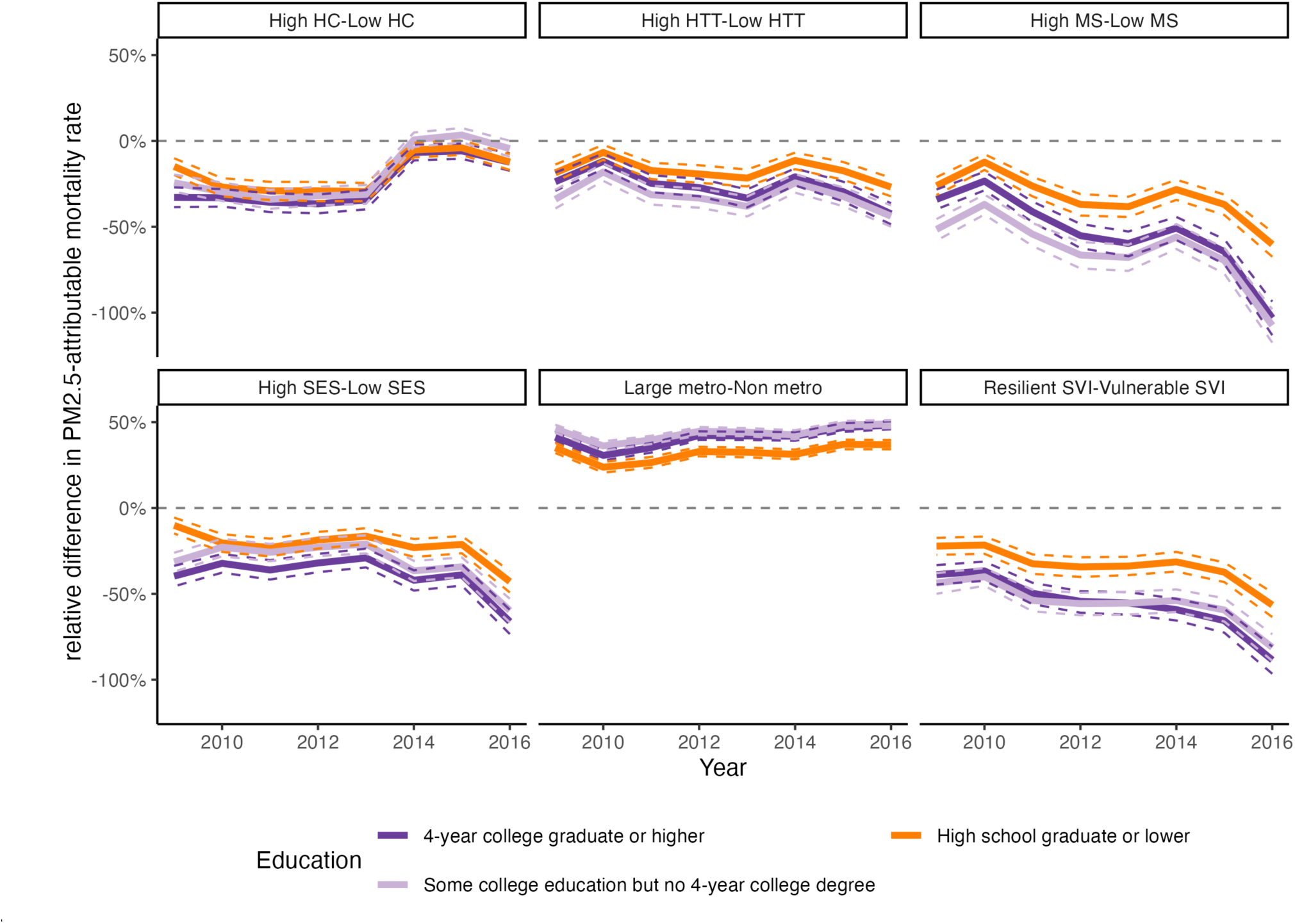
Relative difference in PM_2.5_-attributable mortality rate for each educational attainment group between different sociodemographic groups. The relative difference is calculated as (PM2.5 attributable mortality_1 - PM2.5 attributable mortality_2)/PM2.5 attributable mortality_1. For example, if High HTT = 20 and Low HTT = 36, then the relative difference is (20-36)/20 = -80%. Abbreviations: SES = Socioeconomic Status, HC = Household Characteristics, MS = Minority Status, HTT = Housing Type & Transportation, SVI = Social Vulnerability Index.

**Fig. S 26.**
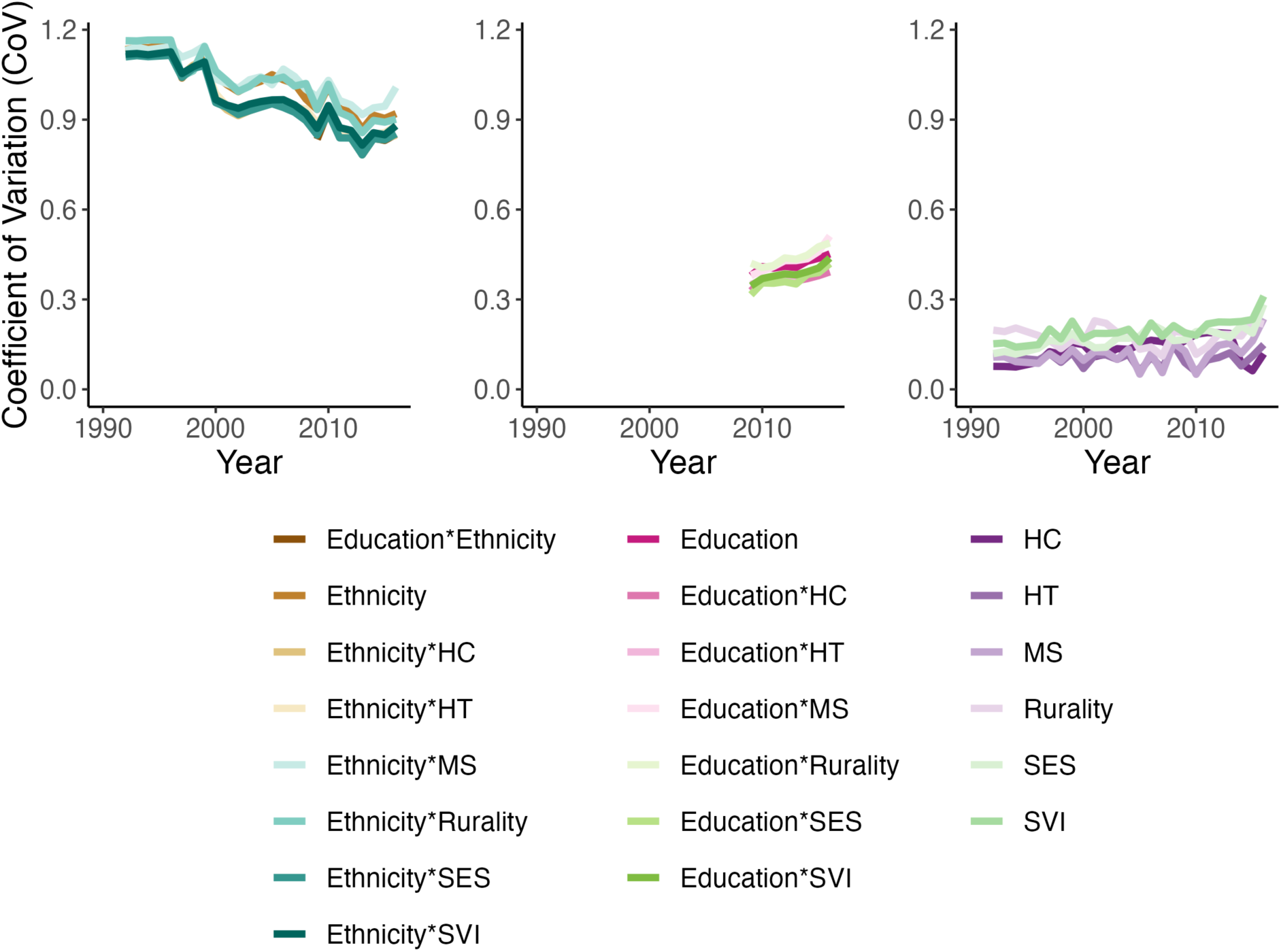
Coefficient of Variation (CoV) of age-adjusted all-cause mortality attributable to PM_2.5_ when stratifying by different sociodemographic groups for the age group 25+ years. The line for “Ethnicity*rural_urban_class” corresponds to the CoV between the 15 racial/ethnic and urbanicity group combinations available. The CoV is the standard deviation of the age-adjusted PM2.5-attributable mortality rate divided by the mean age-adjusted PM2.5-attributable mortality rate. Abbreviations: SES = Socioeconomic Status, HC = Household Characteristics, MS = Minority Status, HTT = Housing Type & Transportation, SVI = Social Vulnerability Index.

**Fig. S 27.**
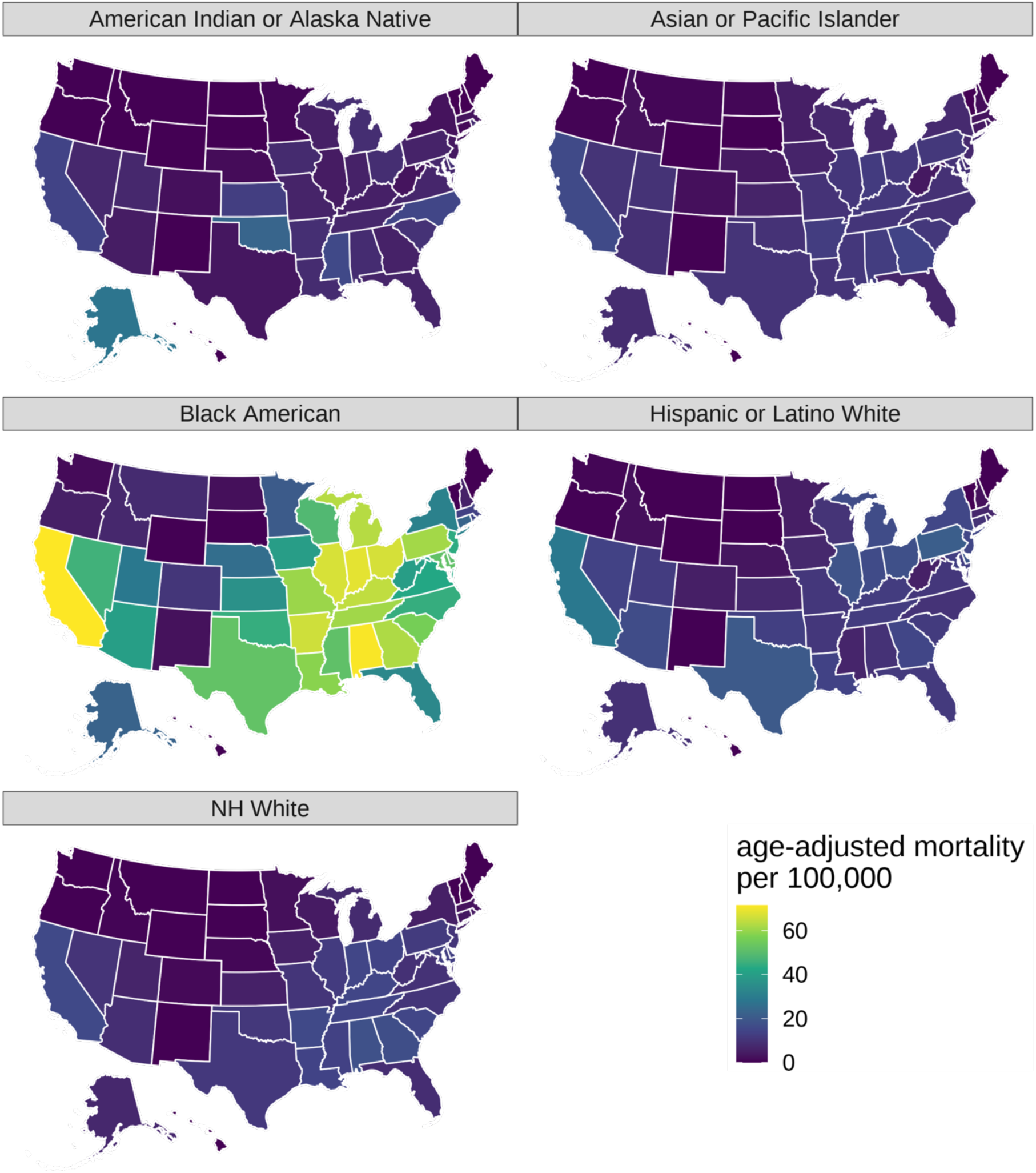
Age-adjusted PM_2.5_-attributable mortality rate for the age group 25+ years in the year 2016, by state and racial/ethnic group. Abbreviations: NH=Non-Hispanic.

**Fig. S 28.**
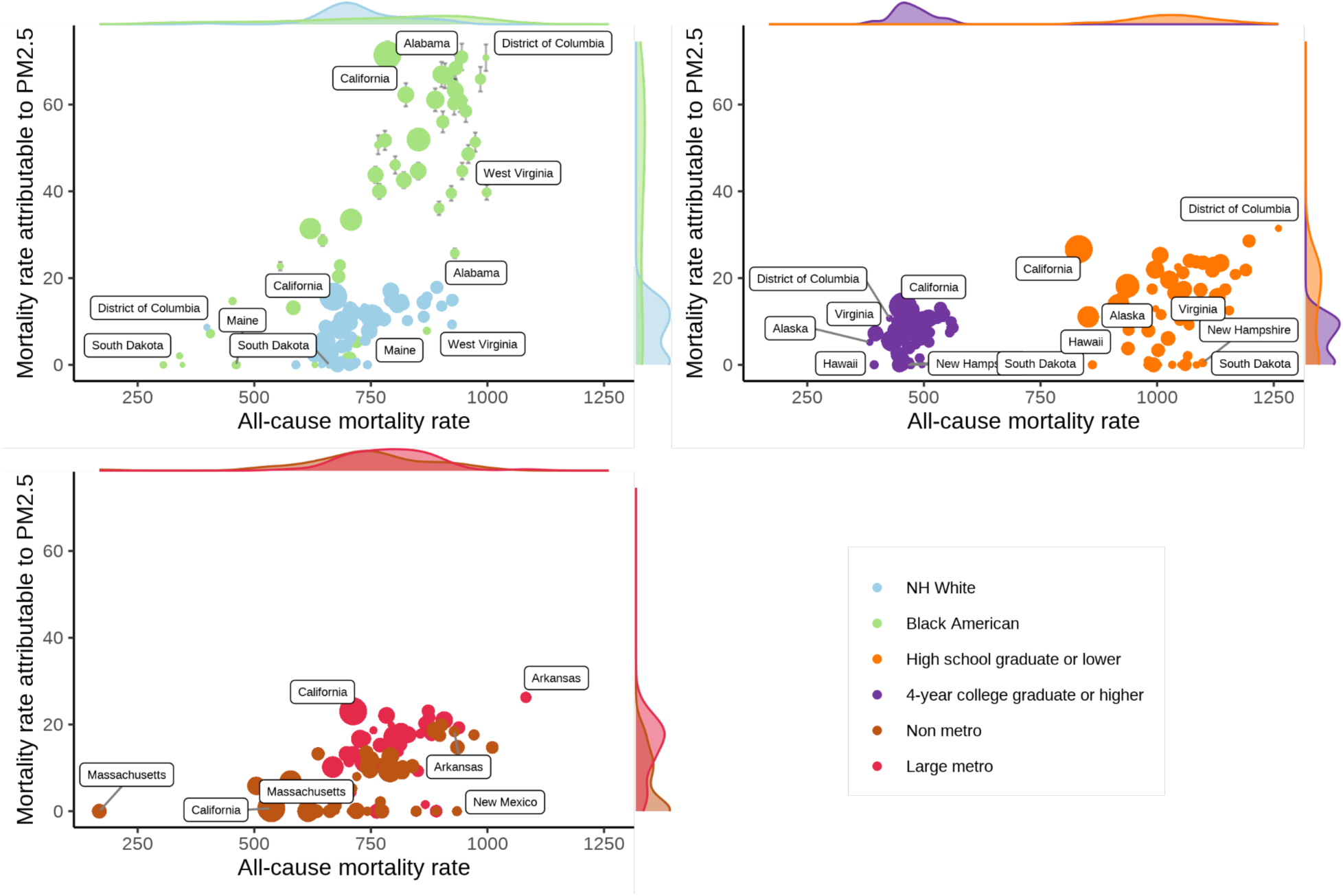
Age-adjusted mortality per 100,000 from all causes and attributable to PM2.5 (among those aged 25+ years) in US states and the District of Columbia in 2016 for the racial/ethnic groups “Black American” and “NH White”, low and high educational attainment, and rurality levels “Non-metro” and “large metro”. The icon area for states is proportional to the state’s population size. The most extreme points are labeled. Shaded areas are marginal kernel density estimates. Error bars denote 95% confidence intervals. Abbreviations: NH=Non-Hispanic.

**Fig. S 29.**
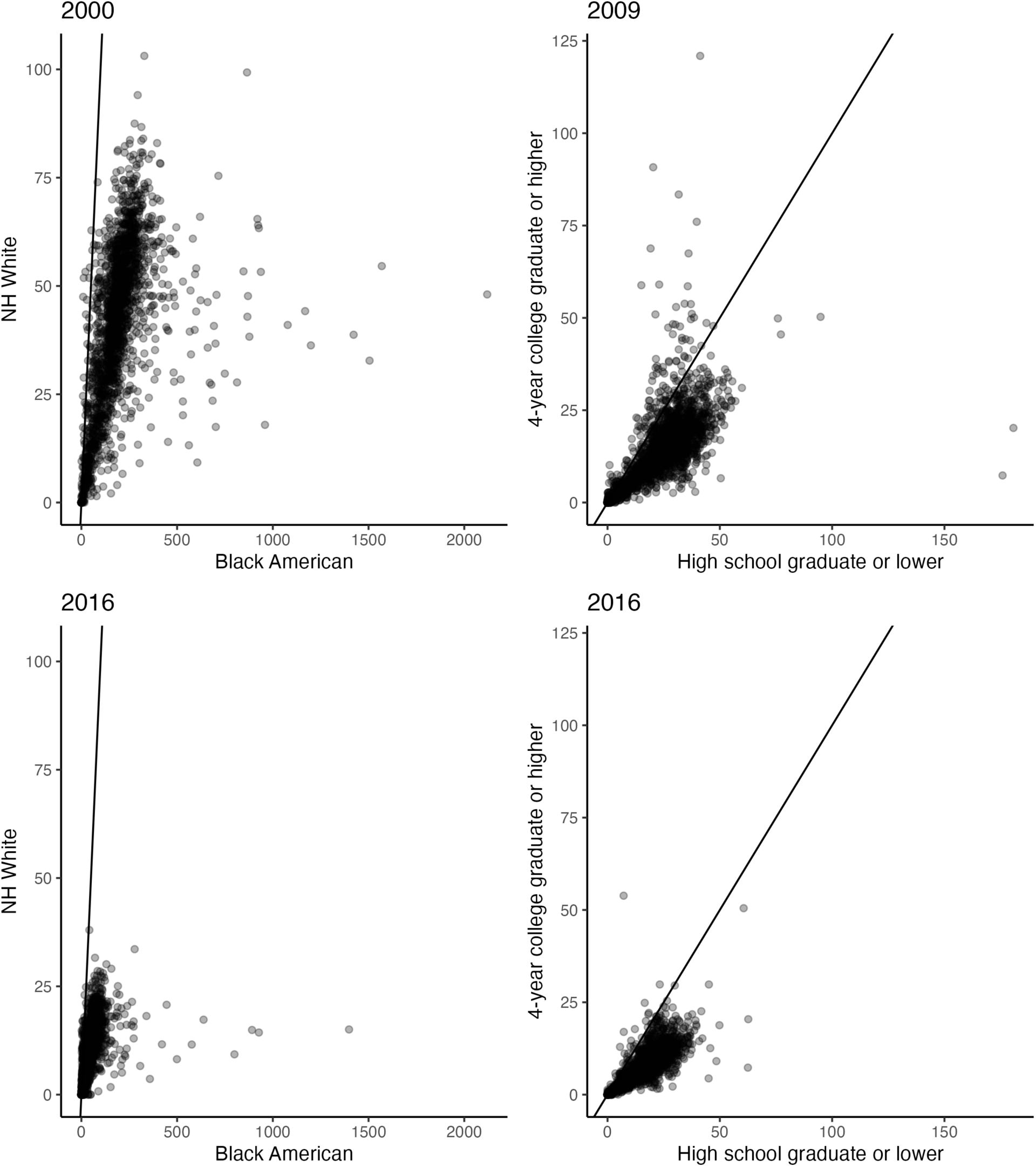
County-level scatter plot of the age-adjusted PM_2.5_-attributable mortality per 100,000 for selected racial/ethnic groups and education levels. These estimates are for the population aged 25+ years in the USA. Each point represents one county. Points on the straight line imply that the age-adjusted PM2.5-attributable mortality rate was the same for the group denoted on the x-axis as for the group denoted on the y-axis. Points below the straight line imply that the age-adjusted PM2.5-attributable mortality rate for the group on the x-axis was higher than for the group on the y-axis. Abbreviations: NH=Non-Hispanic.

**Fig. S 30.**
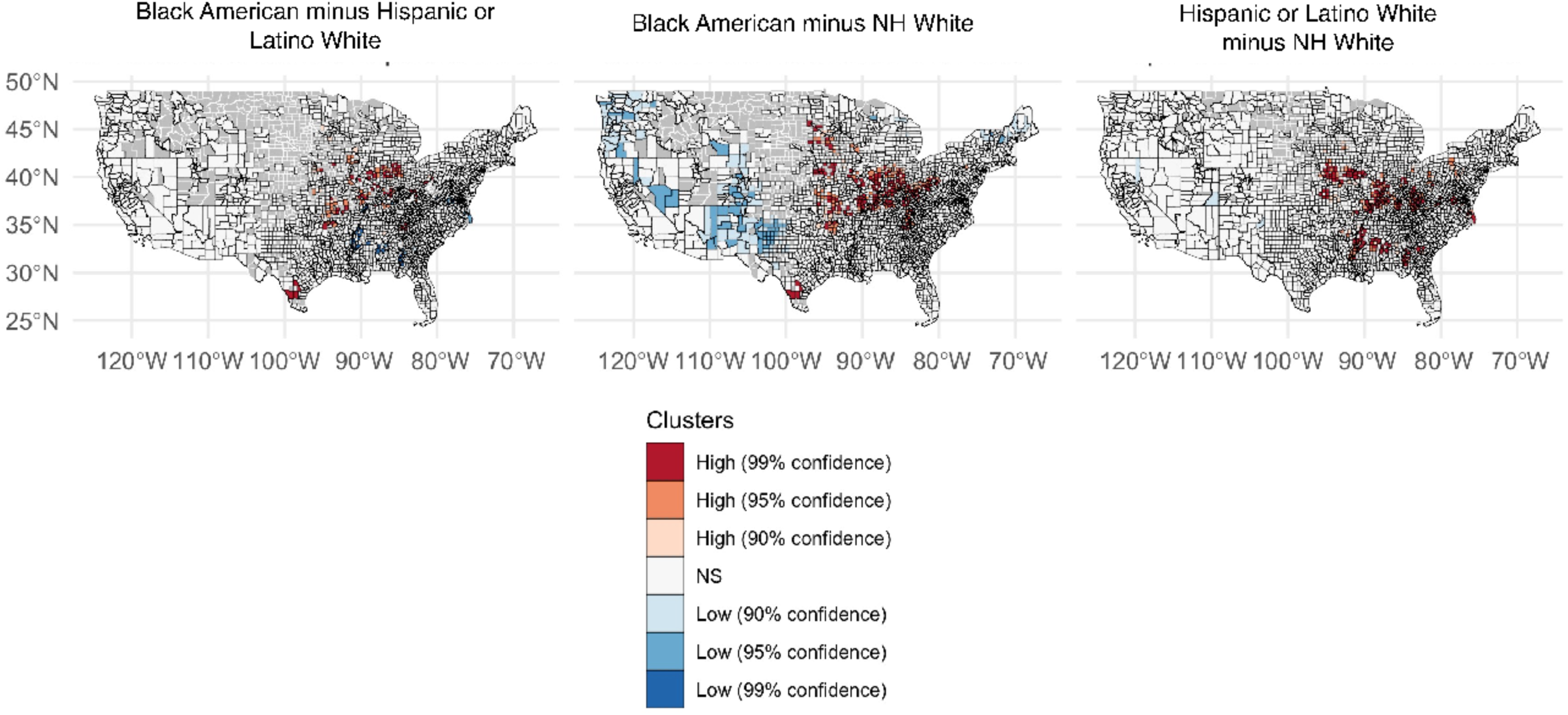
County-level spatial clusters with high (red colors) and low (blue colors) differences in the age-adjusted PM_2.5_-attributable mortality rate between racial/ethnic groups. These maps were created for the (unweighted) mean across the period 2000 to 2016. Clusters were estimated with Local Spatial Autocorrelation as Local Getis-Ord Gi*. See ’Spatial analyses’ in Materials and Methods for details. Abbreviations: NH=Non-Hispanic.

**Fig. S 31.**
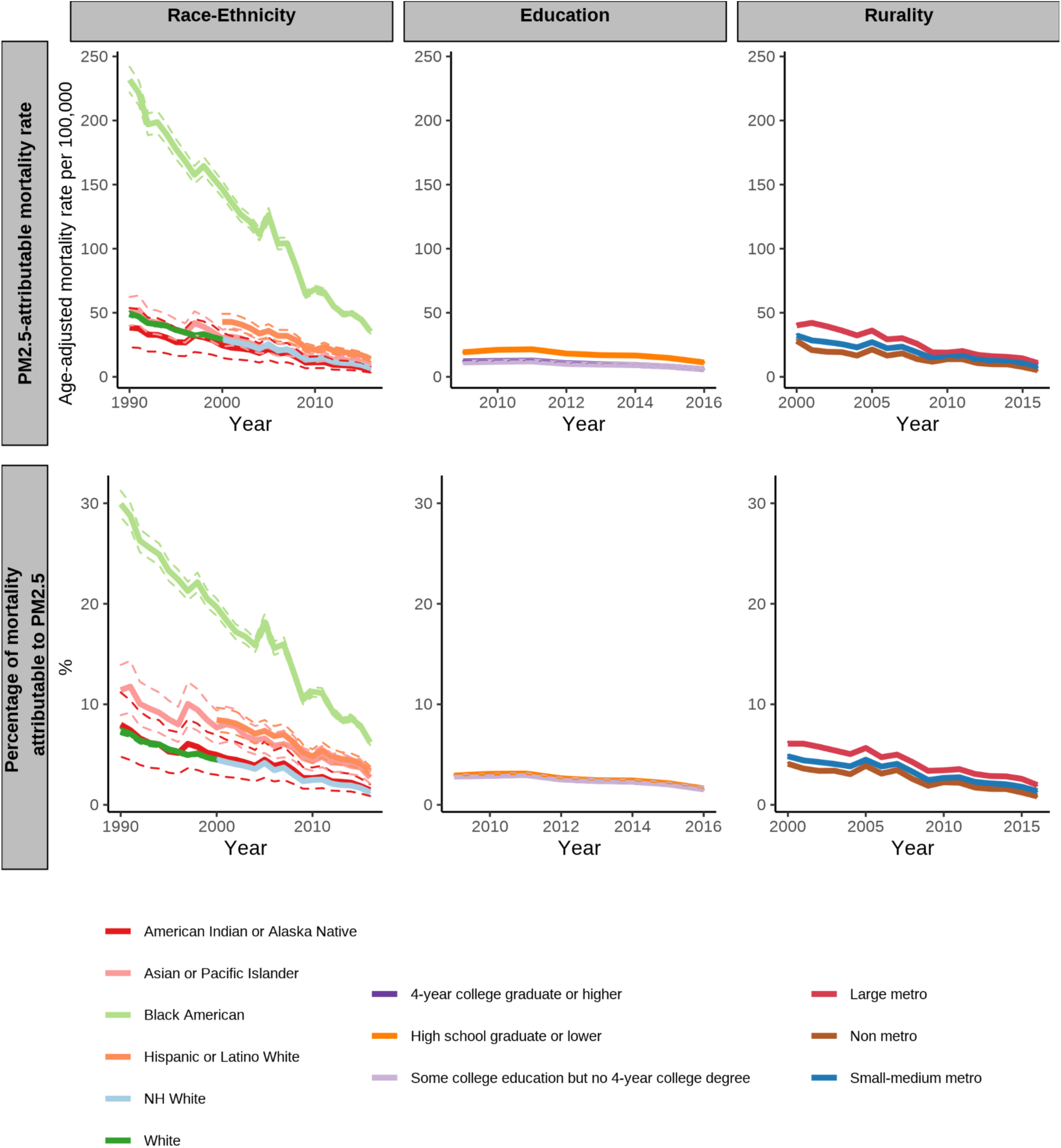
Age-adjusted PM_2.5_-attributable mortality rate by racial/ethnic group, education level, and rurality level for the age group 65+. The first row shows the age-adjusted mortality rate that we estimate is attributable to PM2.5. The second row shows the percentage of all-cause mortality that we estimate is attributable to PM2.5. The dashed lines depict 95% confidence intervals. 95% confidence intervals for rurality are too narrow to be visible. Abbreviations: NH=Non-Hispanic.

**Fig. S 32.**
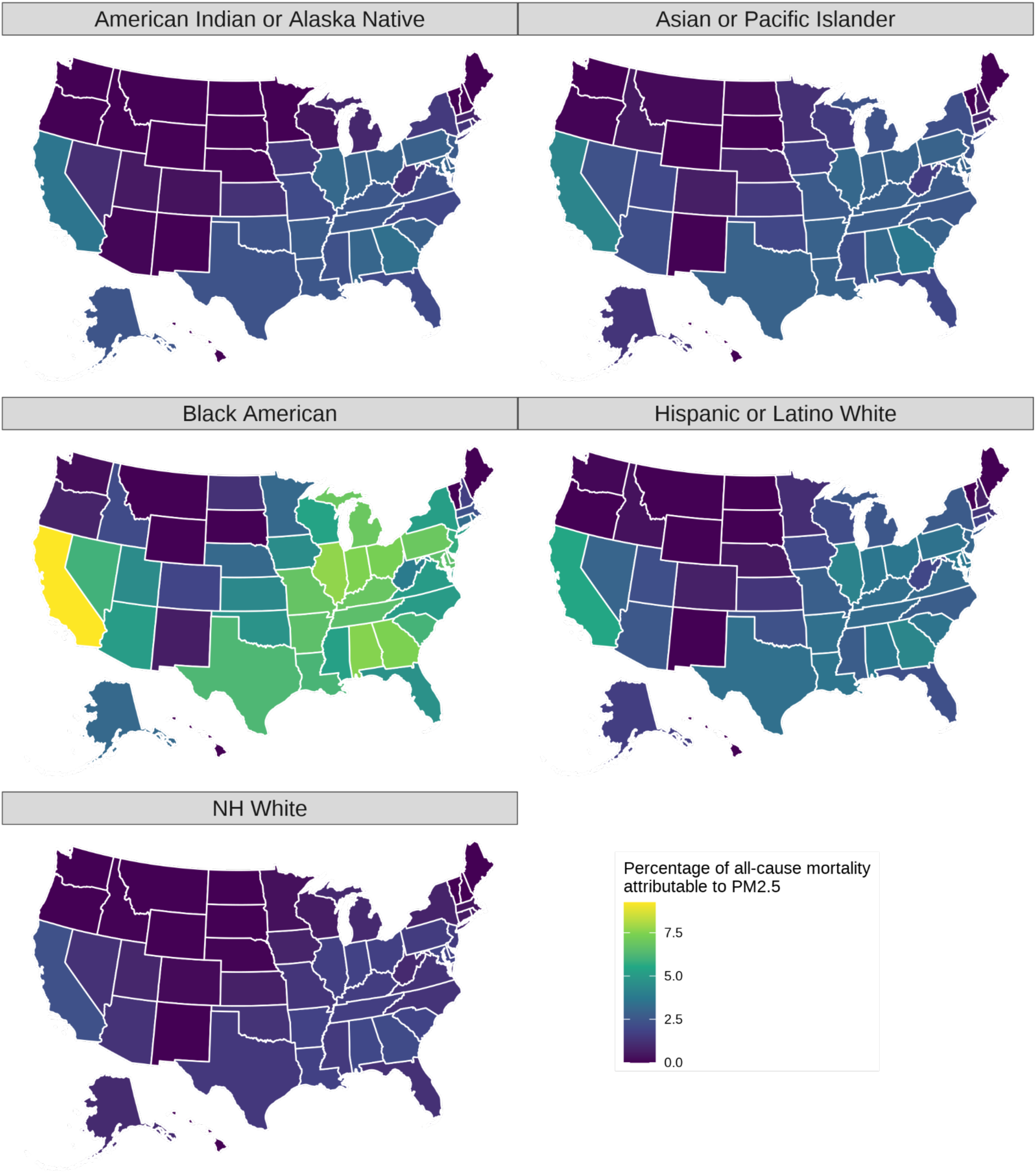
Percentage of the age-adjusted all-cause mortality rate that was attributable to PM_2.5_ in the year 2016, by state and racial/ethnic group for the age group 65+. Abbreviations: NH=Non-Hispanic.

**Fig. S 33.**
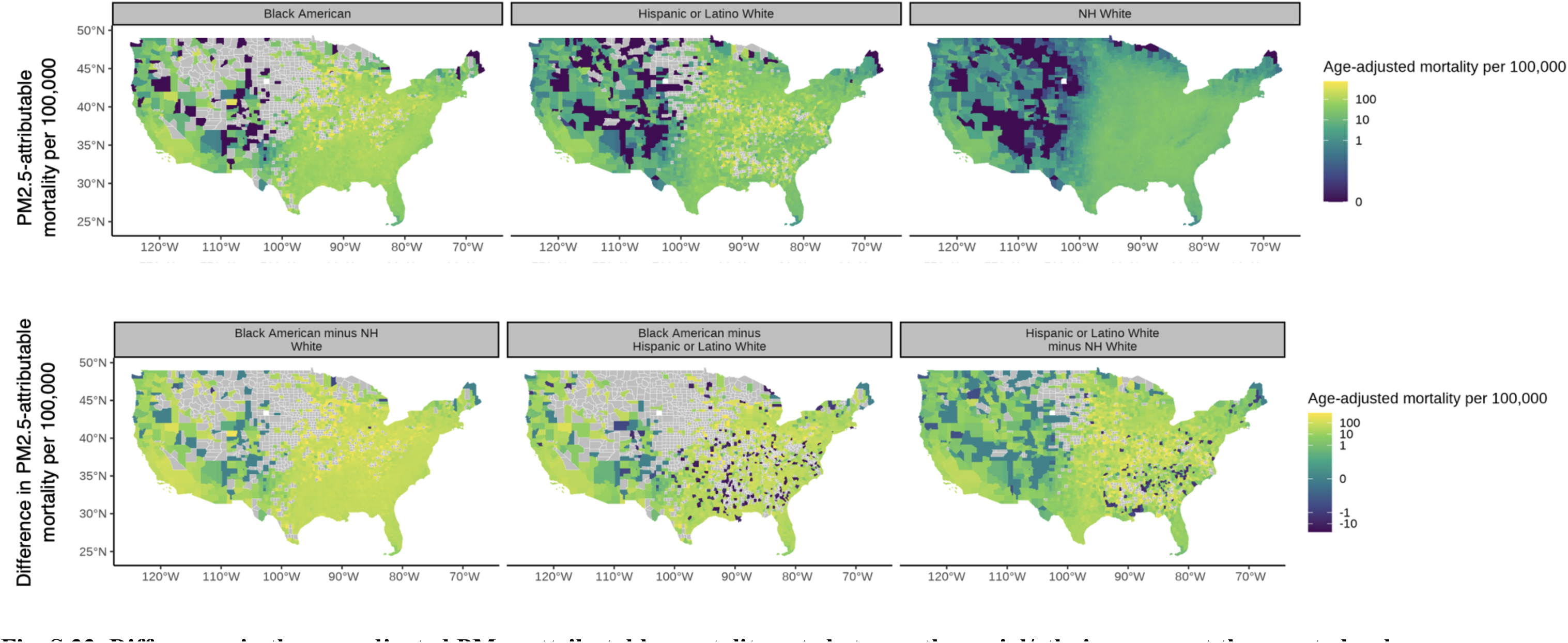
Differences in the age-adjusted PM_2.5_-attributable mortality rate between the racial/ethnic groups at the county level for the period 2000 to 2016 for the age group 65+ years. The first row is the PM2.5-attributable mortality per 100,000 for each racial/ethnic group. The second row is the absolute difference in PM2.5-attributable mortality per 100,000 between racial/ethnic groups. Abbreviations: NH=Non-Hispanic

**Fig. S 34.**
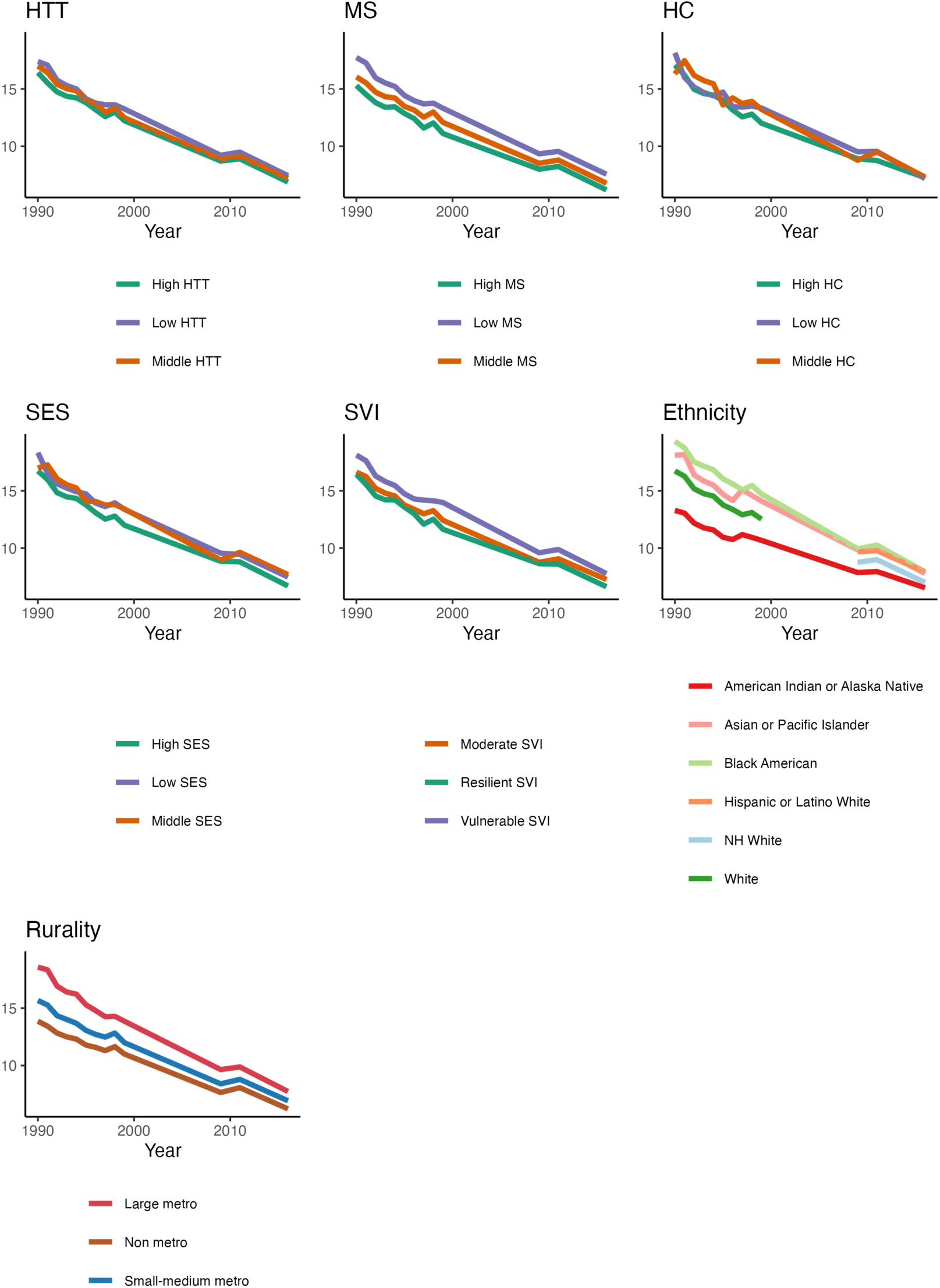
Population-weighted annual mean PM_2.5_ exposure in μg/m^3^ by subpopulation for the age group 65+ years. Abbreviations: NH=Non-Hispanic, SES = Socioeconomic Status, HC = Household Characteristics, MS = Minority Status, HTT = Housing Type & Transportation, SVI = Social Vulnerability Index.

**Fig. S 35.**
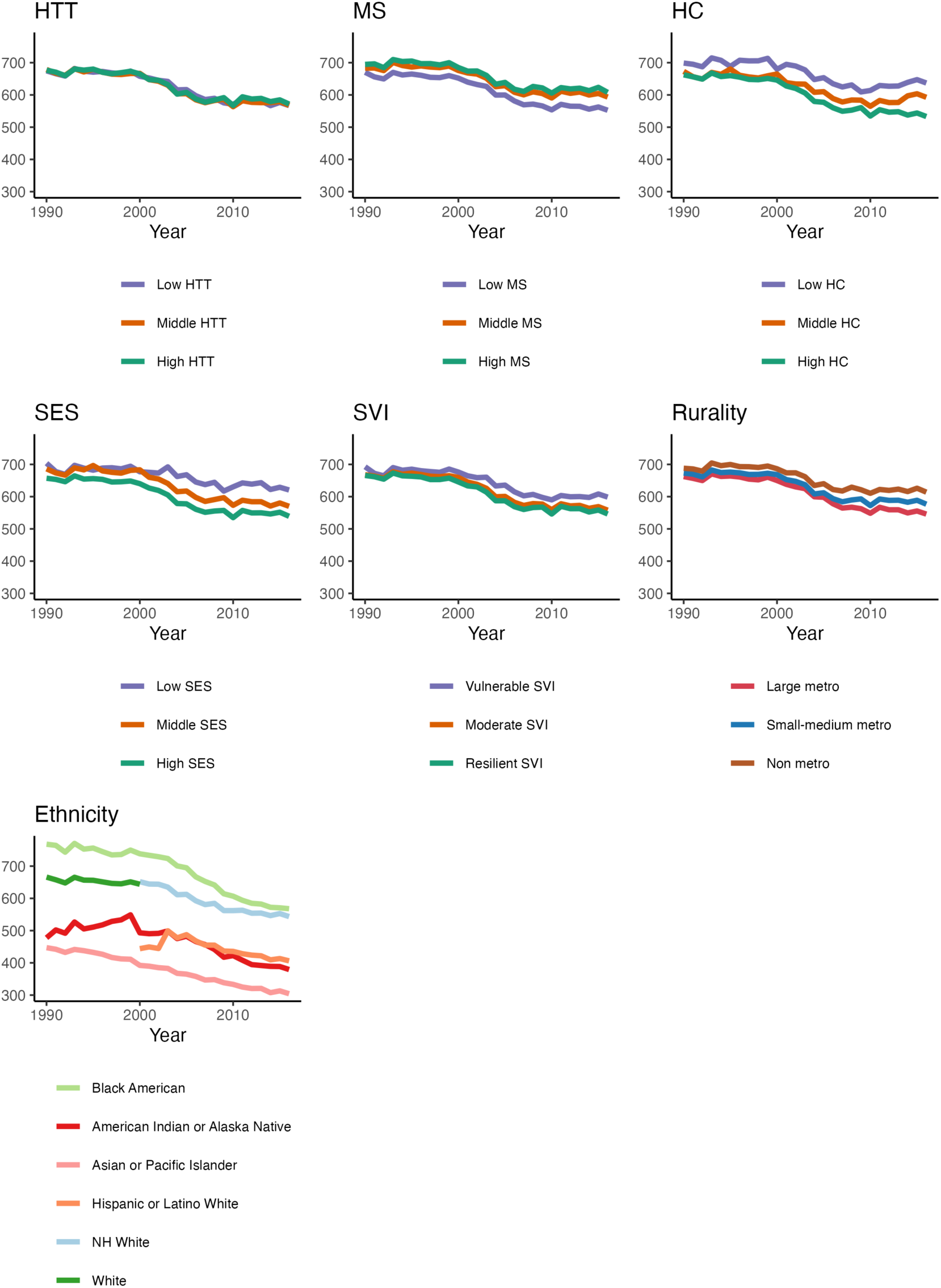
Age-adjusted mortality per 100,000 from all causes by racial/ethnic group, education attainment level, and rurality level for the age-group 65+ years. Abbreviations: NH=Non-Hispanic, SES = Socioeconomic Status, HC = Household Characteristics, MS = Minority Status, HTT = Housing Type & Transportation, SVI = Social Vulnerability Index.

**Fig. S 36.**
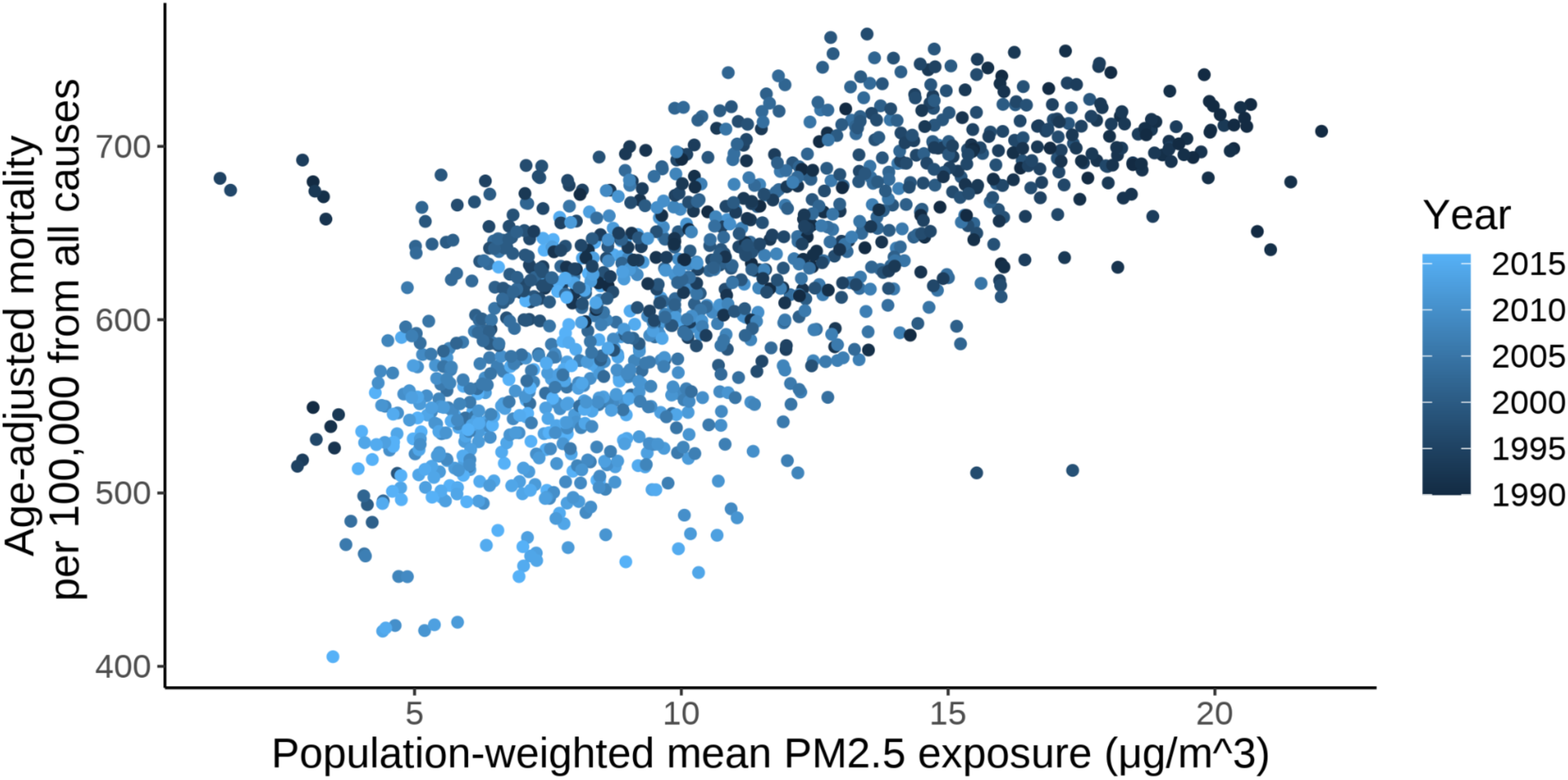
Age-adjusted mortality per 100,000 from all causes and population-weighted average PM_2.5_ exposure for the population aged 65+ years in US states and District of Columbia. Each dot corresponds to a year-state combination.

**Fig. S 37.**
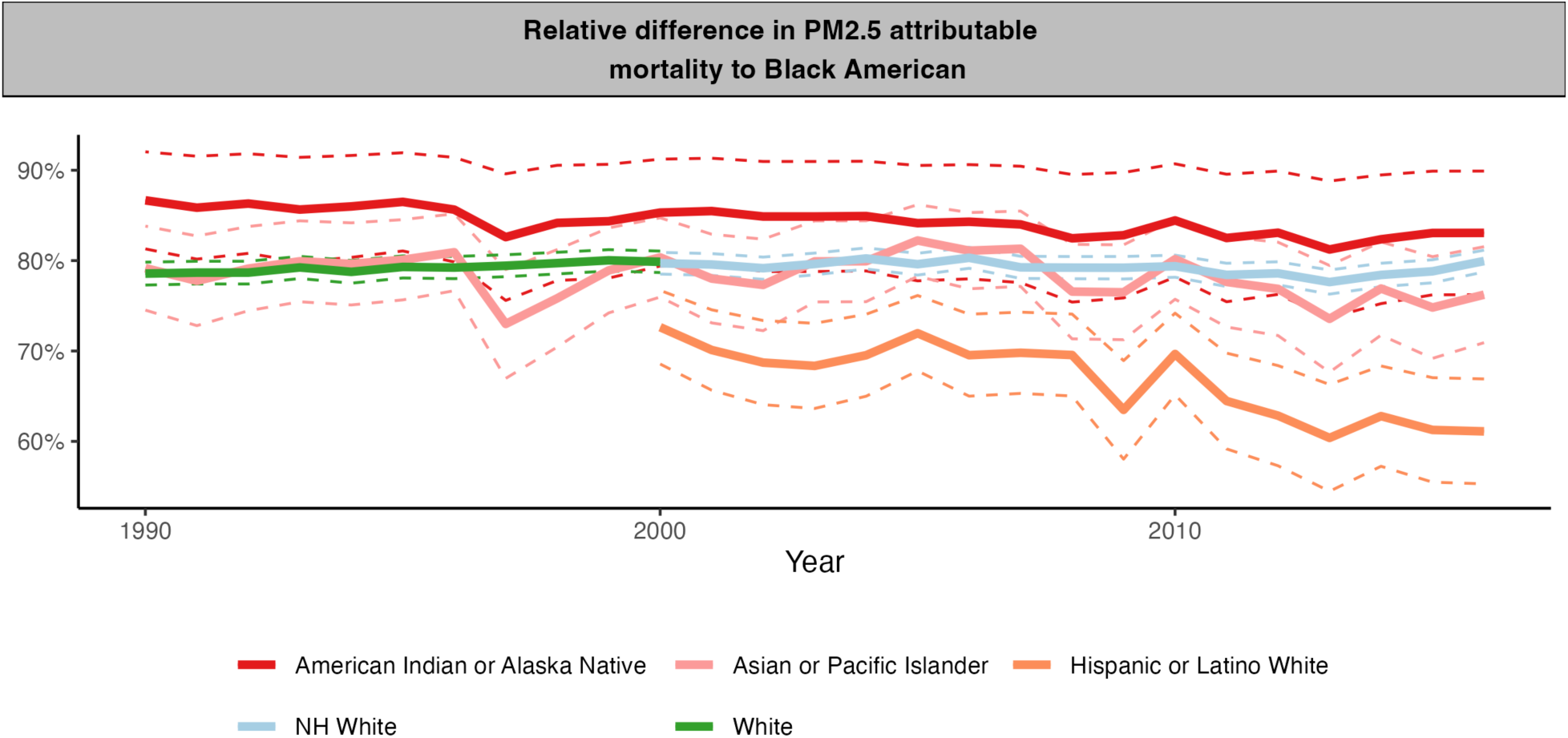
Percent at which the PM_2.5_-attributable mortality rate per 100,000 is lower for each racial/ethnic group relative to Black Americans for the age group 65+ years. This figure is for the age group 65+ years and spans the period 1990 to 2016. Abbreviations: NH=Non-Hispanic.

**Fig. S 38.**
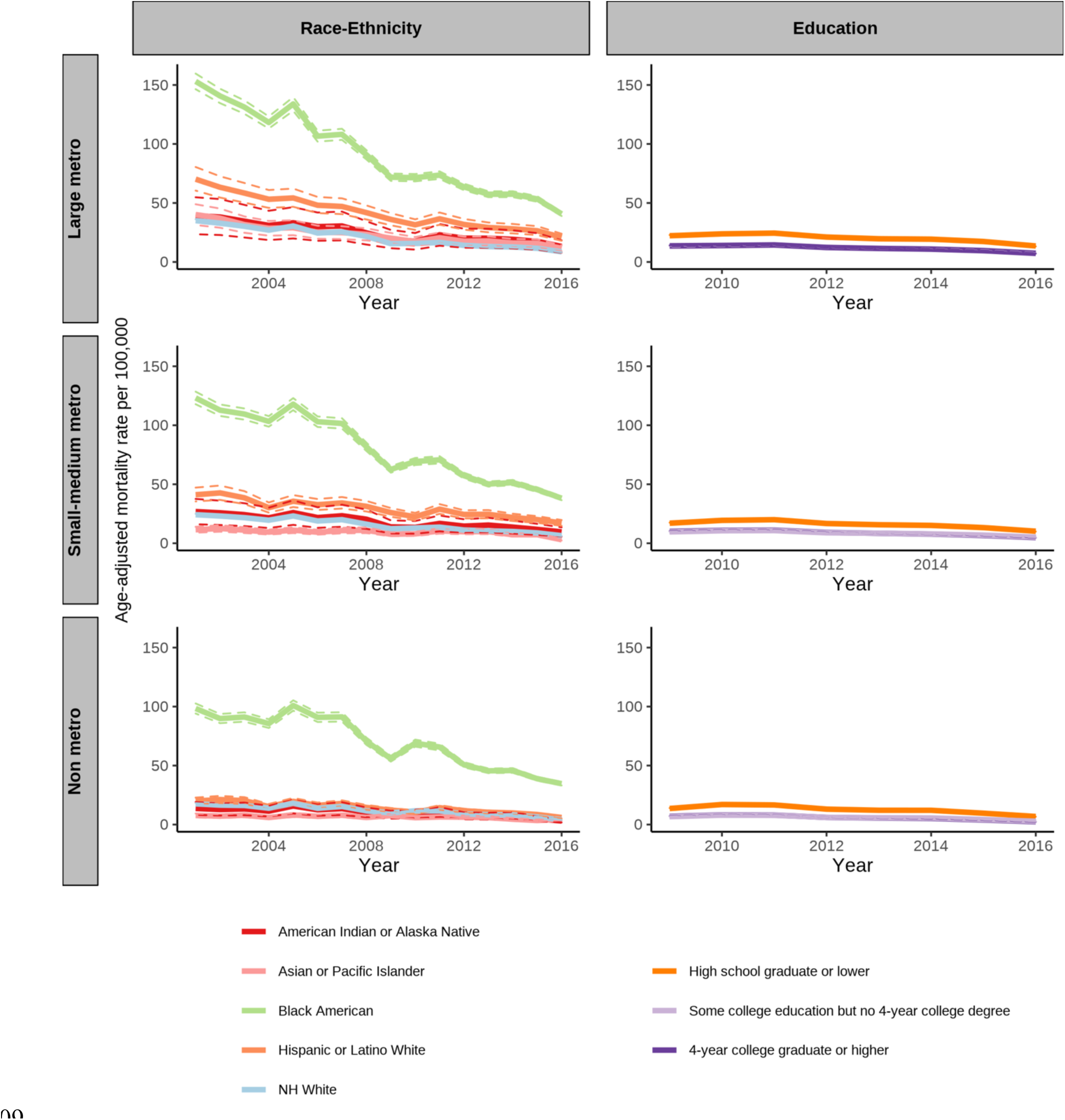
Age-adjusted PM_2.5_-attributable mortality rate for the age group 65+ years in the USA. The left column is by racial/ethnic group and rurality level. The right column is by education and rurality level. Abbreviations: NH=Non-Hispanic.

**Fig. S 39.**
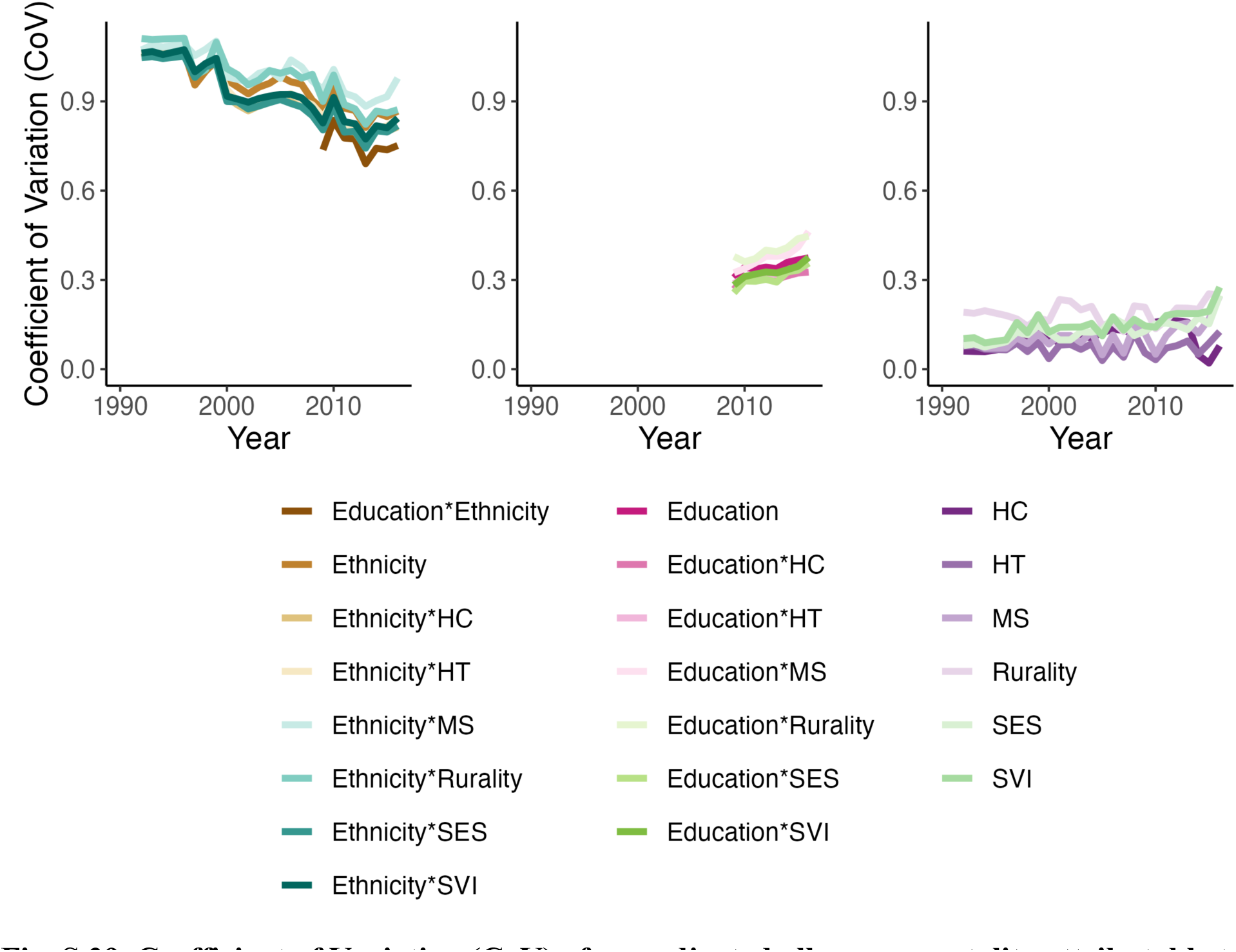
Coefficient of Variation (CoV) of age-adjusted all-cause mortality attributable to PM_2.5_ when stratifying by different sociodemographic groups for the age group 65+ years. The line for “race-ethnicity*education” corresponds to the CoV between the 15 racial/ethnic and education group combinations available. The CoV is the standard deviation of the age-adjusted PM2.5-attributable mortality rate divided by the mean age-adjusted PM2.5-attributable mortality rate. Abbreviations: NH=Non-Hispanic, SES = Socioeconomic Status, HC = Household Characteristics, MS = Minority Status, HTT = Housing Type & Transportation, SVI = Social Vulnerability Index.

**Fig. S 40.**
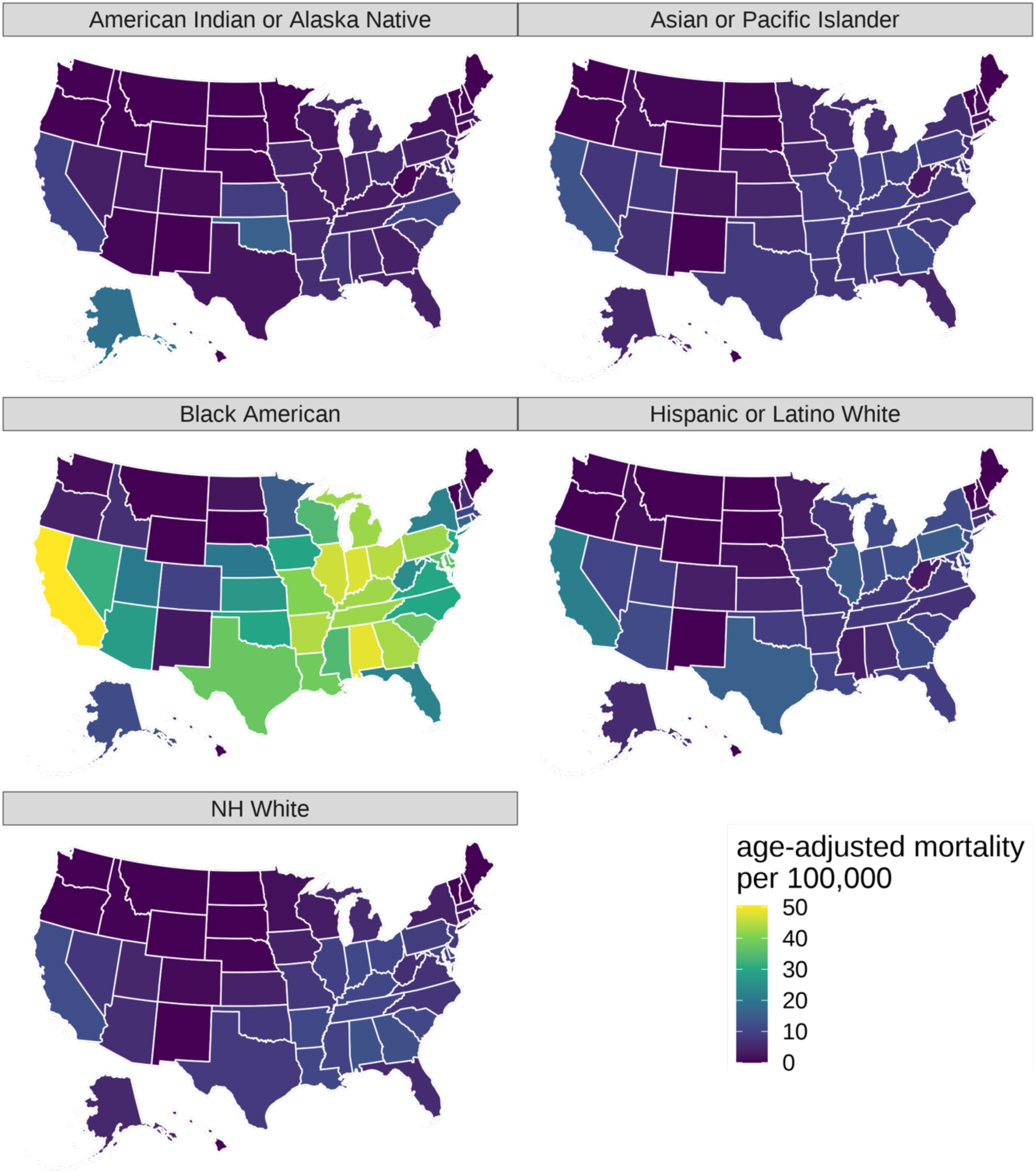
Age-adjusted PM_2.5_-attributable mortality rate for the age group 65+ years in the year 2016, by state and racial/ethnic group. Abbreviations: NH=Non-Hispanic.

**Fig. S 41.**
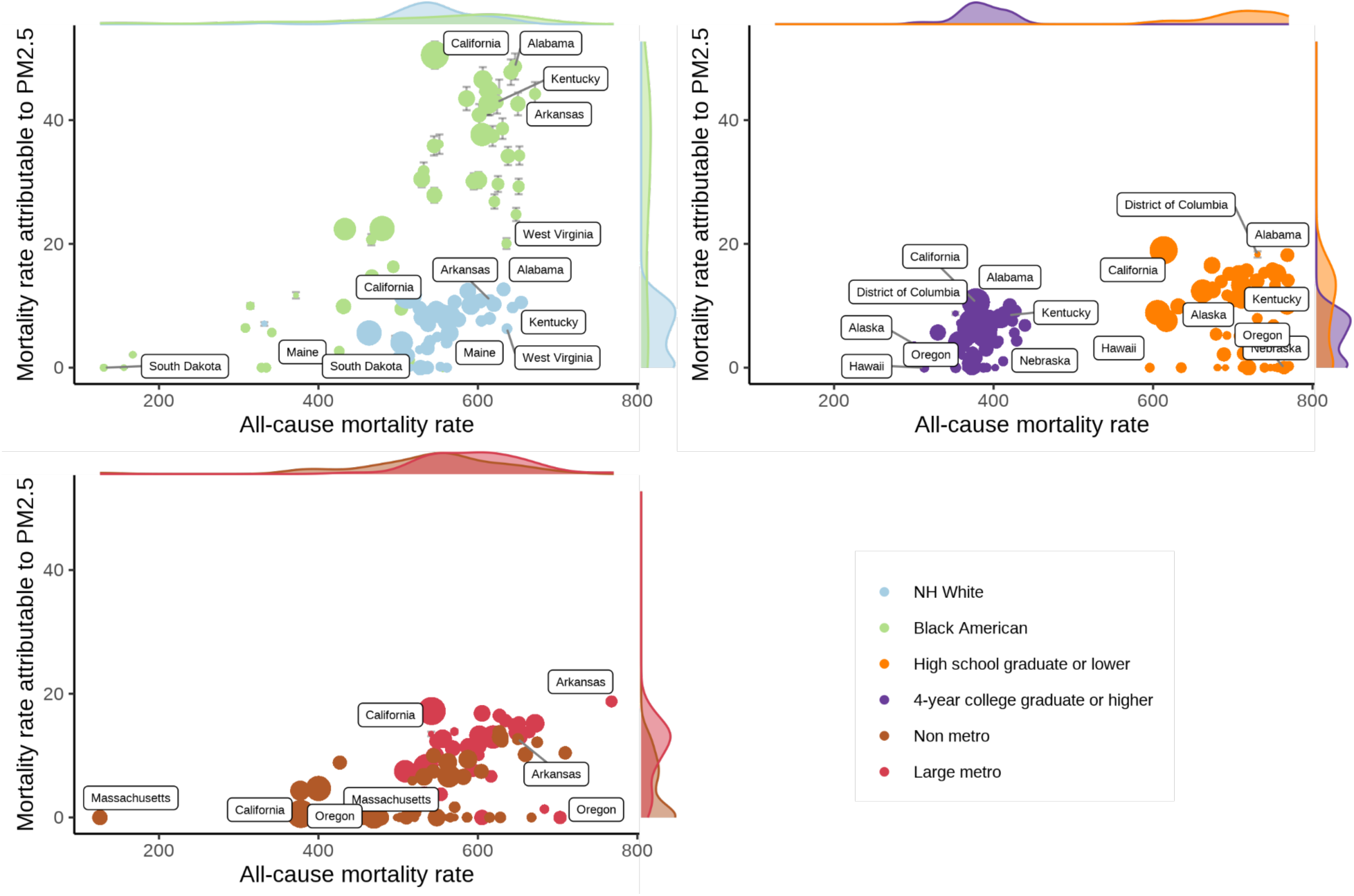
Age-adjusted mortality per 100,000 from all causes and attributable to PM_2.5_ (among those aged 65+ years) in US states in 2016 for the racial/ethnic groups “Black American” and “NH White”, low and high educational attainment, and rurality levels “Non-metro” and “large metro”. The icon area for states is proportional to the state’s population size. The most extreme points are labeled. Shaded areas are marginal kernel density estimates. Error bars denote 95% confidence intervals. Abbreviations: NH=Non-Hispanic.

**Fig. S 42.**
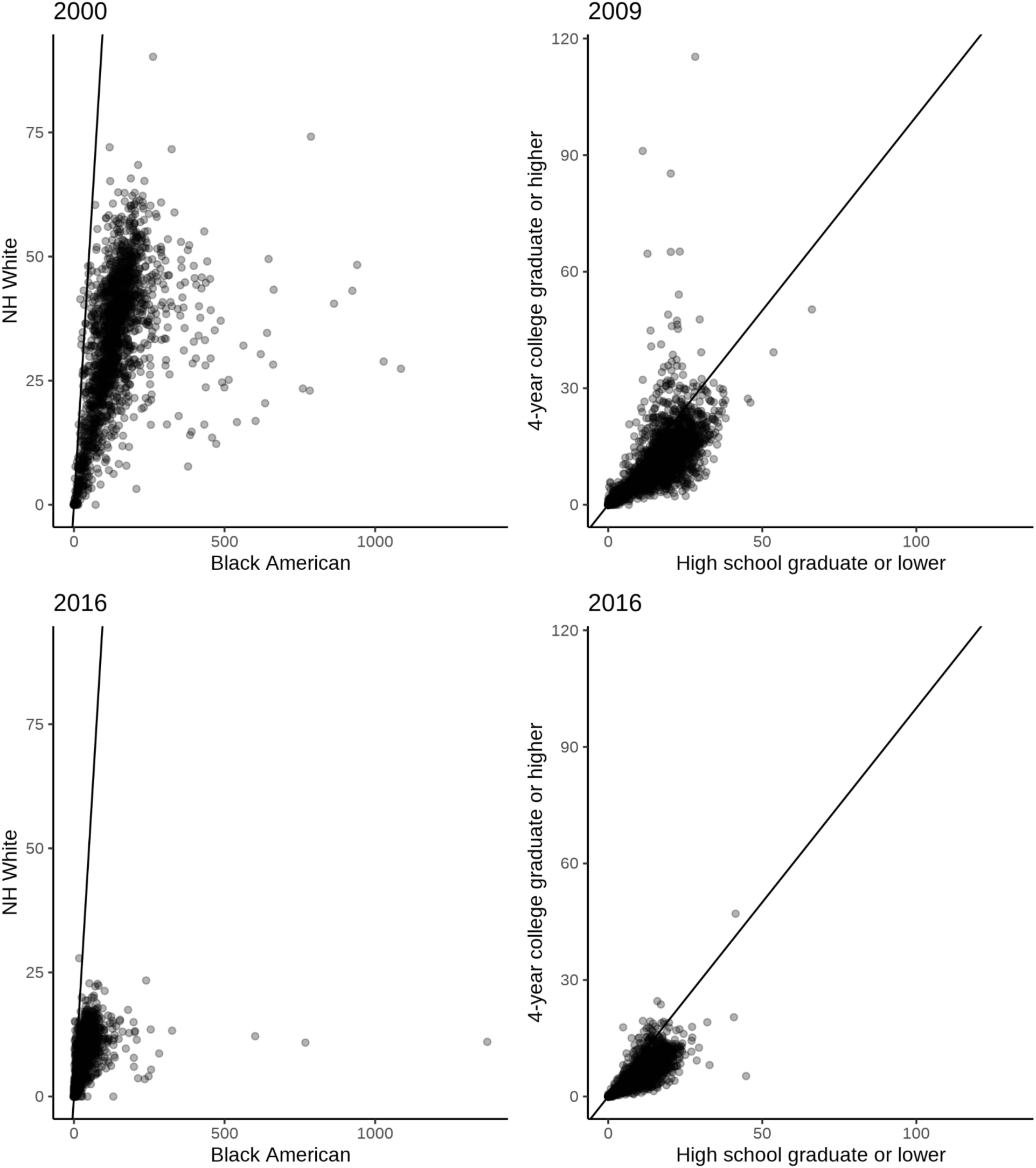
County-level scatter plot of the age-adjusted mortality per 100,000 attributable to PM_2.5_ for selected racial/ethnic groups and education level comparisons for age group 65+ years. These estimates are for the population aged 65+ years in the USA. Each point represents one county. Points on the straight line imply that the age-adjusted PM2.5-attributable mortality rate was the same for the group denoted on the x-axis as for the group denoted on the y-axis. Points below the straight line imply that the age-adjusted PM2.5-attributable mortality rate for the group on the x-axis was higher than for the group on the y-axis. Abbreviations: NH=Non-Hispanic.

**Fig. S 43.**
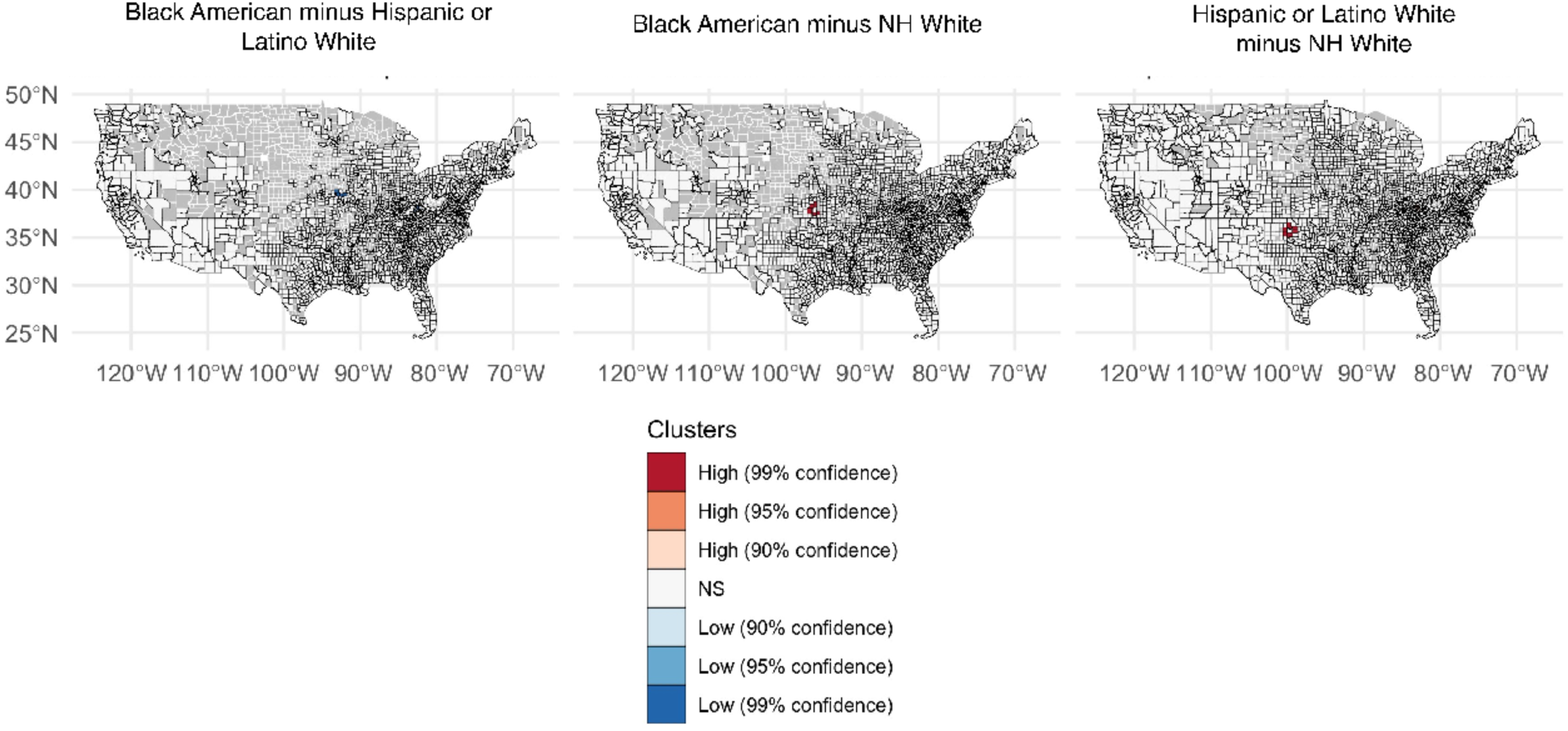
County-level spatial clusters with high (red colors) and low (blue colors) differences in the age-adjusted PM_2.5_-attributable mortality rate between racial/ethnic groups for the age group 65+ years. These maps were created for the (unweighted) mean across the period 2000 to 2016. Clusters were estimated with Local Spatial Autocorrelation as Local Getis-Ord Gi* (see “Spatial analyses” in the Materials and Methods section). Abbreviations: NH=Non-Hispanic.

**Fig. S 44.**
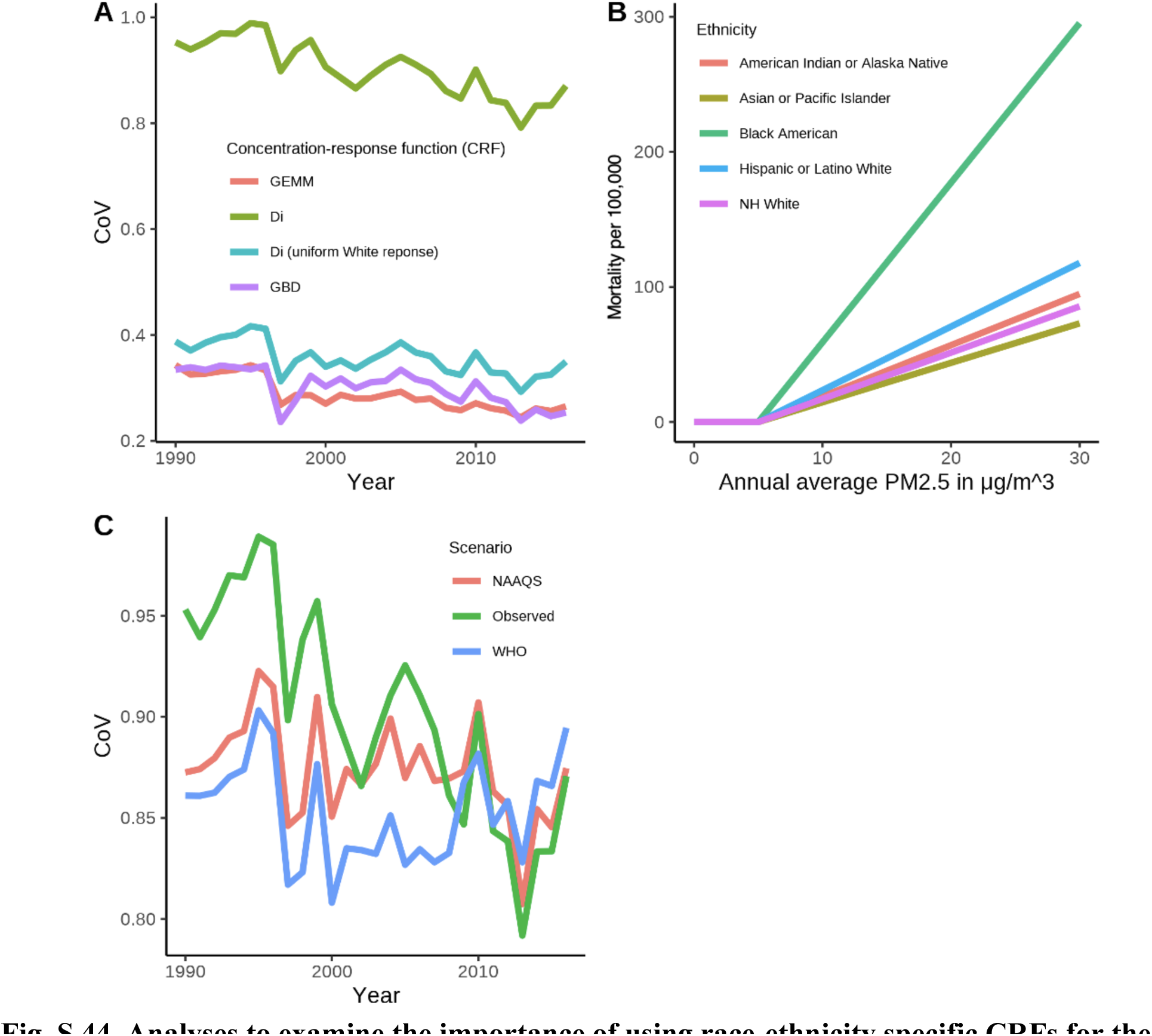
Analyses to examine the importance of using race-ethnicity specific CRFs for the age group 65+ years. (A) is the Coefficient of Variation (CoV) of age-adjusted PM2.5-attributable mortality per 100,000 across raceethnicities for different CRFs. The CoV is the standard deviation of the age-adjusted PM2.5-attributable mortality rate divided by the mean age-adjusted PM2.5-attributable mortality rate. (B) is the age-adjusted mortality per 100,000 attributable to PM2.5 in the USA by racial/ethnic group using the raceethnicity-specific CRF by Di et al.^15^, but assuming the same exposure (the population-weighted mean across the US population) to PM2.5 for all racial/ethnic groups. (C) is the Coefficient of Variation (CoV) of the age-adjusted PM2.5-attributable mortality rate for different stylized scenarios: the real observed PM2.5 exposure (“observed”), as well as a scenario each where all counties are compliant with the National Ambient Air Quality standards (15) set by the US Environmental Protection Agency (EPA) at 12μg/ m3 (“NAAQS”), and the guideline set by the World Health Organization (WHO) at 10μg/m3 (“WHO”). Abbreviations: CRF=concentration response function; NH=Non-Hispanic.

**Fig. S 45.**
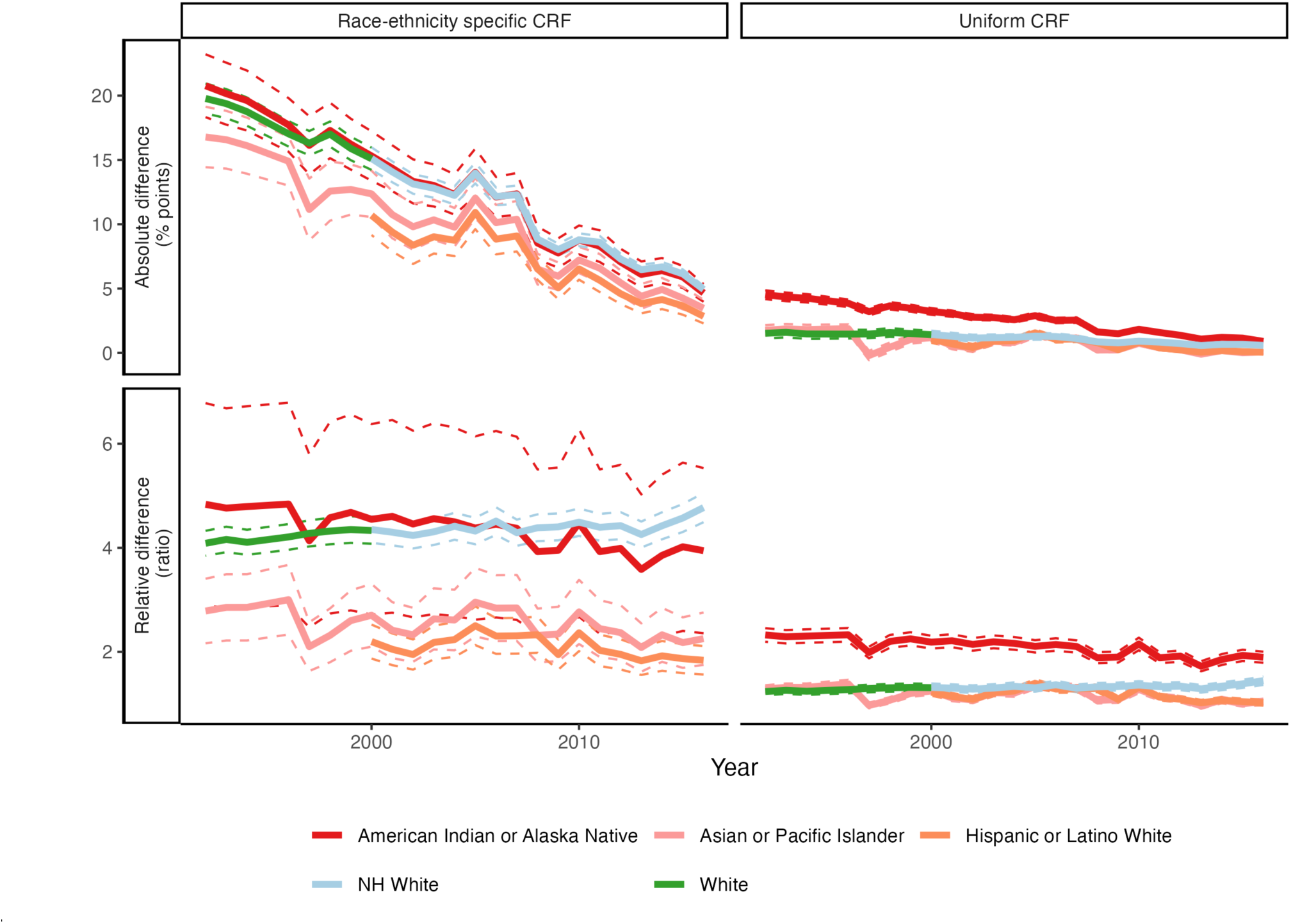
Differences in the proportion of all-cause mortality that was attributable to PM_2.5_ for the age group 65+ years between Black Americans and other racial/ethnic groups. The first row is the *absolute difference* (in percentage points) of the proportion of the age-adjusted all-cause mortality that was attributable to PM2.5 between Black Americans and each racial/ethnic group. Positive values indicate that Black Americans had a higher proportion of all-cause mortality attributable to PM2.5. The second row is the *ratio* (i.e., a measure of the relative difference) of the proportion of the age-adjusted all-cause mortality that was attributable to PM2.5 for Black Americans divided by the proportion of the age-adjusted all-cause mortality that was attributable to PM2.5 for each racial/ethnic group. The first column assumes the race-ethnicity-specific CRF from Di. et al.^15^. The second column assumes a uniform CRF for all racial/ethnic groups by applying the CRF for “NH White” from Di et al. to all racial/ethnic groups. Abbreviations: CRF=concentration response function; NH=Non-Hispanic.

**Fig. S 46.**
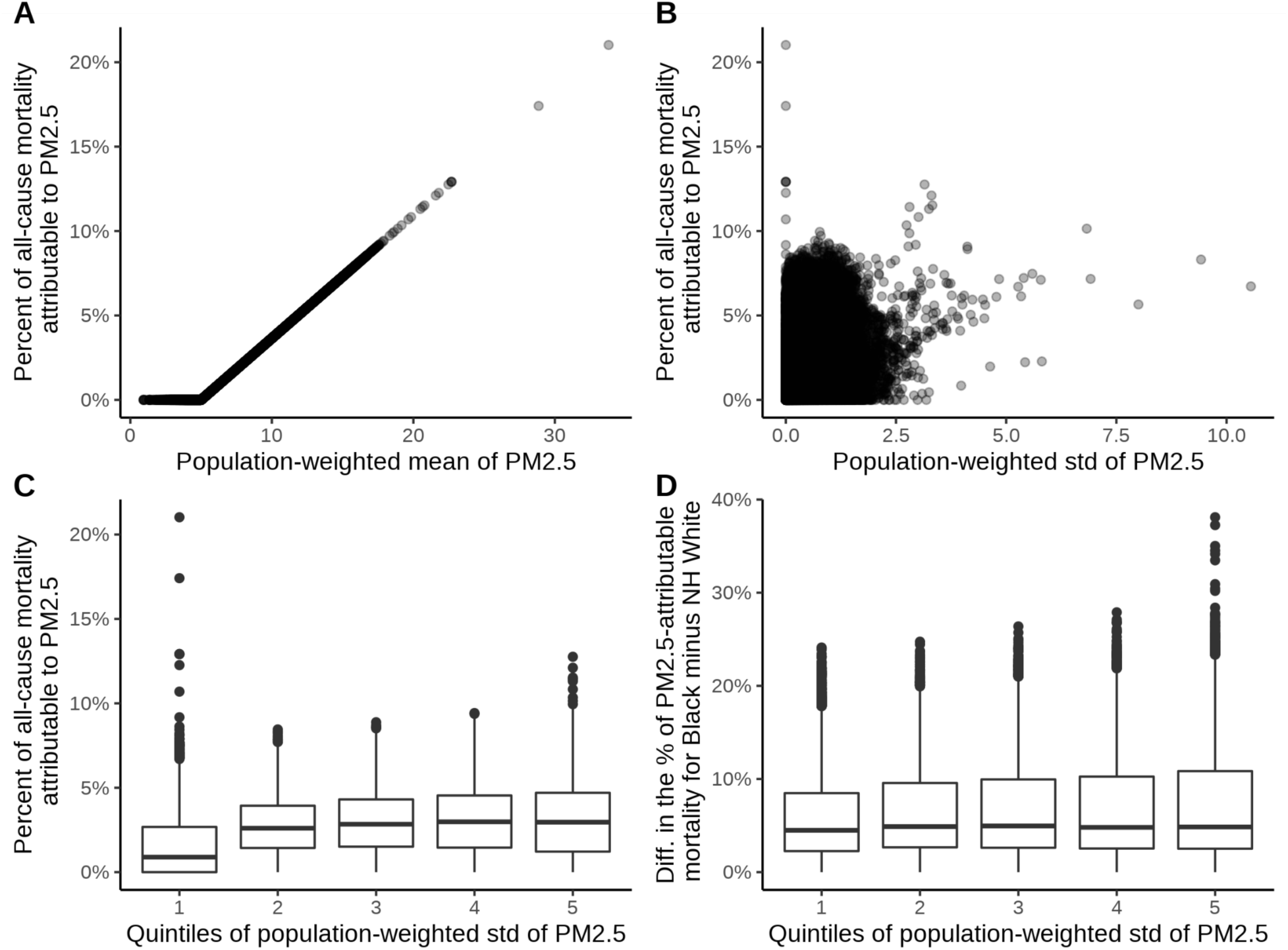
Exploring the link between PM_2.5_-attributable mortality at the county level and the variation of the PM_2.5_ estimates aggregated from the census tract to county level. Panel (**A**) displays a plot with the x-axis showing the mean population-weighted PM2.5 exposure in each county-year combination, and the y-axis depicting the percentage of the age-adjusted all-cause mortality rate that was attributable to PM2.5. Panel (**B**) plots the within-county-year standard deviation in PM2.5 exposure at the census-tract level (x-axis) against the county-level percentage of the age-adjusted all-cause mortality rate that was attributable to PM2.5 (y-axis). Outlier points are California and Alaska. Panel (**C**) shows the same data as Panel B but categorizes the x-axis values (within-county-year standard deviation in PM2.5) into quintiles. Panel (**D**) plots quintiles of the within-county-year standard deviation in PM2.5 exposure (x-axis) against the difference (in percentage points) in the percent of the age-adjusted all-cause mortality rate that was attributable to PM2.5 for Black Americans versus the NH White population (y-axis). Positive values on the y-axis indicate that Black Americans had a higher proportion of all-cause mortality attributable to PM2.5 exposure than the NH White population. Abbreviations: Diff.=difference; NH=Non-Hispanic; std=standard deviation

**Fig. S 47.**
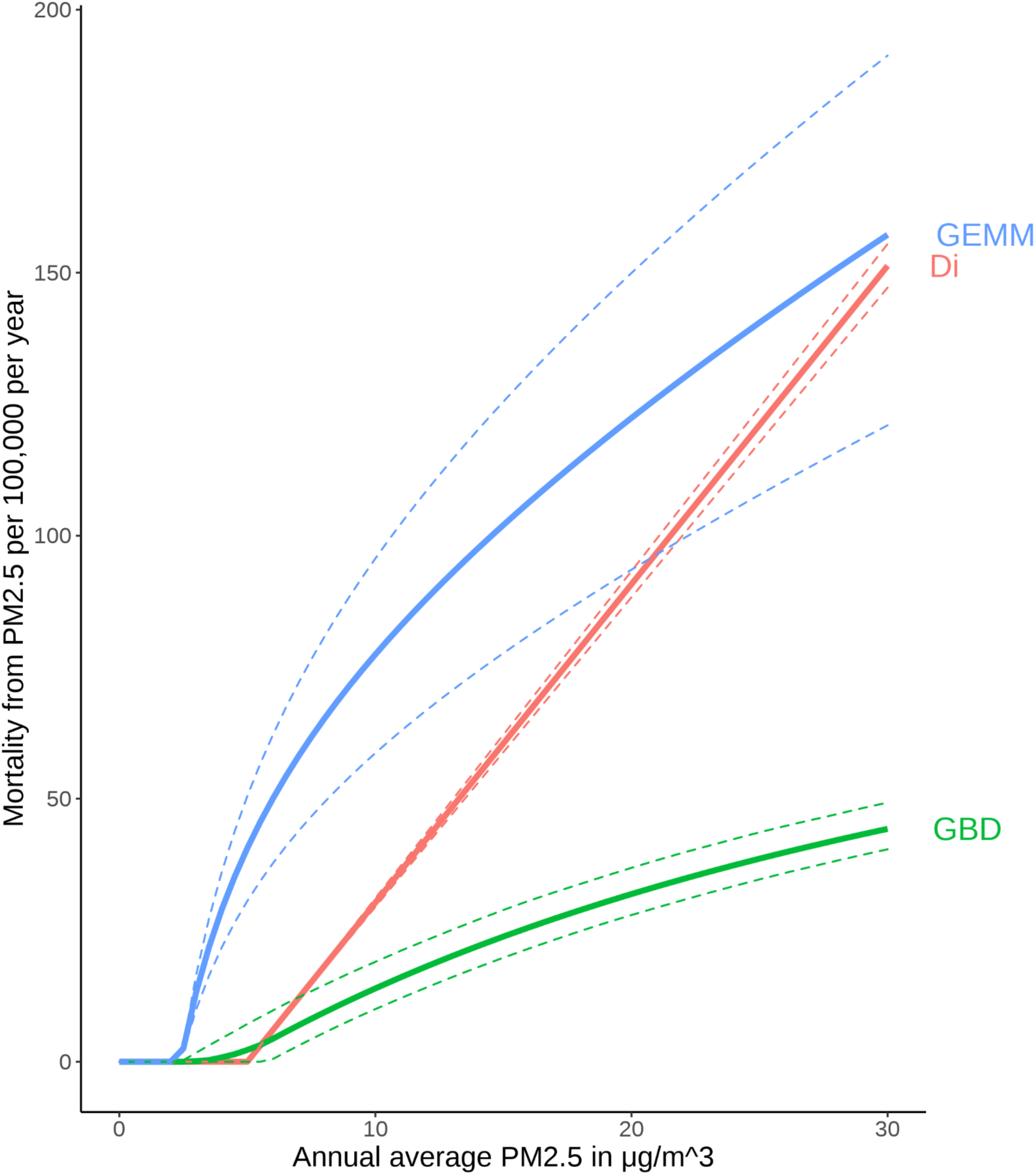
Shape of the CRFs by Di et al. (“Di”), the Global Burden of Disease project (“GBD”), and Burnett et al. (“GEMM”) The dashed lines depict 95% confidence intervals. The baseline mortality rate was taken from the age group 25+ years in the USA in 2016. The y-axis is given in crude (not age-adjusted) mortality per 100,000. GBD^16^ uses ischemic heart disease, ischemic stroke, intracerebral hemorrhage, subarachnoid hemorrhage, lung cancer, chronic obstructive pulmonary disease, lower respiratory infection, and diabetes mellitus type 2 as baseline mortality. GEMM^11^ is based on all non-communicable diseases and lower respiratory infections. Di et al.^15^ is based on all-cause mortality. Abbreviations: CRF=concentration-response function.

**Fig. S 48.**
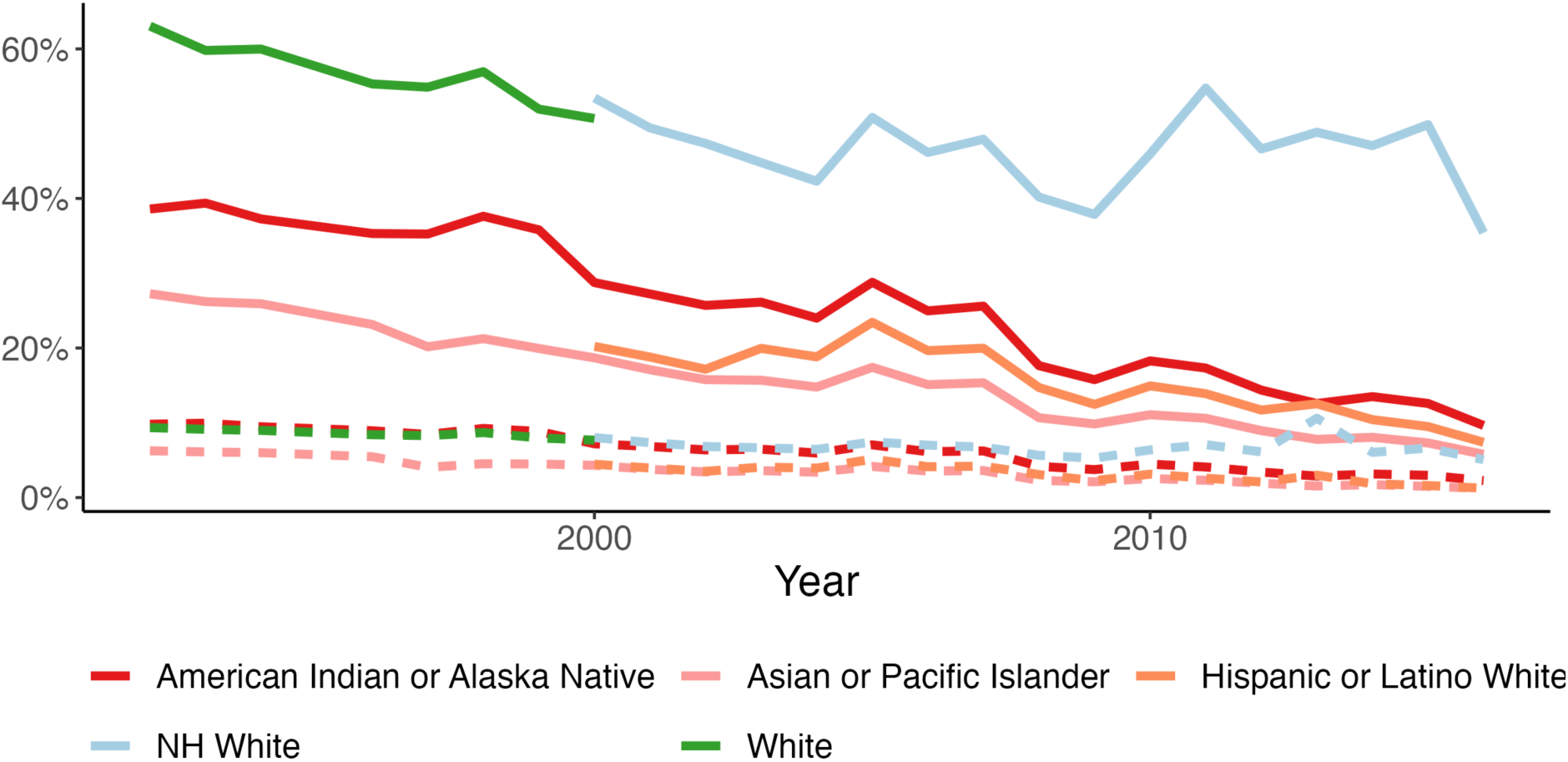
Percent of the difference in the age-adjusted mortality rate between each race-ethnicity and “Black American” that is attributable to PM_2.5_, using either race-ethnicity-specific CRFs (solid lines) or a uniform CRF (dashed lines). The solid lines are the results calculated using race-ethnicity-specific CRFs from Di et al.^15^. The dashed lines are the results calculated when applying the CRF in Di et al.^15^ for “NH White” to all race-ethnicities. Abbreviations: CRF=Concentration-response function; NH=Non-Hispanic.

**Fig. S 49.**
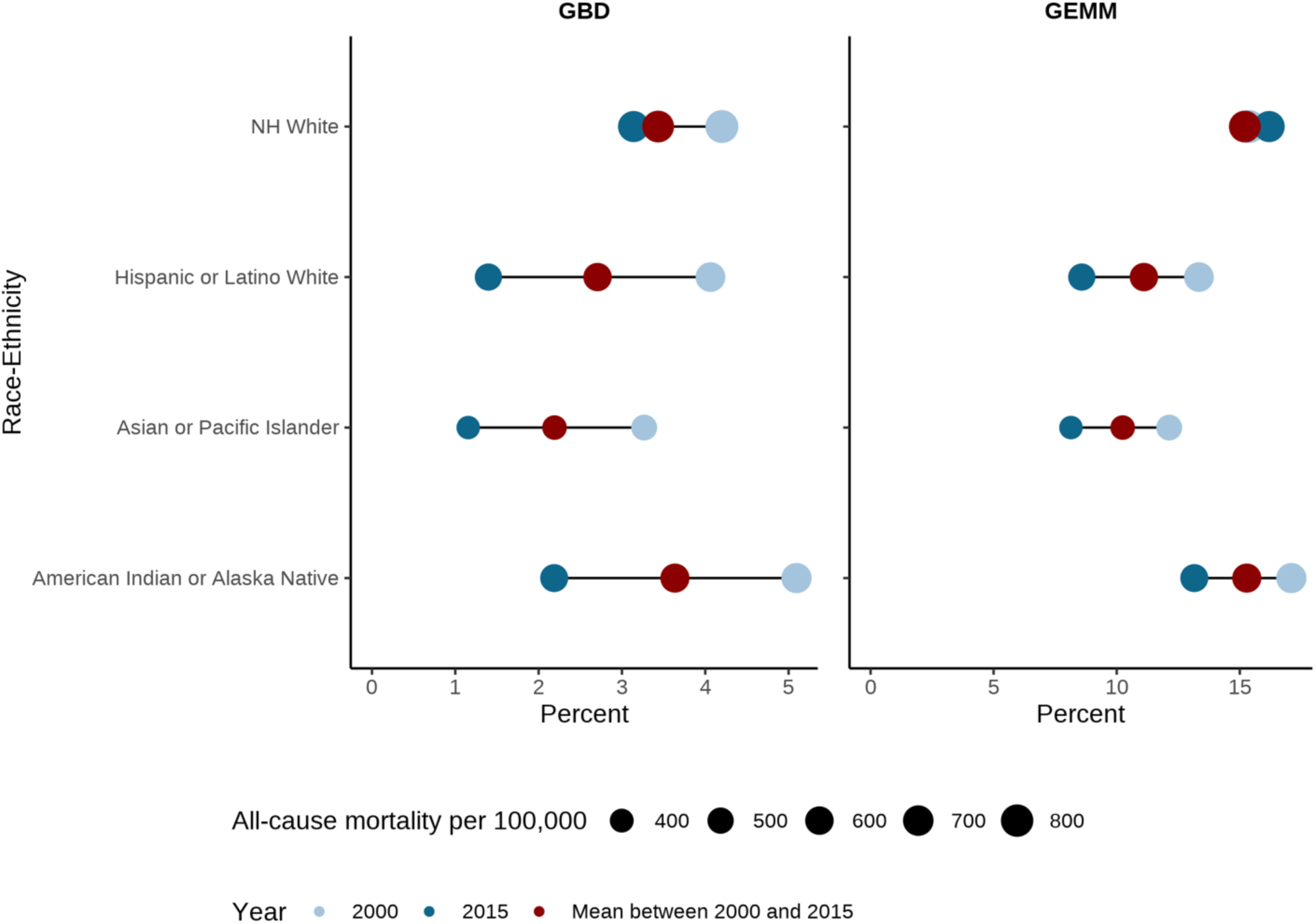
Extent to which the difference in the age-adjusted mortality rate between each racial/ethnic group and Black Americans can be attributed to PM_2.5_. Of the racial/ethnic groups considered in this study, Black Americans have both the highest all-cause mortality rate and highest PM2.5-attributable mortality rate. Both columns are the percent of the difference in all-cause mortality between each race-ethnicity and Black Americans that can be attributed to PM2.5 exposure. The left colum is GBD ^16^, the right column is GEMM^11^. Dark blue dots denote 2015 and light blue dots denote 2000. Red dots denote the unweighted mean from 2000 to 2015. The dot sizes are proportional to the age-adjusted all-cause mortality rate that is attributable to PM2.5. Abbreviations: NH=Non-Hispanic

**Fig. S 50.**
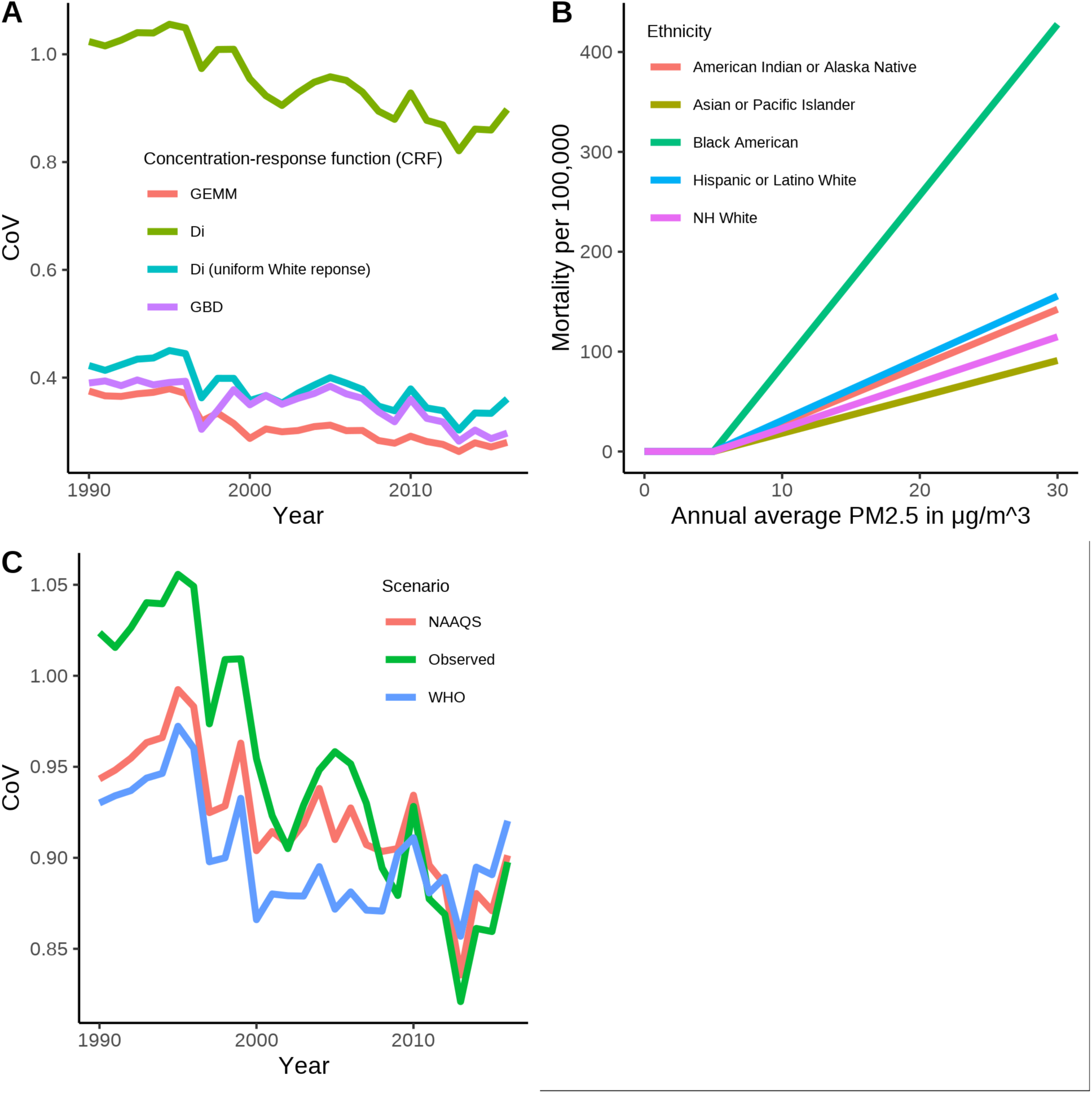
Analyses to examine the importance of using race-ethnicity specific CRFs for the age group 25+ years. (A) is the Coefficient of Variation (CoV) of age-adjusted PM2.5-attributable mortality per 100,000 across racial/ethnic groups for different CRFs. The CoV is the standard deviation of the age-adjusted PM2.5-attributable mortality rate divided by the mean age-adjusted PM2.5-attributable mortality rate. (B) is the age-adjusted mortality per 100,000 attributable to PM2.5 in the USA by racial/ethnic group using the race-ethnicity-specific CRF by Di et al.^15^, but assuming the same exposure (the population-weighted mean across the US population) to PM2.5 for all racial/ethnic groups. (C) is the Coefficient of Variation (CoV) of the age-adjusted PM2.5-attributable mortality rate for different stylized scenarios: the real observed PM2.5 exposure (“observed”), as well as a scenario each where all counties are compliant with the National Ambient Air Quality standards^18^ set by the US Environmental Protection Agency (EPA) at 12μg/ m^3^ (“NAAQS”), and the guideline set by the World Health Organization (WHO) at 10μg/m^3^ (“WHO”). Abbreviations: NH=Non-Hispanic, CoV = Coefficient of Variation; CRF=Concentration-response function.

**Fig. S 51.**
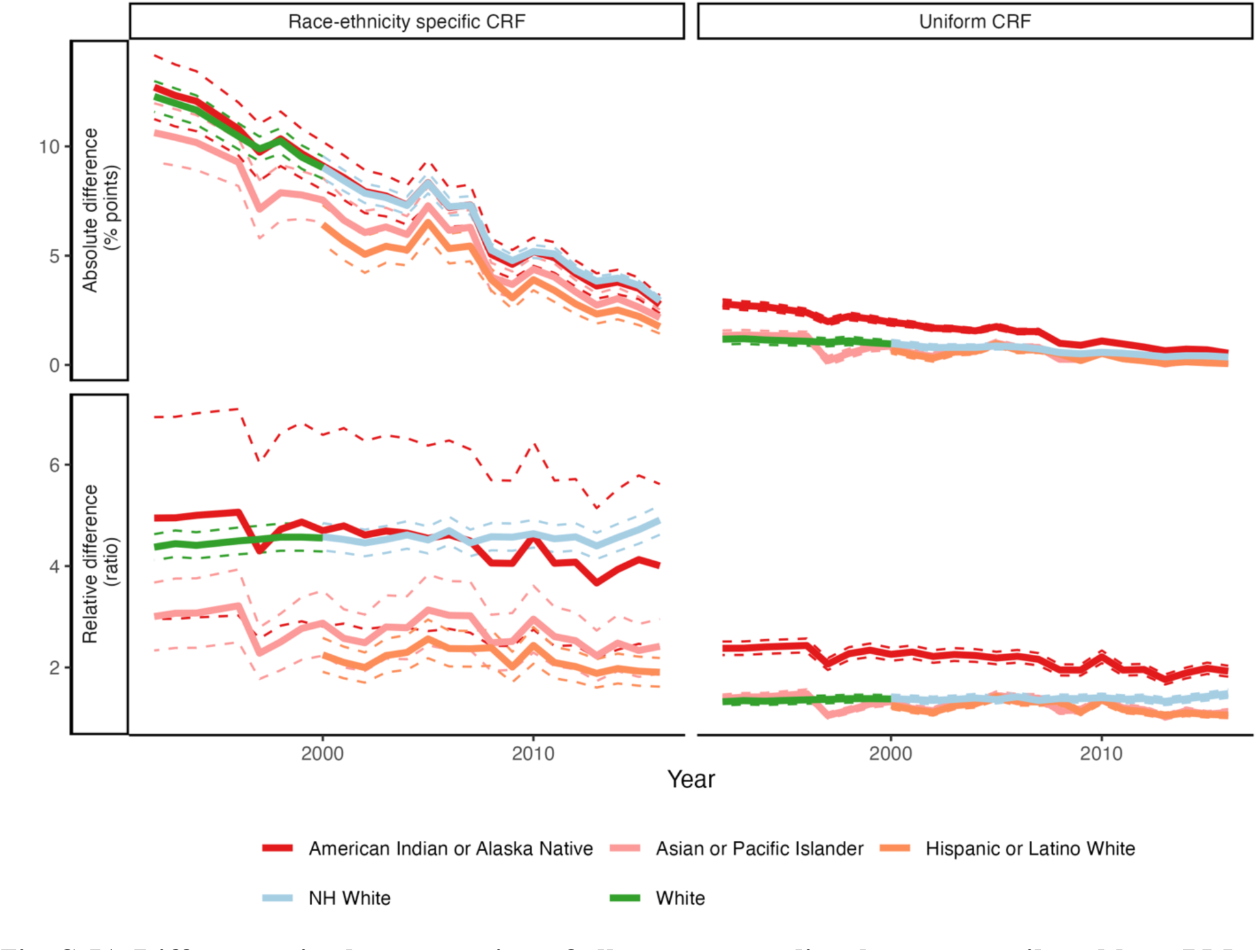
Differences in the proportion of all-cause mortality that was attributable to PM_2.5_ for the age group 25+ years between Black Americans and other racial/ethnic groups. The first row is the *absolute difference* (in percentage points) of the proportion of the age-adjusted all-cause mortality that was attributable to PM2.5 between Black Americans and each racial/ethnic group. Positive values indicate that Black Americans had a higher proportion of all-cause mortality attributable to PM2.5. The second row is the *ratio* (i.e., a measure of the relative difference) of the proportion of the age-adjusted all-cause mortality that was attributable to PM2.5 for Black Americans divided by the proportion of the age-adjusted all-cause mortality that was attributable to PM2.5 for each racial/ethnic group. The first column assumes the race-ethnicity-specific CRF from Di. et al.^15^. The second column assumes a uniform CRF for all racial/ethnic groups by applying the CRF for “NH White” from Di et al. to all racial/ethnic groups. Abbreviations: CRF = concentration response function, NH=Non-Hispanic.

**Fig. S 52.**
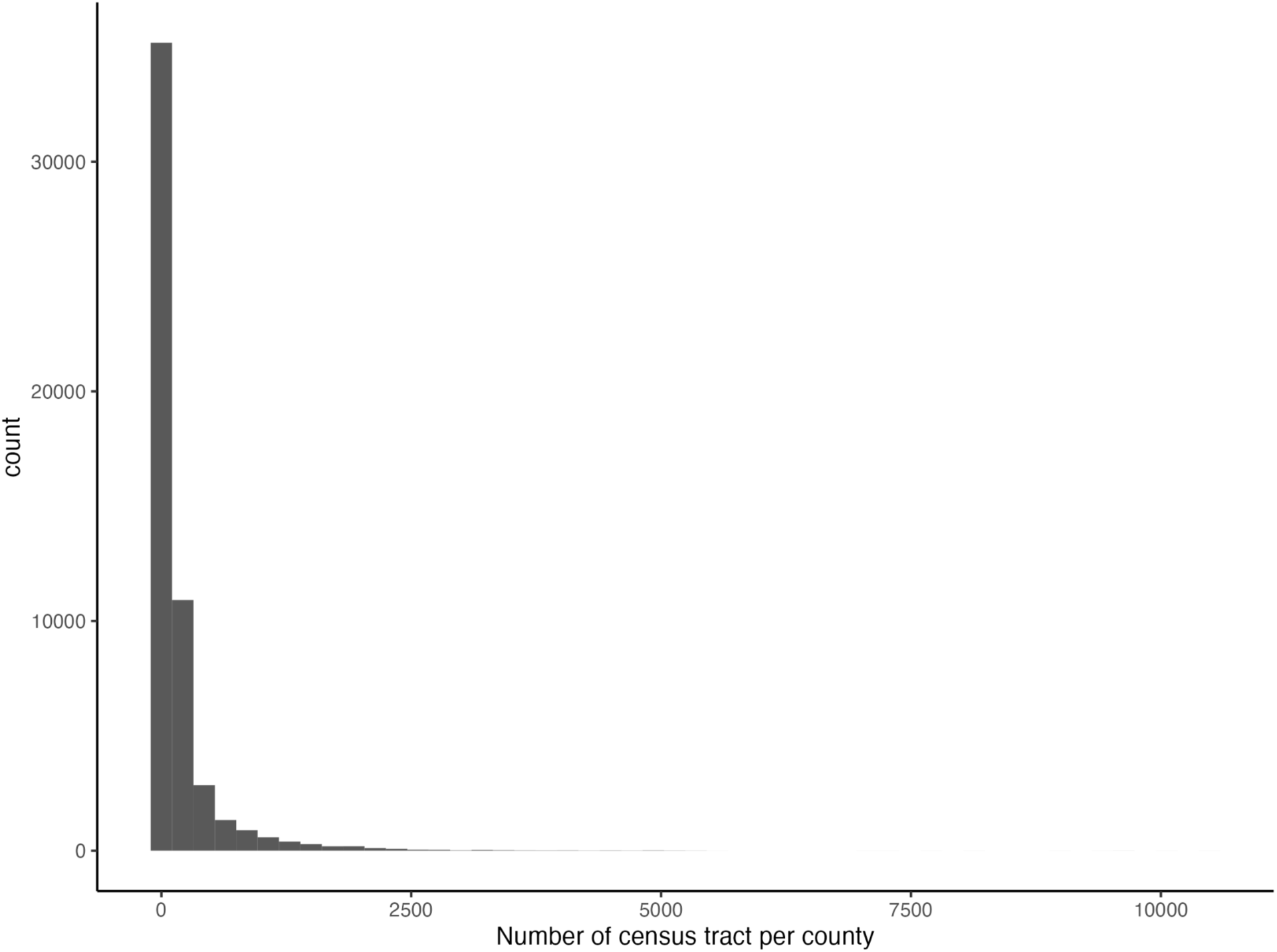
Histogram of the number of census tract per county for the period 2000 to 2016.

**Fig. S 53.**
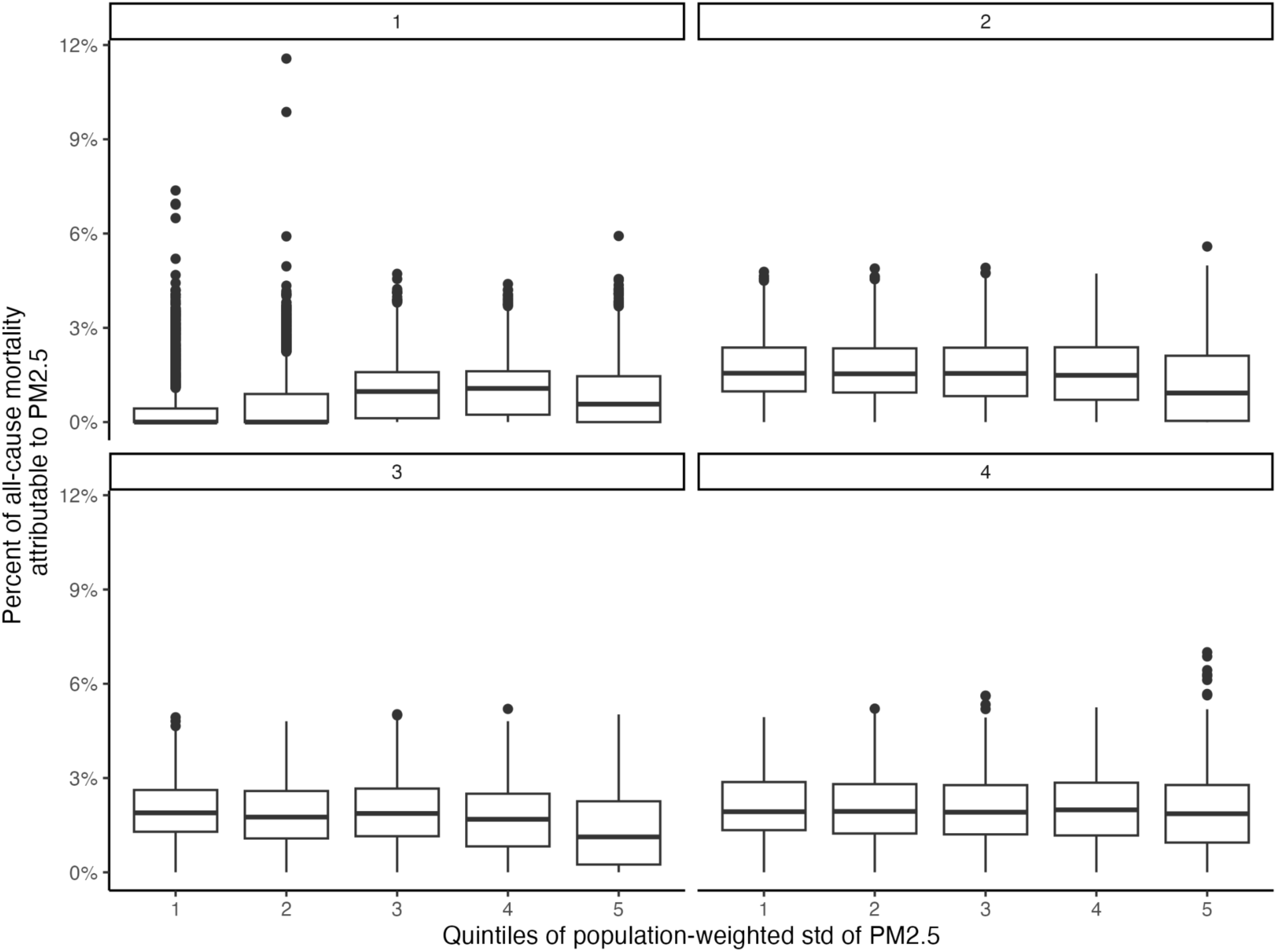
Association of the variation in census-tract level PM_2.5_ measurements with county-level PM_2.5_-attributable mortality for the period 2000 to 2016 stratified by quantiles of the number of census tracts per county.

**Fig. S 54.**
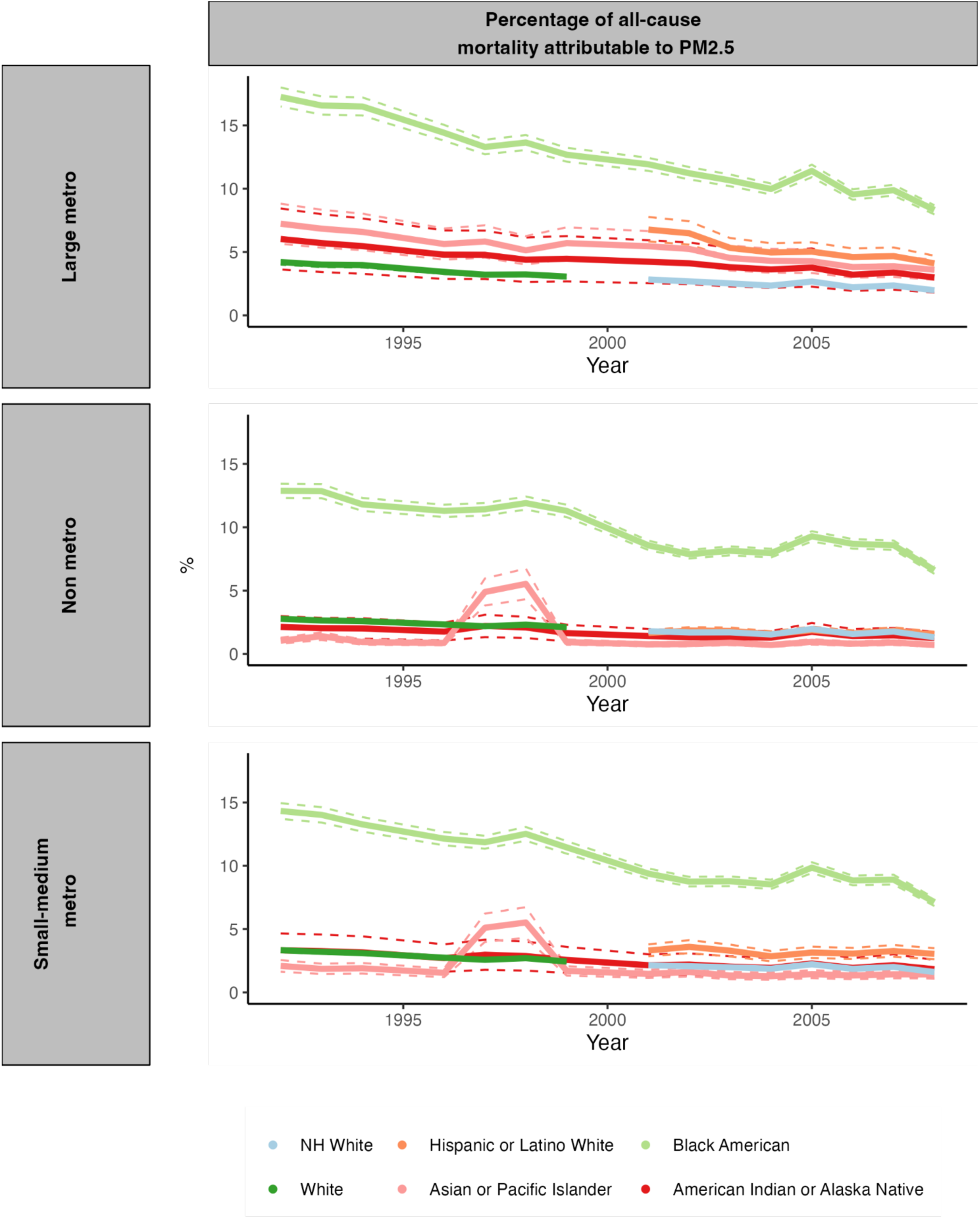
Percentage of all-cause mortality attributable to PM_2.5_ for each racial/ethnic group stratified by rurality. The dashed lines depict 95% confidence intervals. Abbreviations: NH=Non-Hispanic.

**Fig. S 55.**
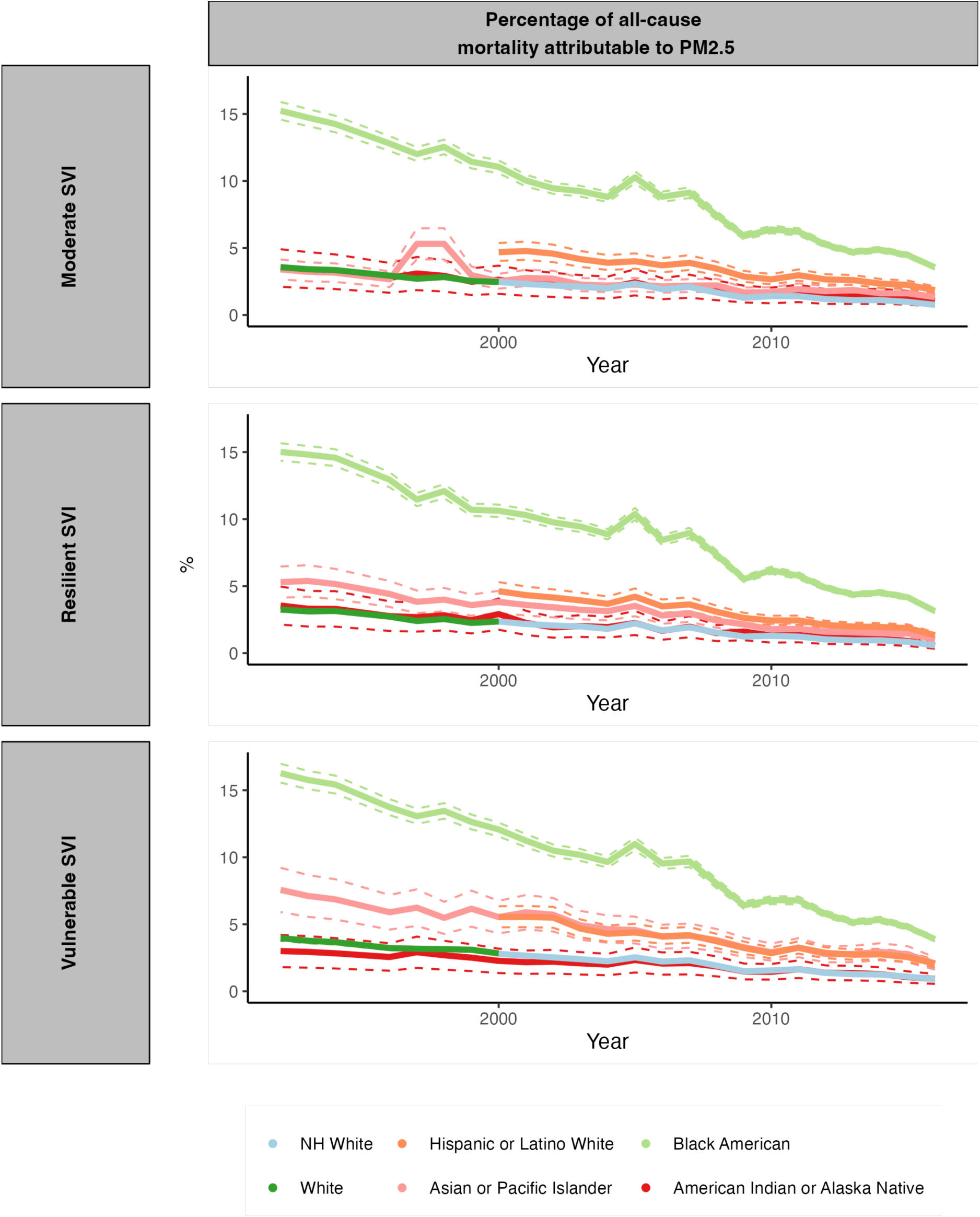
Percentage of all-cause mortality attributable to PM_2.5_ for each racial/ethnic group stratified by the social vulnerability index. The dashed lines depict 95% confidence intervals. Abbreviations: NH=Non-Hispanic.

**Fig. S 56.**
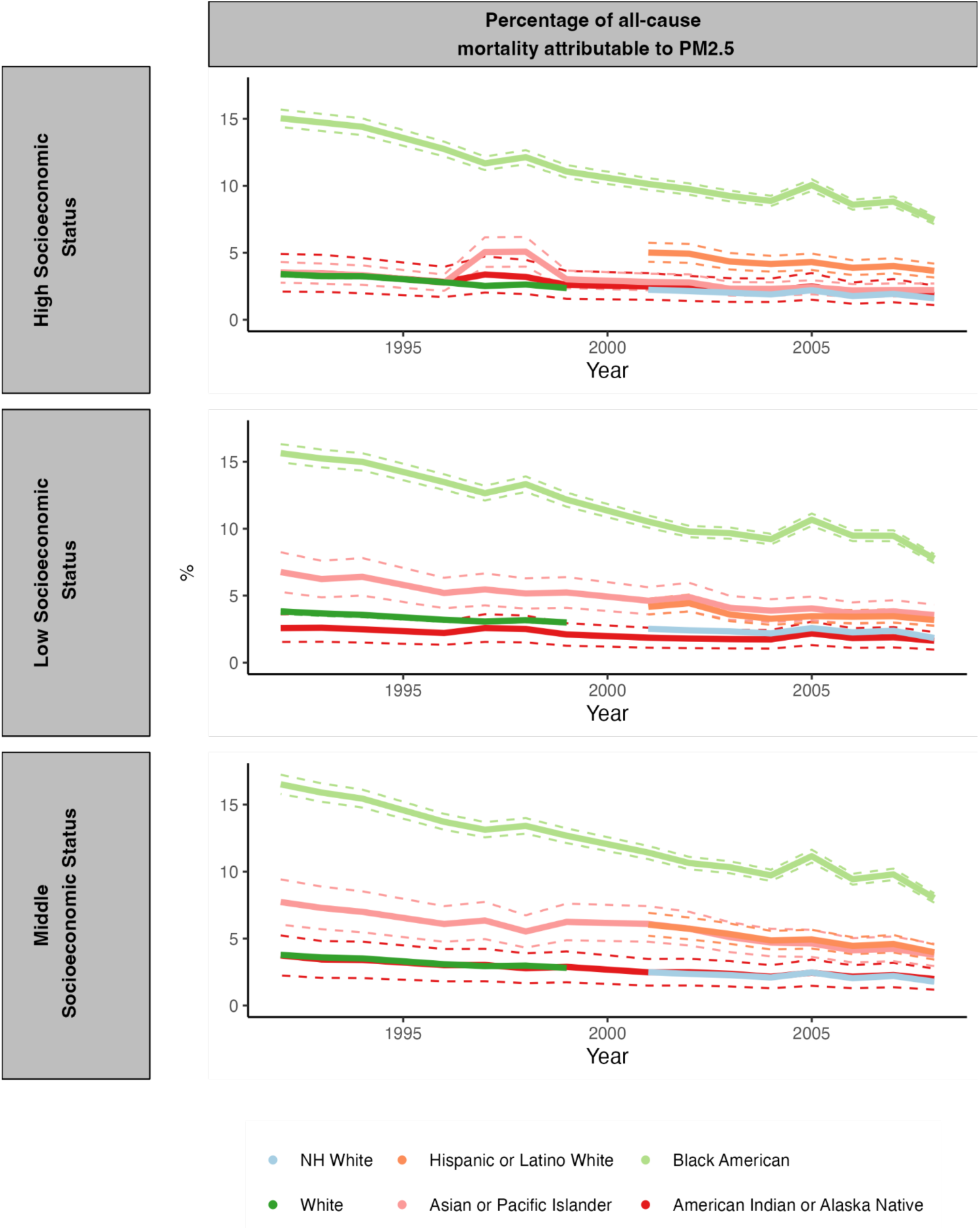
Percentage of all-cause mortality attributable to PM_2.5_ for each racial/ethnic group stratified by socioeconomic status. The dashed lines depict 95% confidence intervals. Abbreviations: NH=Non-Hispanic.

**Fig. S 57.**
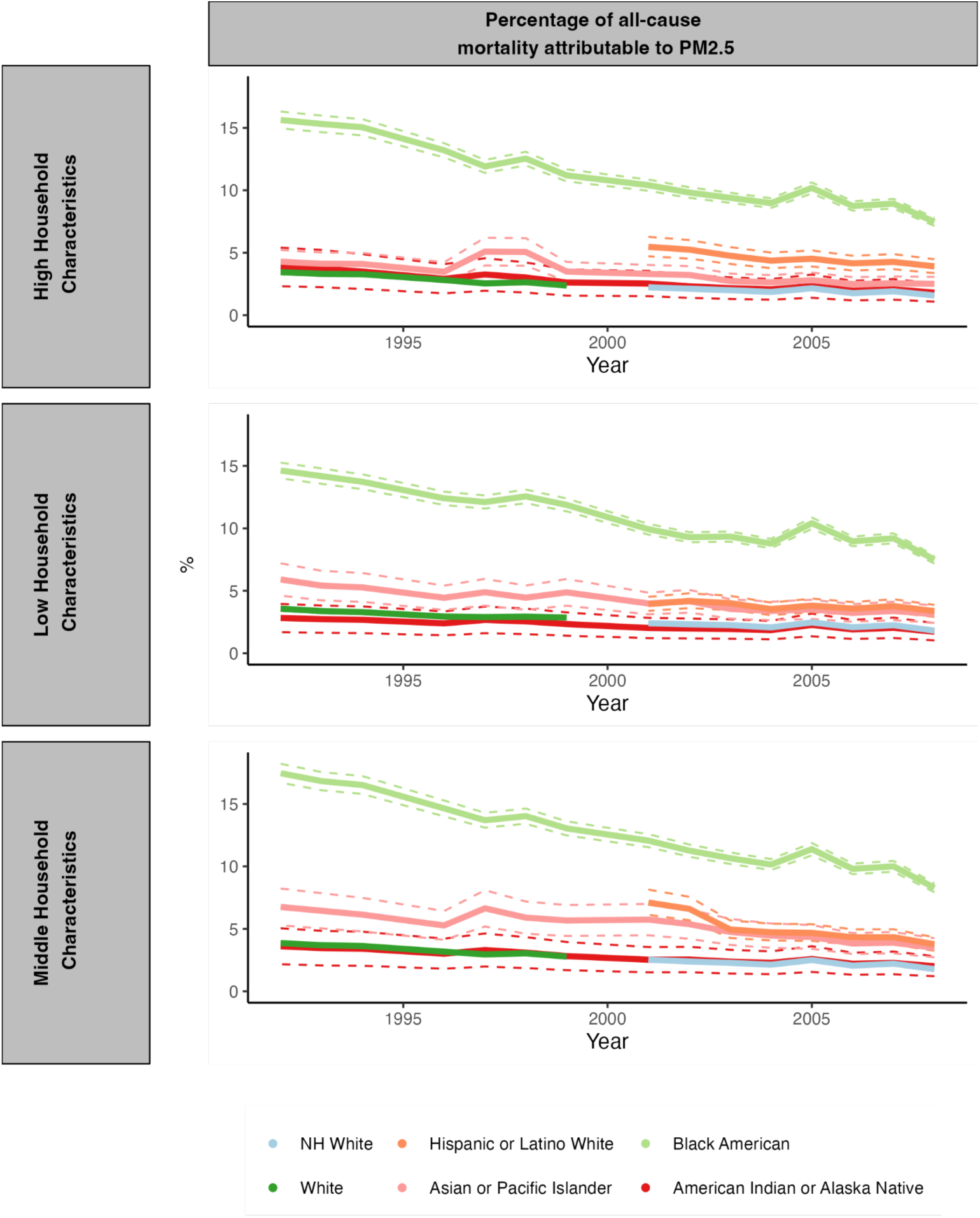
Percentage of all-cause mortality attributable to PM_2.5_ for each racial/ethnic group stratified by household characteristics. The dashed lines depict 95% confidence intervals. Abbreviations: NH=Non-Hispanic.

**Fig. S 58.**
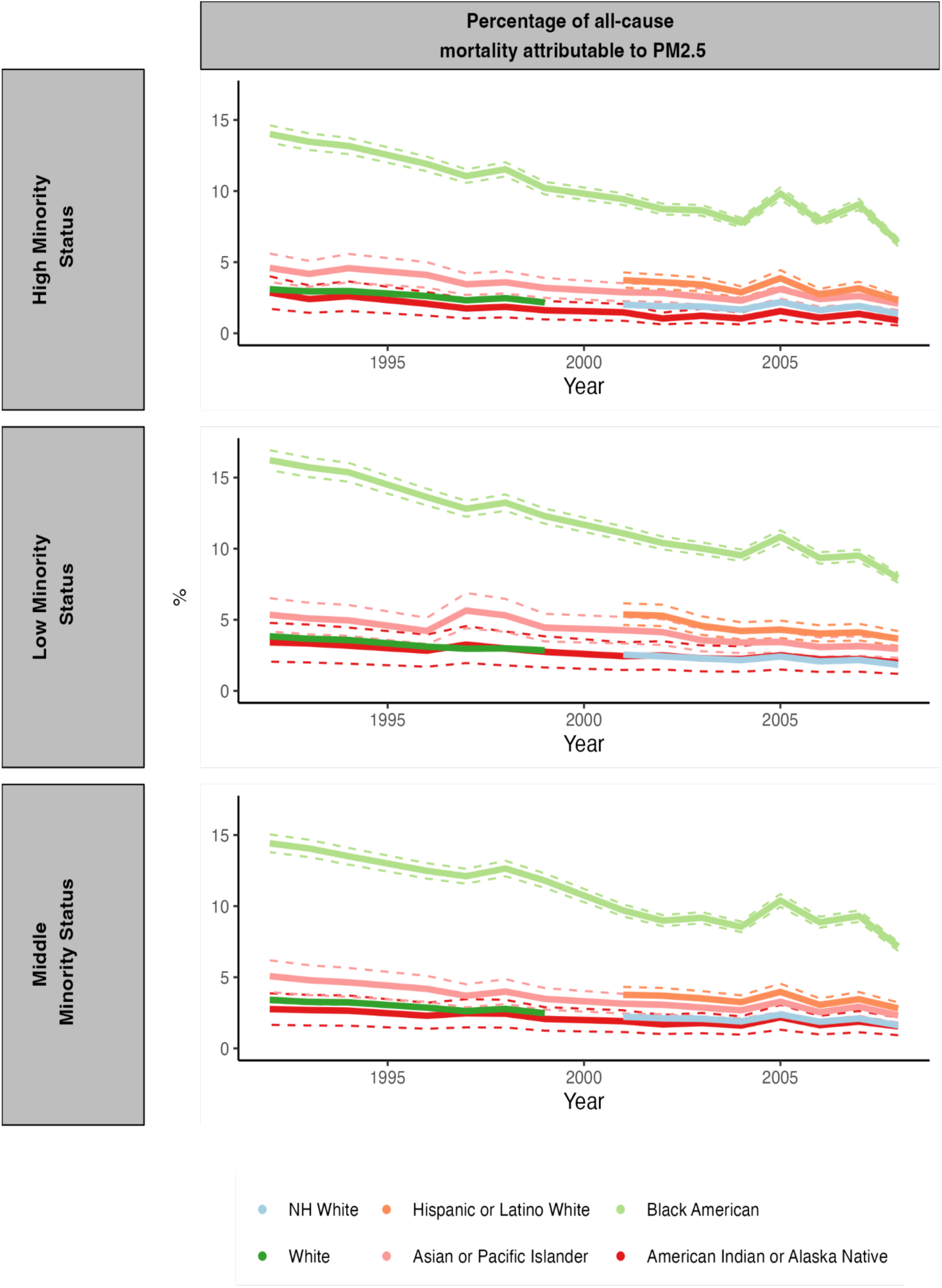
Percentage of all-cause mortality attributable to PM_2.5_ for each racial/ethnic group stratified by minority status. The dashed lines depict 95% confidence intervals. Abbreviations: NH=Non-Hispanic.

**Fig. S 59.**
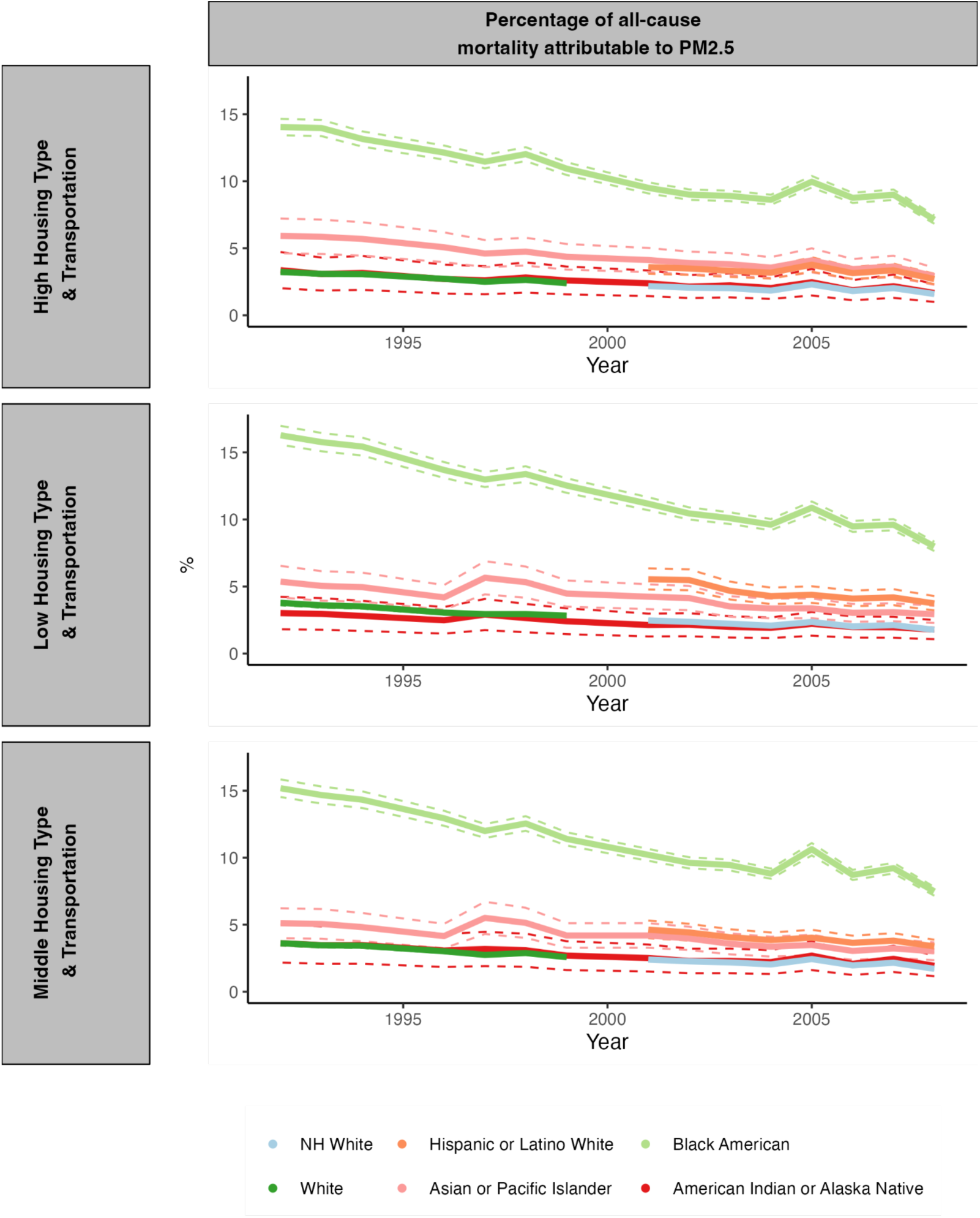
Percentage of all-cause mortality attributable to PM_2.5_ for each racial/ethnic group stratified by housing type & transportation. The dashed lines depict 95% confidence intervals. Abbreviations: NH=Non-Hispanic.

**Fig. S 60.**
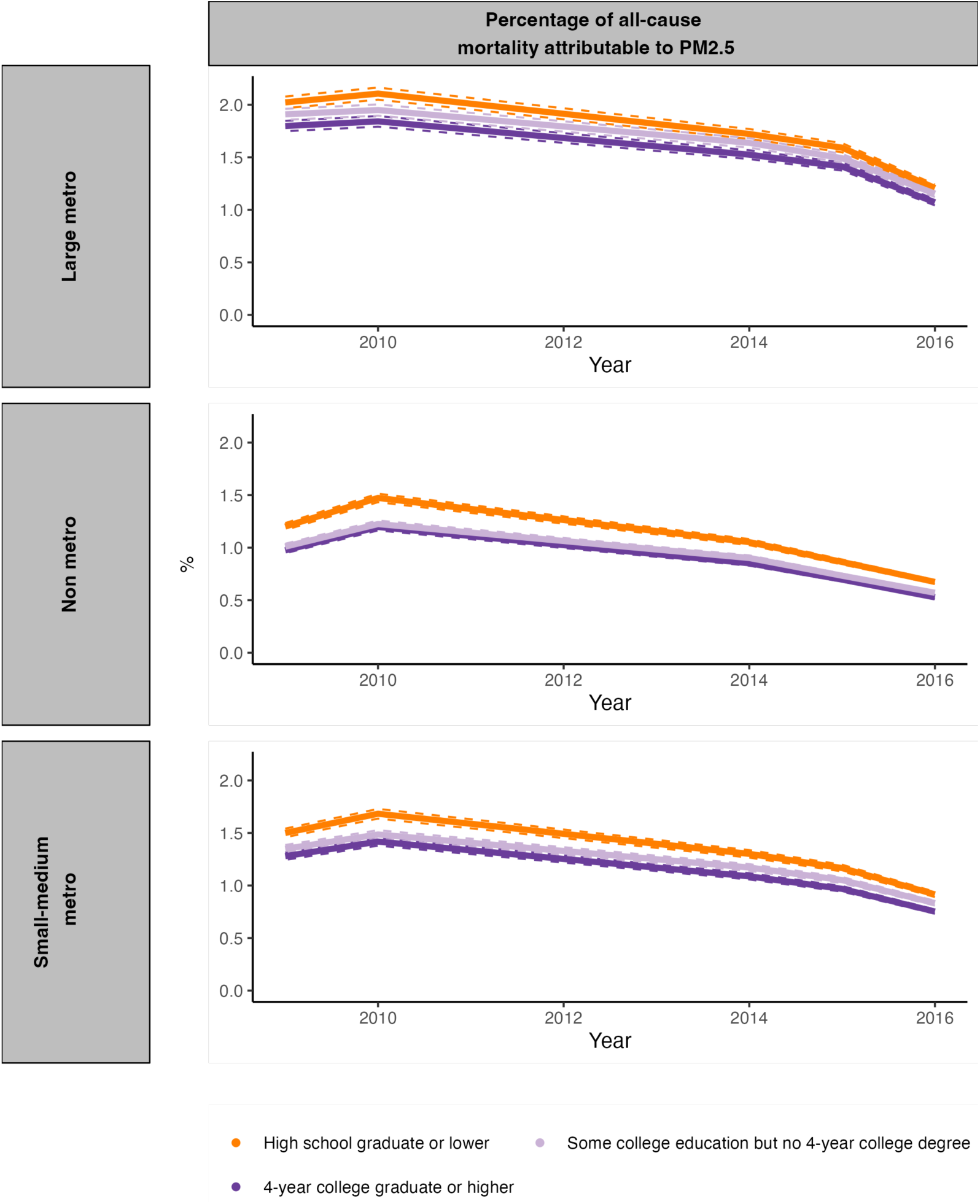
Percentage of all-cause mortality attributable to PM_2.5_ for each educational attainment level stratified by rurality. The dashed lines depict 95% confidence intervals. Abbreviations: NH=Non-Hispanic.

**Fig. S 61.**
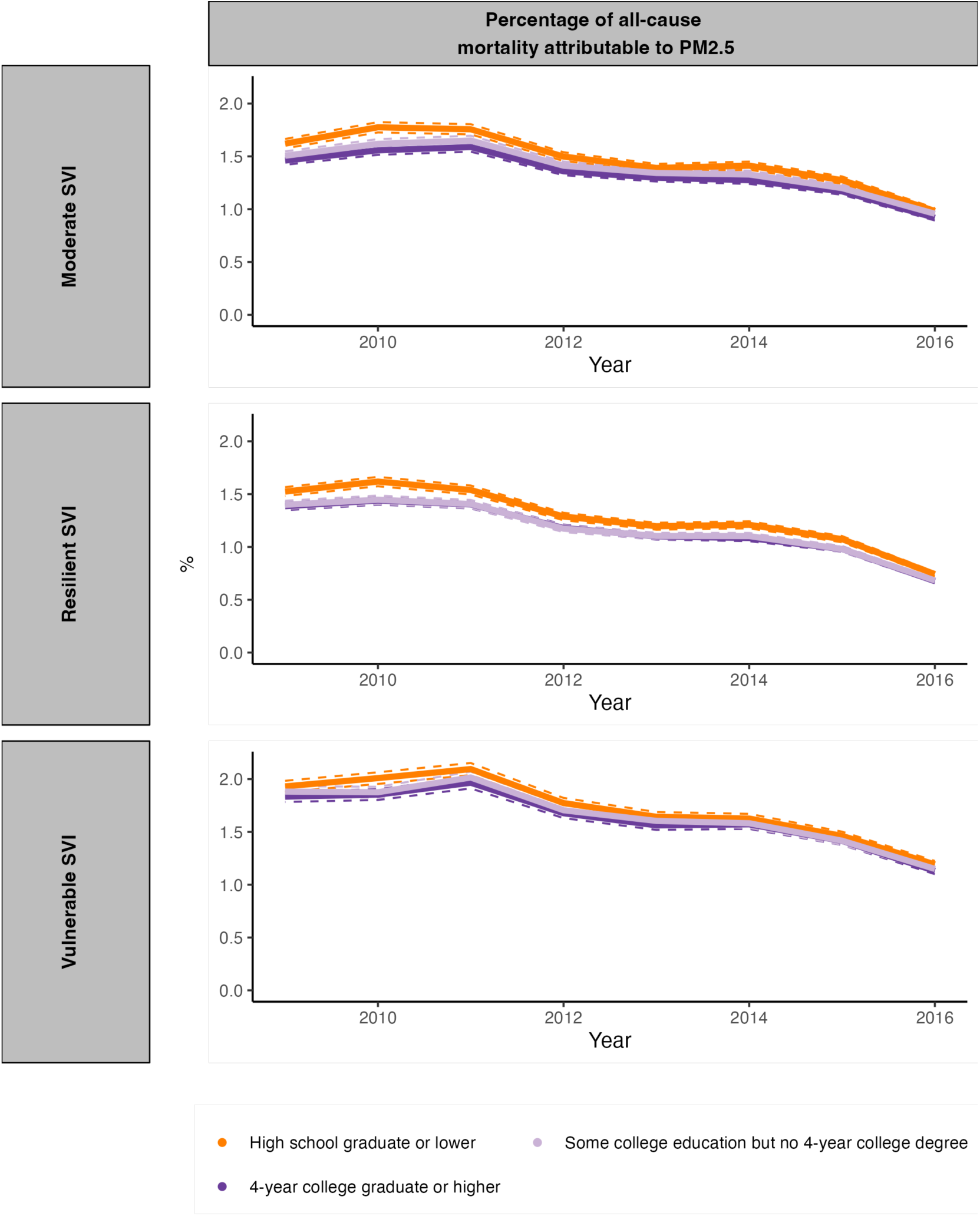
Percentage of all-cause mortality attributable to PM_2.5_ for each educational attainment level stratified by the social vulnerability index. The dashed lines depict 95% confidence intervals. Abbreviations: NH=Non-Hispanic.

**Fig. S 62.**
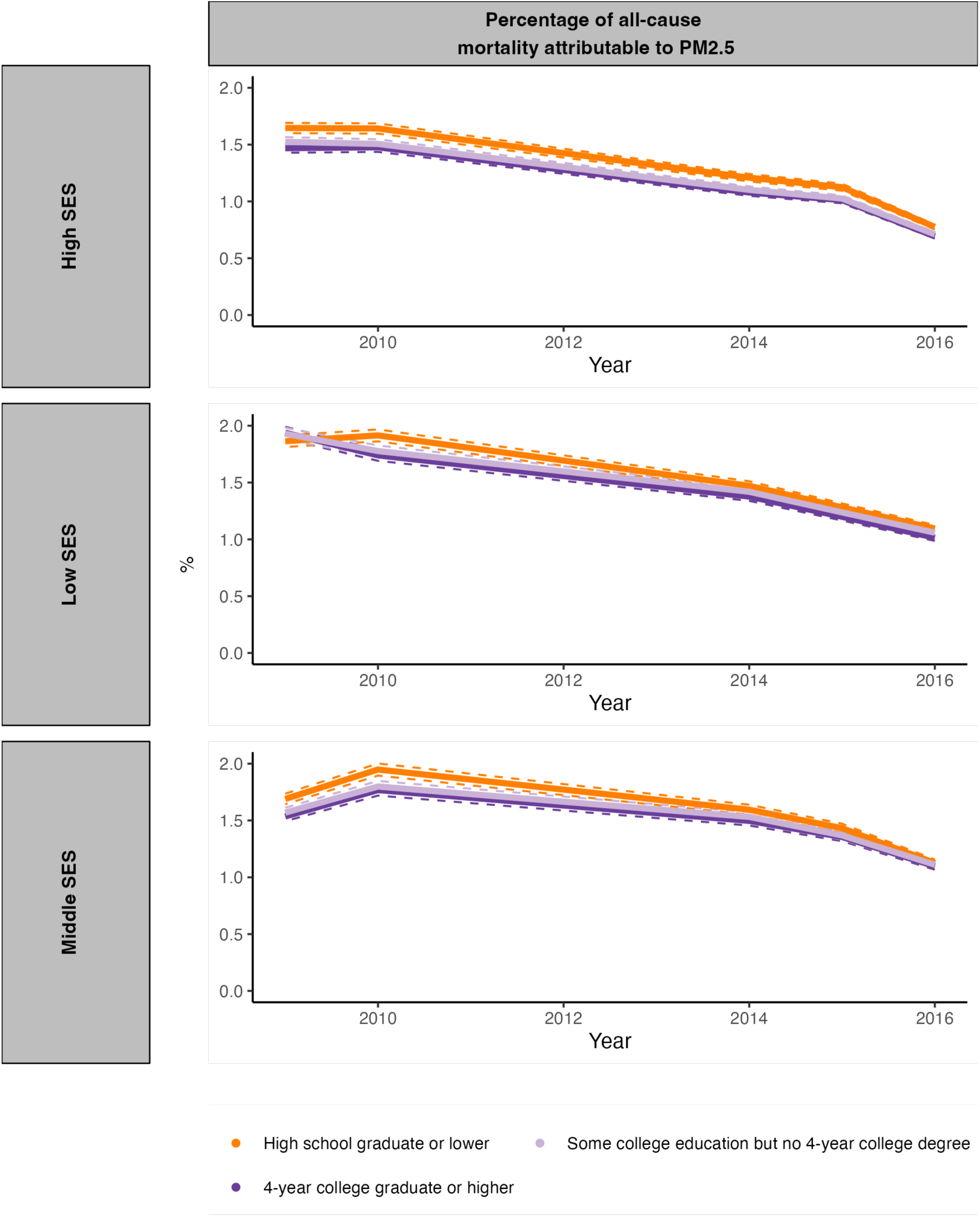
Percentage of all-cause mortality attributable to PM_2.5_ for each educational attainment level stratified by Socioeconomic Status. The dashed lines depict 95% confidence intervals. Abbreviations: NH=Non-Hispanic.

**Fig. S 63.**
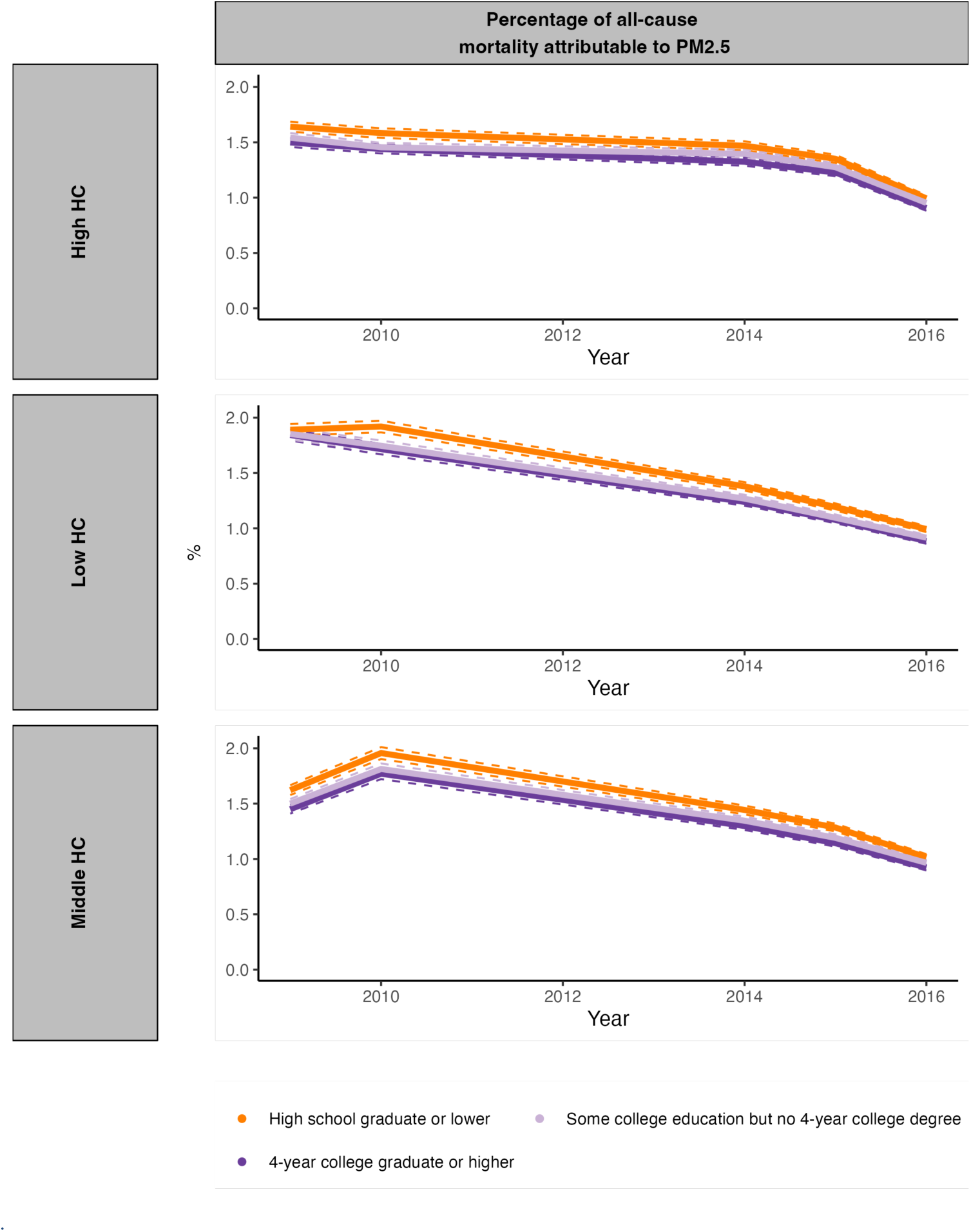
Percentage of all-cause mortality attributable to PM_2.5_ for each educational attainment level stratified by Household Characteristic. The dashed lines depict 95% confidence intervals. Abbreviations: NH=Non-Hispanic

**Fig. S 64.**
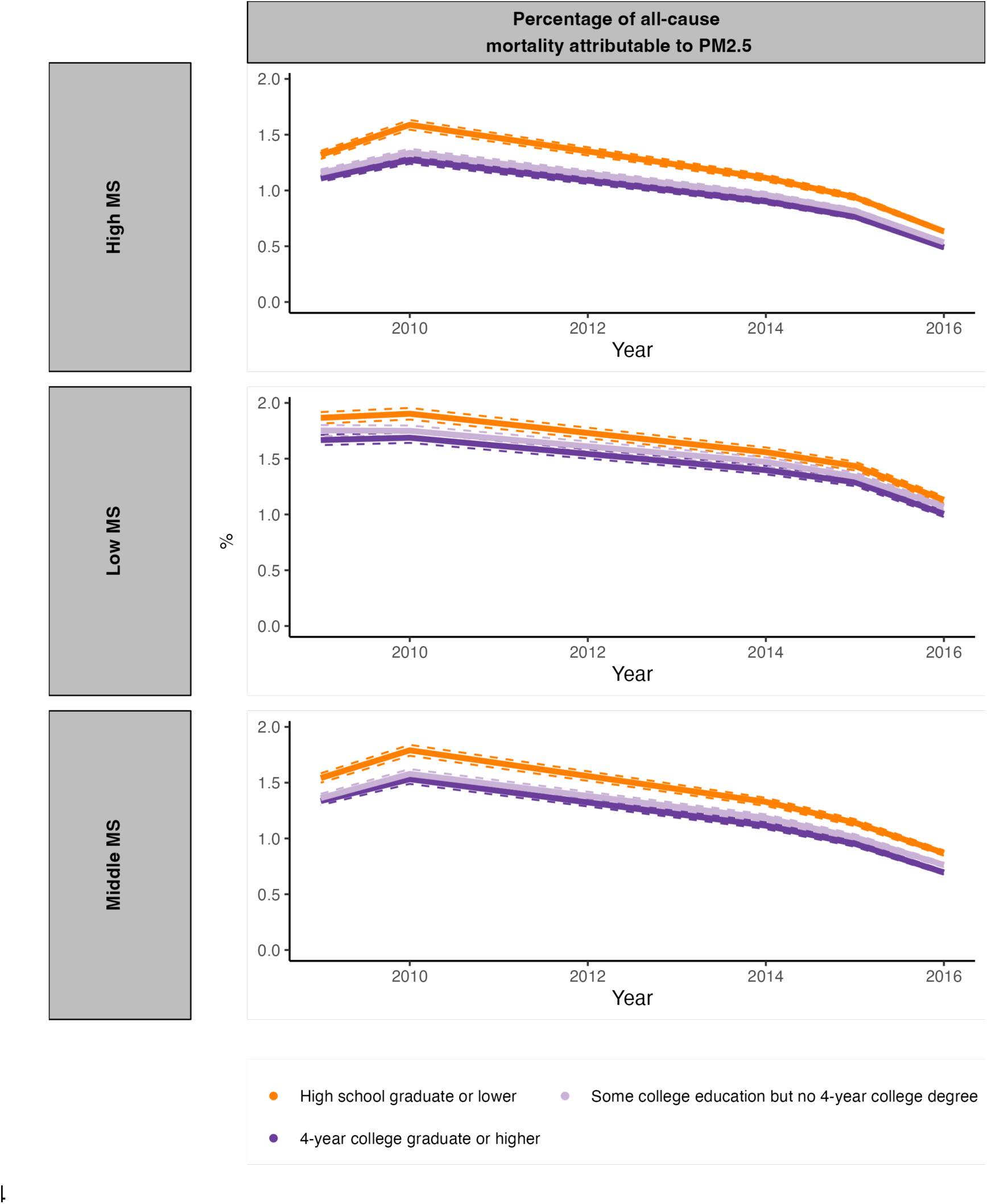
Percentage of all-cause mortality attributable to PM_2.5_ for each educational attainment level stratified by Minority Status. The dashed lines depict 95% confidence intervals. Abbreviations: NH=Non-Hispanic.

**Fig. S 65.**
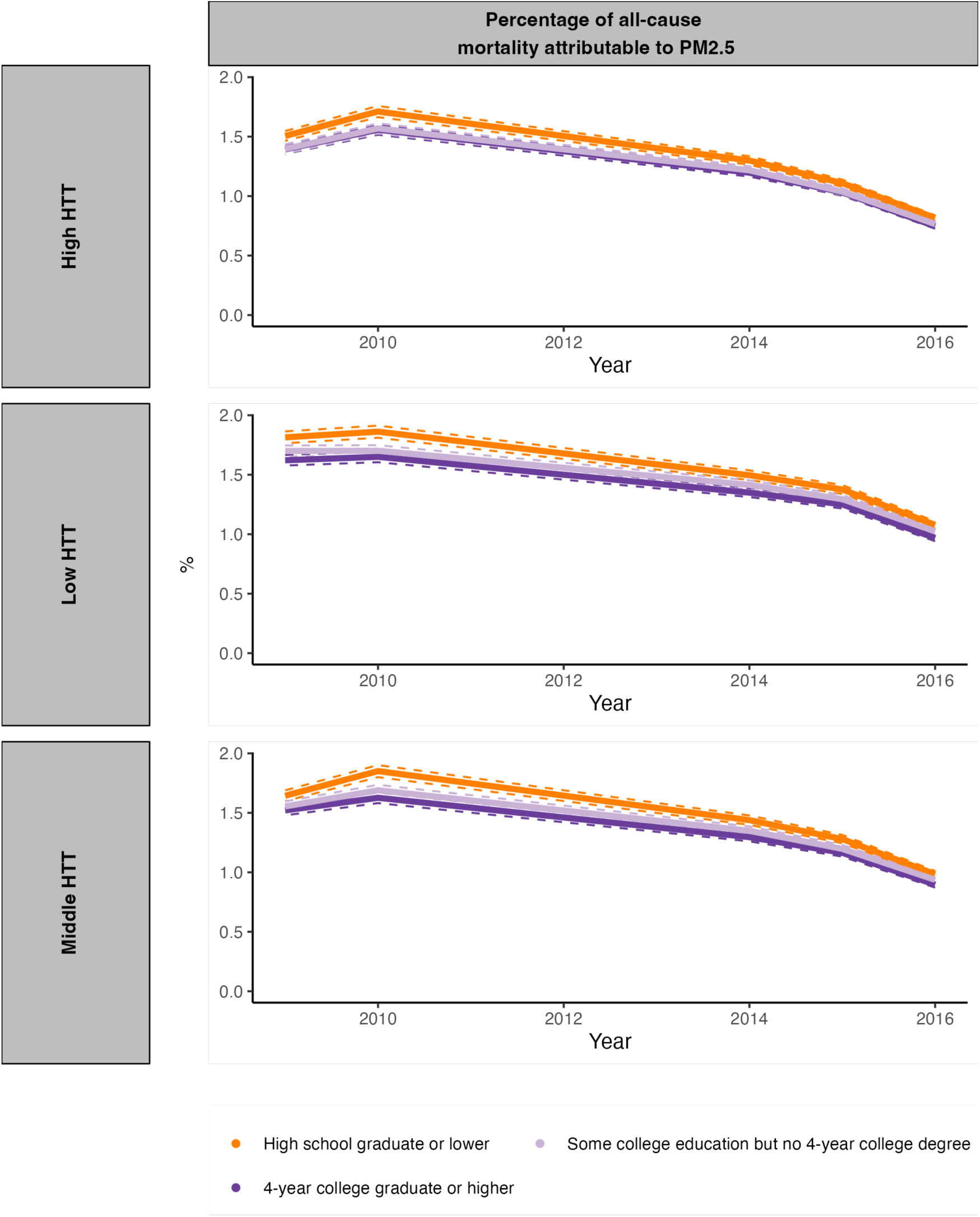
Percentage of all-cause mortality attributable to PM_2.5_ for each educational attainment level stratified by Housing Type & Transportation. The dashed lines depict 95% confidence intervals. Abbreviations: NH=Non-Hispanic.

**Table S 1.**
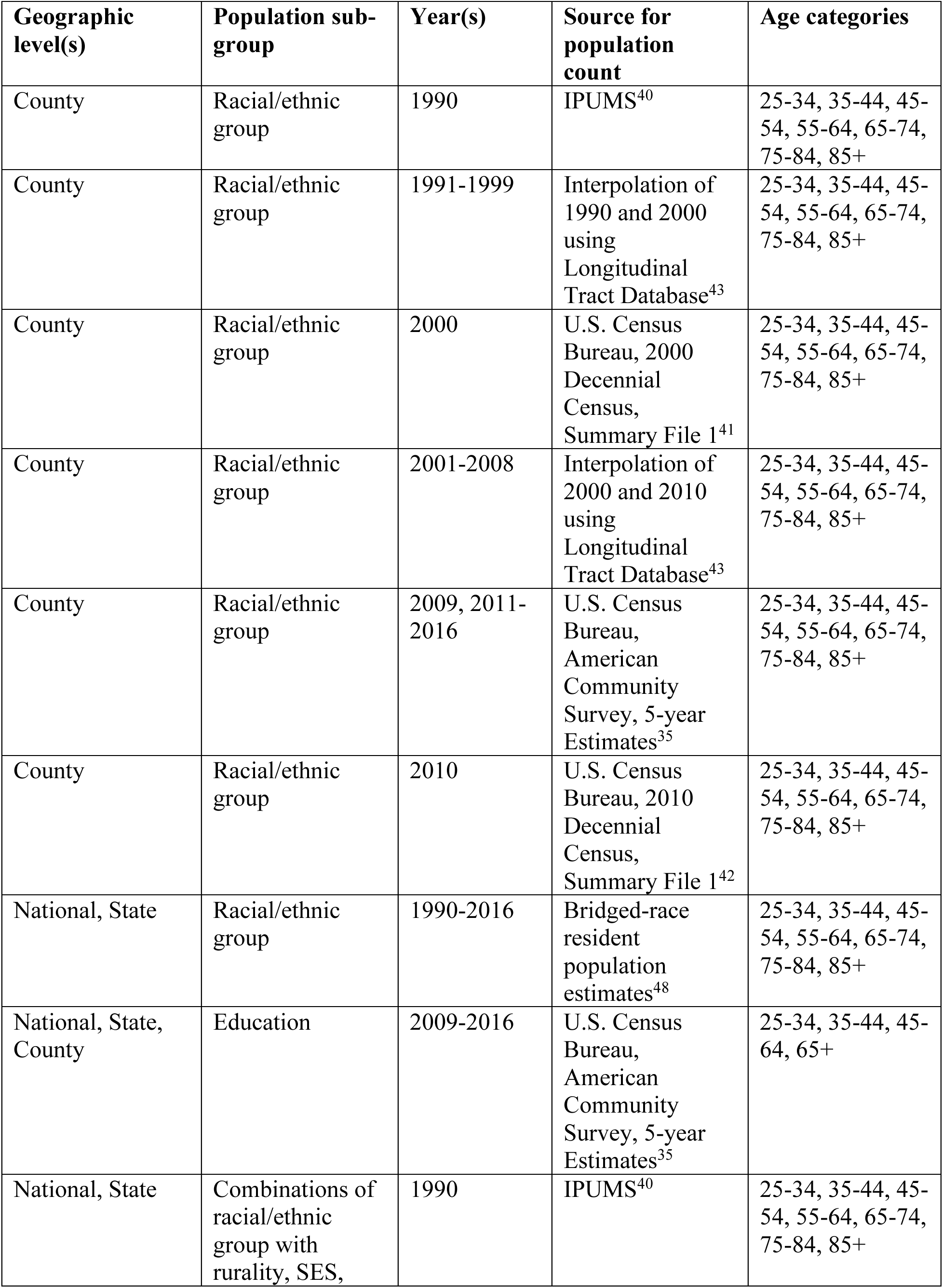
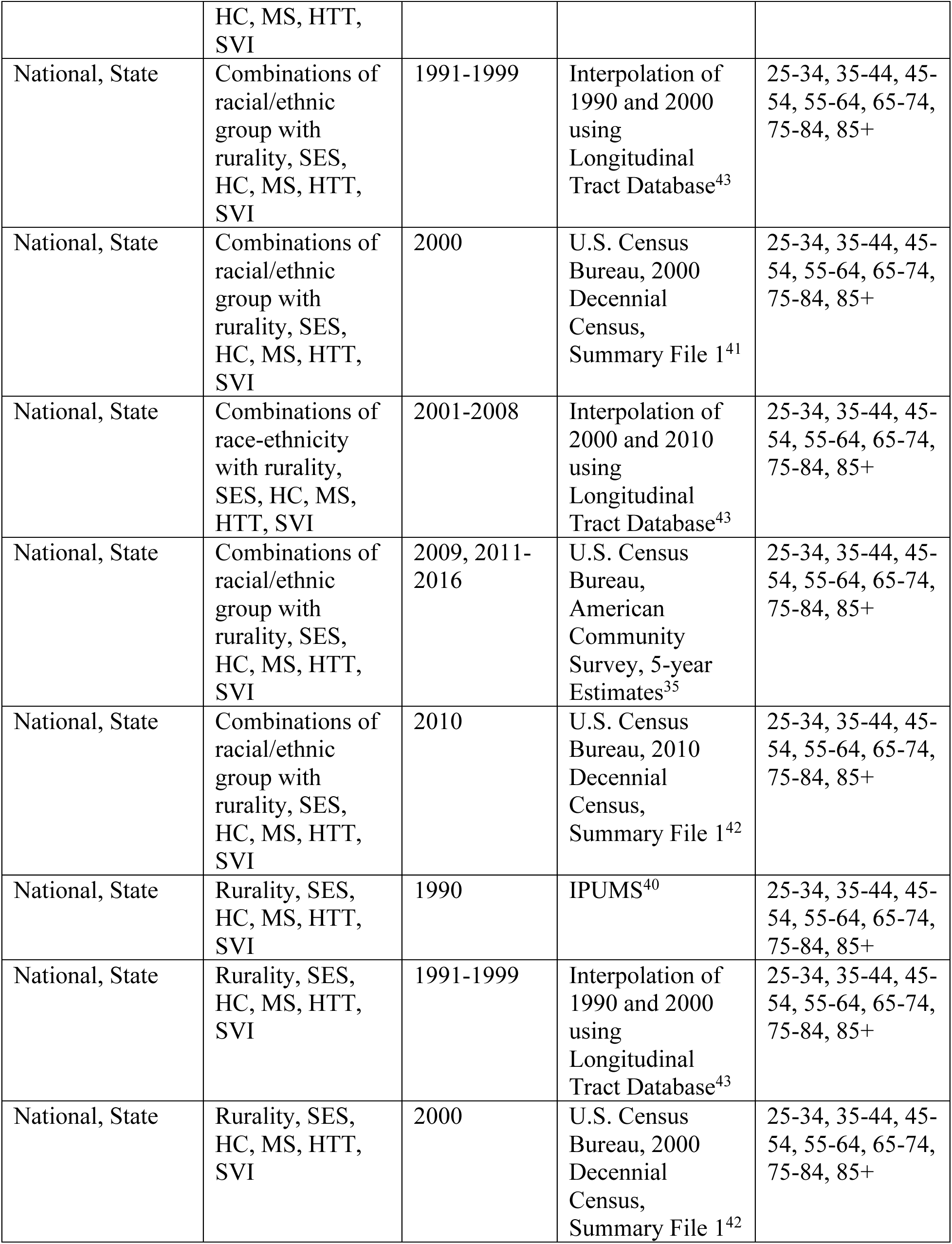
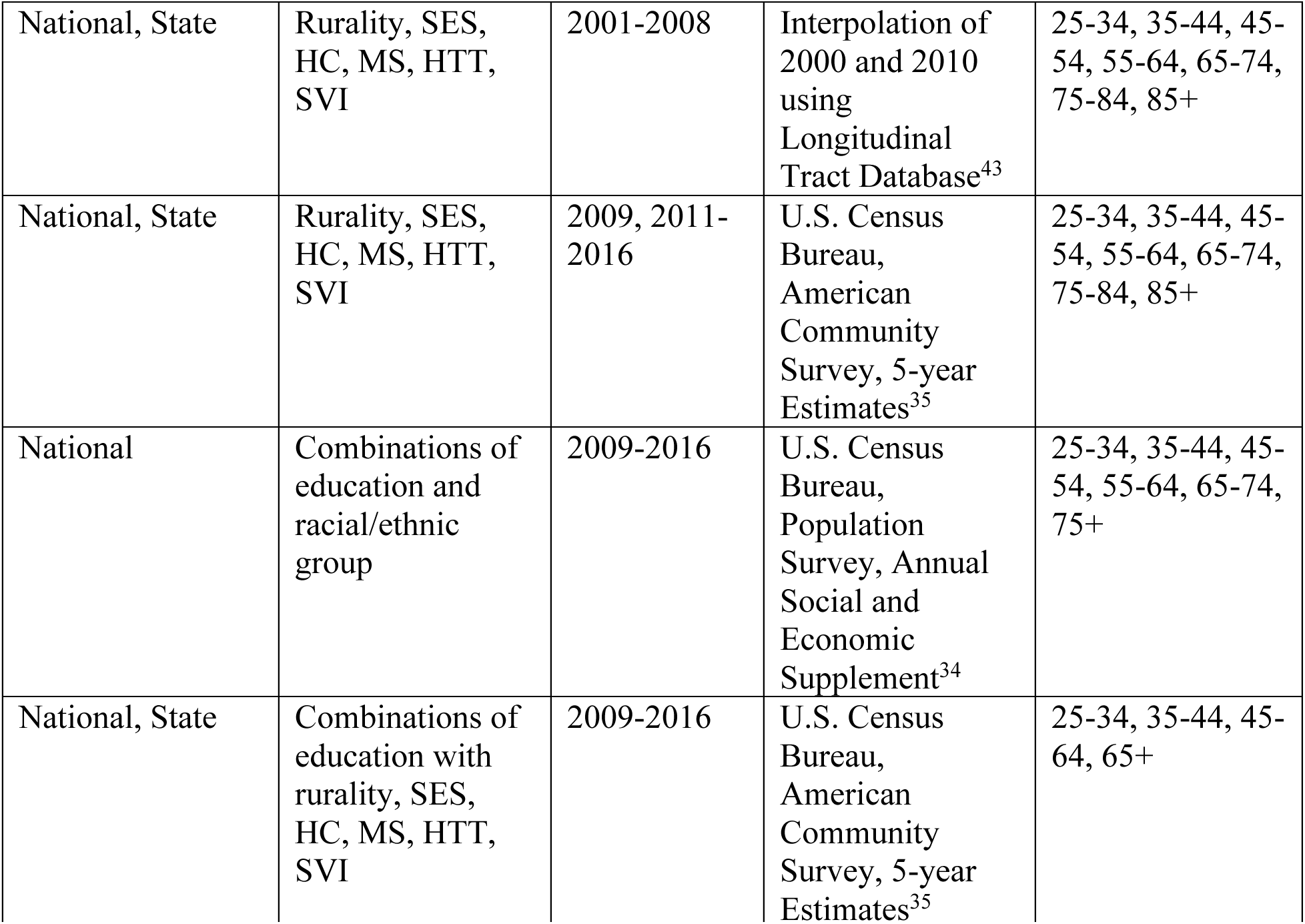
Data sources for the population counts used in the calculation of age-adjusted mortality rates. The US 2000 standard population table, which is also used for the calculation in the National Vital Statistics Report, has the age categories 25-34, 35-44, 45-54, 55-64, 65-74, 75-84, and 85+. Population counts that were available in more granular age categories were aggregated to these coarser categories. Abbreviations: SES = Socioeconomic Status, HC = Household Characteristics, MS = Minority Status, HTT = Housing Type & Transportation, SVI = Social Vulnerability Index.

**Table S 2.**
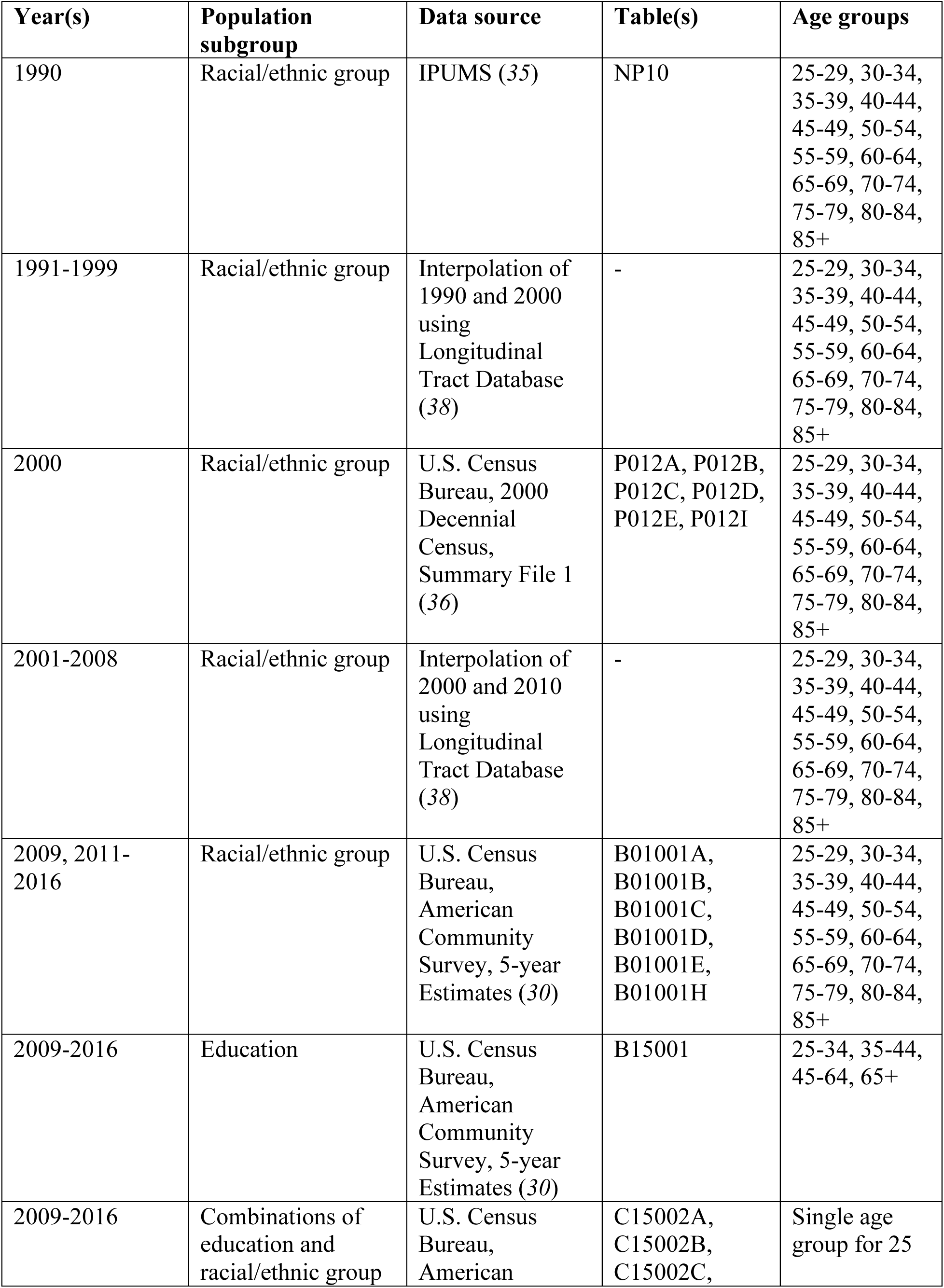
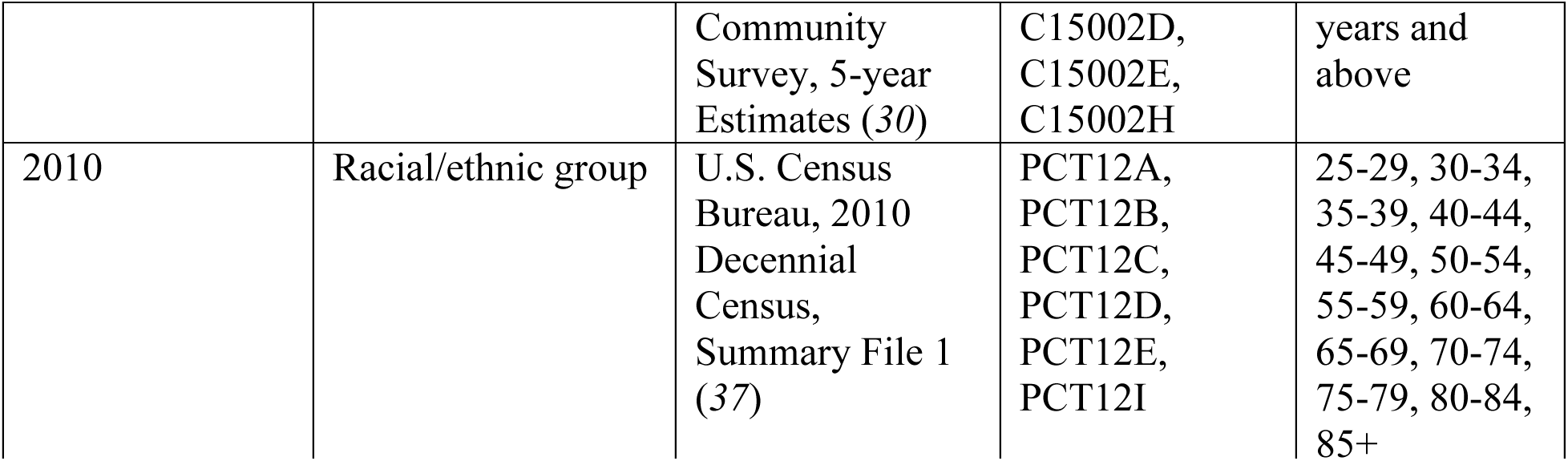
Data sources for the population counts used in the estimation of population-weighted mean PM_2.5_ exposure. All these data sources were at the census-tract level. Data from the U.S. Census Bureau is available at https://data.census.gov/cedsci/. Data from IPUMS is available at https://data2.nhgis.org/main.

**Table S 3.**
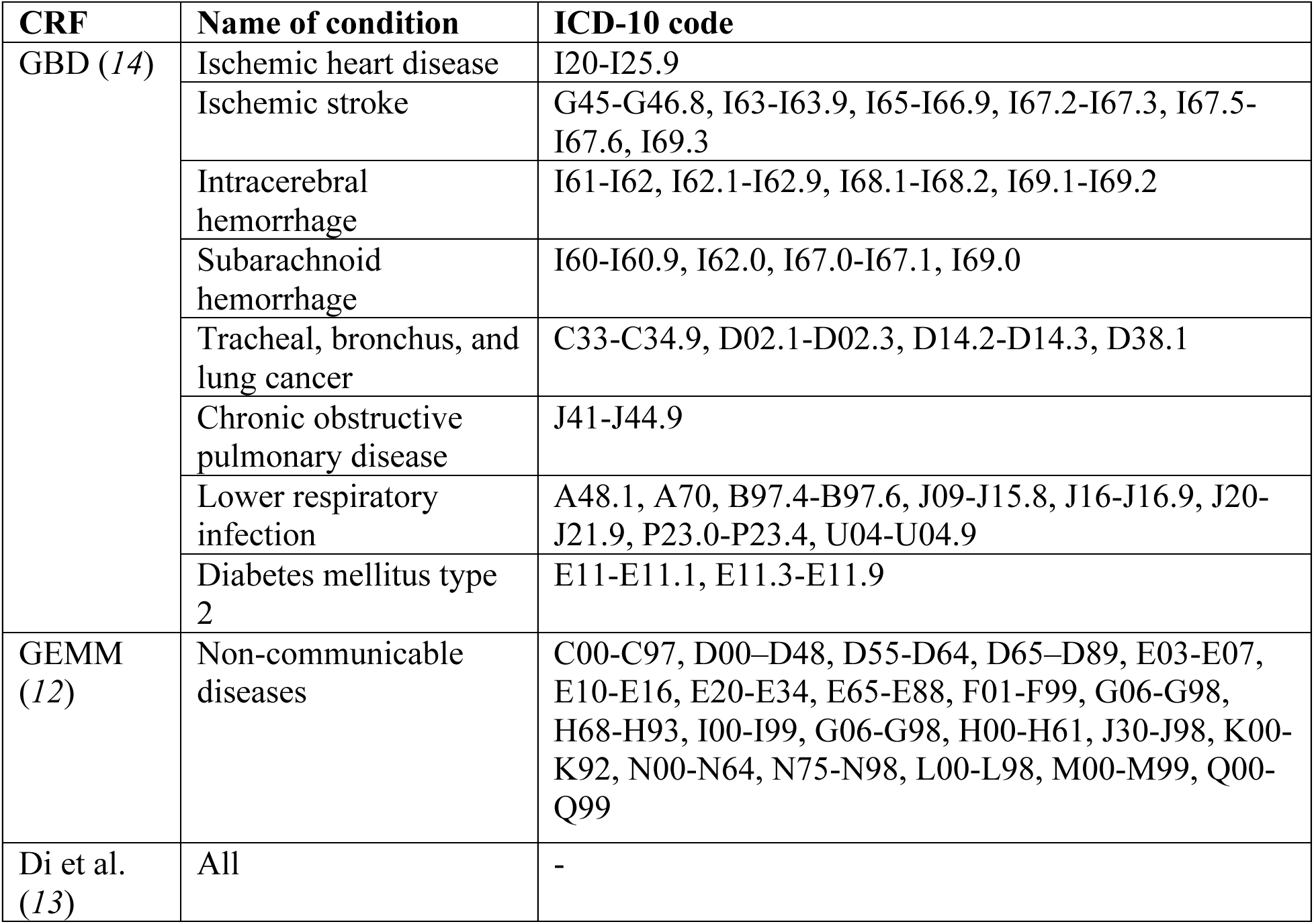
ICD-10 codes used in each concentration-response function. Abbreviations: CRF=concentration-response function.

